# Emergence and Expansion of the SARS-CoV-2 Variant B.1.526 Identified in New York

**DOI:** 10.1101/2021.02.23.21252259

**Authors:** Medini K. Annavajhala, Hiroshi Mohri, Pengfei Wang, Manoj Nair, Jason E. Zucker, Zizhang Sheng, Angela Gomez-Simmonds, Anne L. Kelley, Maya Tagliavia, Yaoxing Huang, Trevor Bedford, David D. Ho, Anne-Catrin Uhlemann

## Abstract

Recent months have seen surges of SARS-CoV-2 infection across the globe with considerable viral evolution^1-3^. Extensive mutations in the spike protein may threaten efficacy of vaccines and therapeutic monoclonal antibodies^4^. Two signature mutations of concern are E484K, which plays a crucial role in the loss of neutralizing activity of antibodies, and N501Y, a driver of rapid worldwide transmission of the B.1.1.7 lineage. Here, we report the emergence of variant lineage B.1.526 that contains E484K and its alarming rise to dominance in New York City in early 2021. This variant is partially or completely resistant to two therapeutic monoclonal antibodies in clinical use and less susceptible to neutralization by convalescent plasma or vaccinee sera, posing a modest antigenic challenge. The B.1.526 lineage has now been reported from all 50 states in the US and numerous other countries. B.1.526 rapidly replaced earlier lineages in New York upon its emergence, with an estimated transmission advantage of 35%. Such transmission dynamics, together with the relative antibody resistance of its E484K sub-lineage, likely contributed to the sharp rise and rapid spread of B.1.526. Although SARS-CoV-2 B.1.526 initially outpaced B.1.1.7 in the region, its growth subsequently slowed concurrent with the rise of B.1.1.7 and ensuing variants.

## Main

While evolution of SARS-CoV-2 was deemed to be slow at the beginning of the global pandemic^5^, multiple major variants of concern have emerged over the past year^1-3,6^. These lineages are each characterized by numerous mutations in the spike protein, raising concerns that they may escape from therapeutic monoclonals and vaccine-induced antibodies. The hallmark mutation of B.1.1.7, a SARS-CoV-2 variant of concern that emerged in the UK, is N501Y located in the receptor-binding domain (RBD) of spike^1^. This variant is seemingly more transmissible and virulent^7-9^, perhaps due to a higher binding affinity of N501Y for ACE2^10^ or a greater propensity to evade host innate immune responses^11^. Two other variants of concern, B.1.351^2^ and P.1^12^, share the N501Y mutation with B.1.1.7 but also contain an E484K substitution in RBD^2,3^. P.1 emerged as part of a second surge in Manaus, Brazil despite a high pre-existing SARS-CoV-2 seroprevalence in the population^13^. Reinfections with P.1 and another related Brazilian variant P.2 harboring E484K, have been documented^14,15^. Our previous study on B.1.351 demonstrated that this variant is refractory to neutralization by a number of monoclonal antibodies directed to the top of RBD, including several that have received emergency use authorization^4^. B.1.351 was markedly more resistant to neutralization by convalescent plasma and vaccinee sera. Importantly, these effects were in part mediated by the E484K mutation. These finding are worrisome in light of recent reports that three vaccine trials showed a substantial drop in efficacy in South Africa^16,17^. Likewise, P.1 was also relatively resistant to antibody neutralization, although not as severely^18^. We therefore implemented rapid molecular screening for signature mutations implicated in the success of these early variants of concern.

### Rapid screening for SARS-CoV-2 mutations

We first developed rapid PCR-based single-nucleotide-polymorphism (SNP) assays (Extended Data Fig. 1) to search for N501Y and E484K mutations in SARS-CoV-2 positive clinical samples stored in the Columbia University Biobank. Between November 1, 2020 and May 1, 2021, 1,602 samples were successfully genotyped by PCR. We identified 182/1,602 (11%) samples with E484K and 63/1,602 (3.9%) with N501Y. Eight samples contained both mutations. The earliest case with E484K was collected in mid-November 2020. The proportion of E484K PCR-screened cases substantially increased from 2.0% at the end of 2020 to 24.3% between February 21^st^ and March 5^th^, 2021 (Fig. 1a), when targeted PCR genotyping was replaced by whole-genome sequencing. Viruses harboring N501Y also increased over time, from the earliest detection in mid-January to 5.3% of screened isolates by the beginning of March.

**Figure 1.**
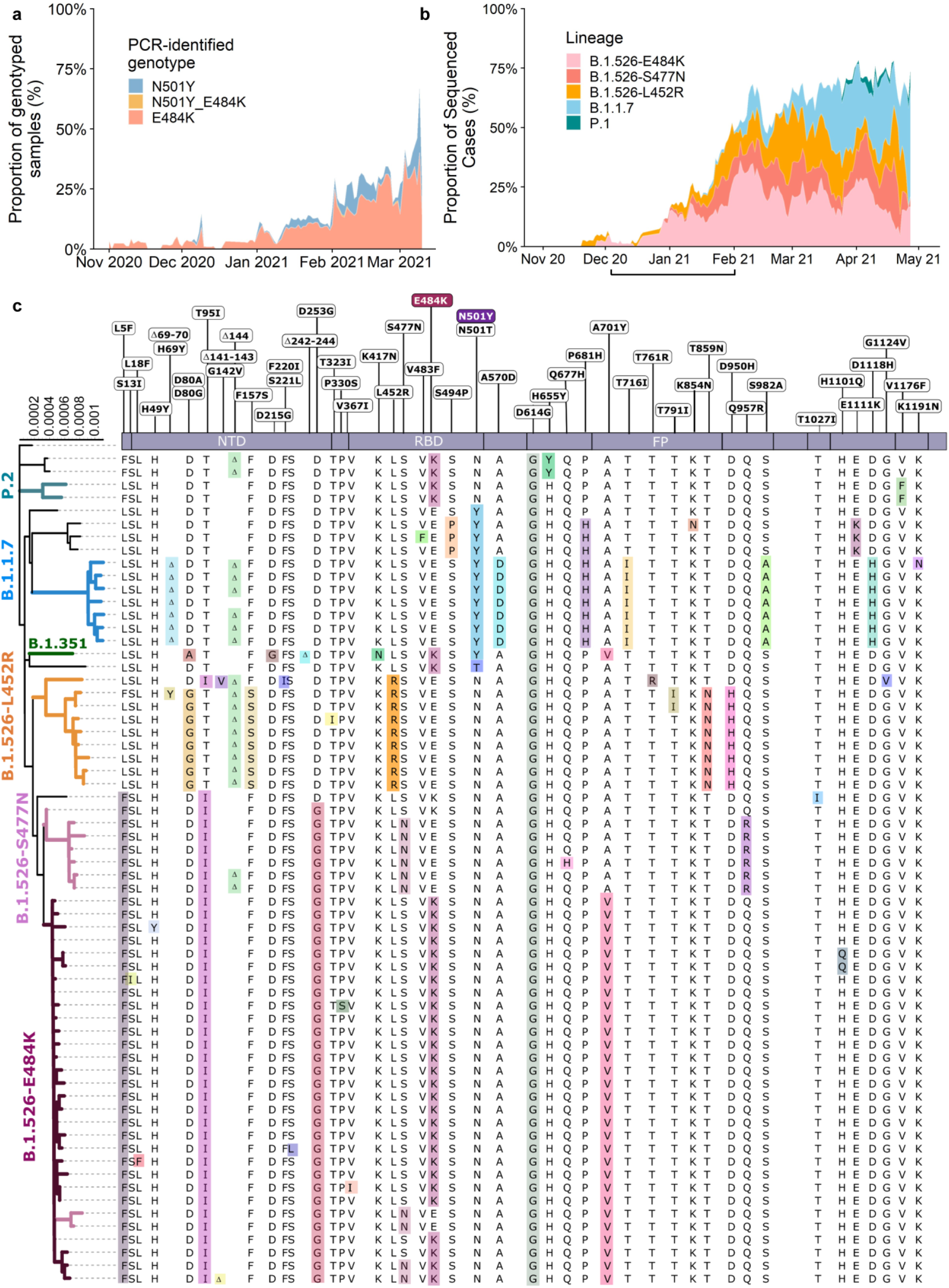
Prevalence of E484K-harboring SARS-CoV-2 and B.1.526. **(a)** Detection of viruses with key signature mutations in spike over time. The earliest detected E484K-harboring variant was collected in mid-November 2020. The prevalence of E484K (samples with E484K/total PCR-genotyped samples) subsequently increased over time, from 4.8% in early December 2020 up to 24.3% in early March 2021. Throughout late 2020 and early 2021, we identified fewer N501Y-than E484K-harboring isolates, with a maximum of 5.9% of N501Y during mid-February 2021. **(b)** Distribution of different viral lineages identified by whole genome sequencing. Within our genomic collection (n=1,507), the B.1.526 lineage rose rapidly in early 2021, replacing the majority of other lineages (shown as the white blank space) present during this timeframe. This was followed by a steady rise in B.1.1.7 by mid-2021. The marking below the X axis denotes the time-period used to calculate the growth advantage of B.1.526 over other earlier viruses. **(c)** Phylogenetic tree of SARS-CoV-2 variants identified by sequencing and alignment of key spike mutations. Unique patterns of spike protein mutations present in genomes sequenced from our hospital center with at least one mutation of interest or concern (E484K, N501Y, S477N, or L452R; n=64) are shown. Residues at which at least one sample harbored a mutation are displayed above the S-protein schematic.

### Genomic surveillance of SARS-CoV-2

We next performed untargeted whole genome nanopore sequencing of nasopharyngeal samples collected throughout the study period with cycle threshold (Ct)≤35. We successfully obtained 1,507 SARS-CoV-2 whole genomes (59% of samples with Ct≤35; Extended Data Fig. 2). Sequencing results verified the E484K and N501Y substitutions in all samples identified by PCR screening. Of sequenced N501Y isolates, 31/41 (76%) were consistent with the B.1.1.7 lineage. Samples which harbored both N501Y and E484K were genotyped as P.1 (n=6), B.1.351 (n=1), and B.1.623 (n=1). However, quite unexpectedly, the large majority of PCR-screened cases with E484K (n=98/128, 77%) fell within a single lineage, B.1.526,^19^ recently labeled the Iota variant by the WHO^20^.

Analysis of the entire collection of CUIMC genomic sequences (Fig. 1b) showed that by May 2021, SARS-CoV-2 variants (including B.1.526, B.1.1.7, and more recently P.1) comprised two-thirds of all sequenced isolates, replacing the vast majority of earlier lineages (Fig. 1b). The proportion of cases caused by B.1.526 rose rapidly from late 2020 through February 2021, and remained at approximately 40-50% of all sequenced cases from March to May 2021, despite a concurrent increase in B.1.1.7. In fact, during the months of December and January when the prevalence of B.1.1.7 was still negligible (Fig. 1b, marking under horizontal axis), the frequency of all viruses in the B.1.526 lineage rose from <5% to 50% while the frequency of other lineages declined from >95% to 50% (Fig. 1b, where white blank space represents other lineages). Calculations using these numbers in a head-to-head comparison and an established mathematical method^21^ indicate that B.1.526 has a growth advantage of ∼5% per day. Likewise, fitting a logistic regression model to 478 individual observations from the extended timeframe of November 2020 through January 2021 shows that B.1.526 had a similar growth advantage of 4.6% per day (95% CI 2.8–6.5% per day). Given that the serial interval for SARS-CoV-2 transmission is about 7 days^22^ in the absence of any intervention, these results suggest that B.1.526 is ∼35% more transmissible than non-variant viruses.

Demographic and clinical features, including clinical outcomes, were largely comparable in patients with E484K versus those without the signature E484K or N501Y mutations, and between patients with B.1.526-E484K versus those with non-variant lineages^23^ (Extended Data Table 1). However, significantly lower Ct values were associated with both E484K (29.49 vs 30.71, p=0.013) and B.1.526-E484K (27.65 vs 28.81 in non-variant lineages, p=0.015), indicating a modestly higher viral load in these variant samples. A significantly higher proportion of patients B.1.526-E484K were also admitted to the hospital or presented to the emergency department (p=0.037).

### Signature B.1.526 lineage mutations

We identified signature spike-protein mutations in the B.1.526 lineage by comparing all genomes generated in this study (Fig. 1c). Phylogenetic examination showed that the B.1.526 lineage is comprised of two closely related sub-lineages harboring either E484K (B.1.526-E484K; defined as Pangolin lineage B.1.526) or S477N (B.1.526-S477N; Pangolin lineage B.1.526.2), and the additional sub-lineage B.1.526.1, harboring the L452R substitution (B.1.526-L452R). Both B.1.526-E484K and B.1.526-S477N share characteristic spike-protein mutations L5F, T95I, D253G, D614G, and either A701V or Q957R along with either E484K or S477N. Non-spike mutations widely shared by B.1.526-E484K and B.1.526-S477N isolates include: T85I in ORF1a-nsp2; L438P in ORF1a-nsp4, a 9bp deletion Δ106-108 in ORF1a-nsp6; P323L in ORF1b-nsp12; Q88H in ORF1b-nsp13; Q57H in ORF3a; and P199L and M234I in the N gene. While B.1.526-L452R isolates shared a number of mutations across the genome in ORF-1ab, ORF-3ab, ORF-8, and N, it does not share characteristic spike mutations with B.1.526-E484K and B.1.526-S477N.

To further investigate the evolutionary history of B.1.526, we performed phylogenetic analyses on genomes in this collection and in GISAID harboring the ORF1a-nsp6 deletion Δ106-108, along with mutation A20262G that uniquely defines the parent clade containing B.1.526 and related viruses (Fig. 2a). We observed a stepwise emergence of the key lineage-defining mutations, with T95I, D253G, and L5F appearing in the earliest phylogenetic nodes. Isolates subsequently branched into four sub-lineages, with two major groups B.1.526-E484K and B.1.526-S477N containing A701V, with a smaller sub-lineage B.1.526-S477N containing Q957R. The B.1.526-L452R lineage, which also emerged in parallel, is related to B.1.526-E484K and B.1.526-S477N yet forms a distinct phylogenetic branch (Fig. 1c).

**Figure 2.**
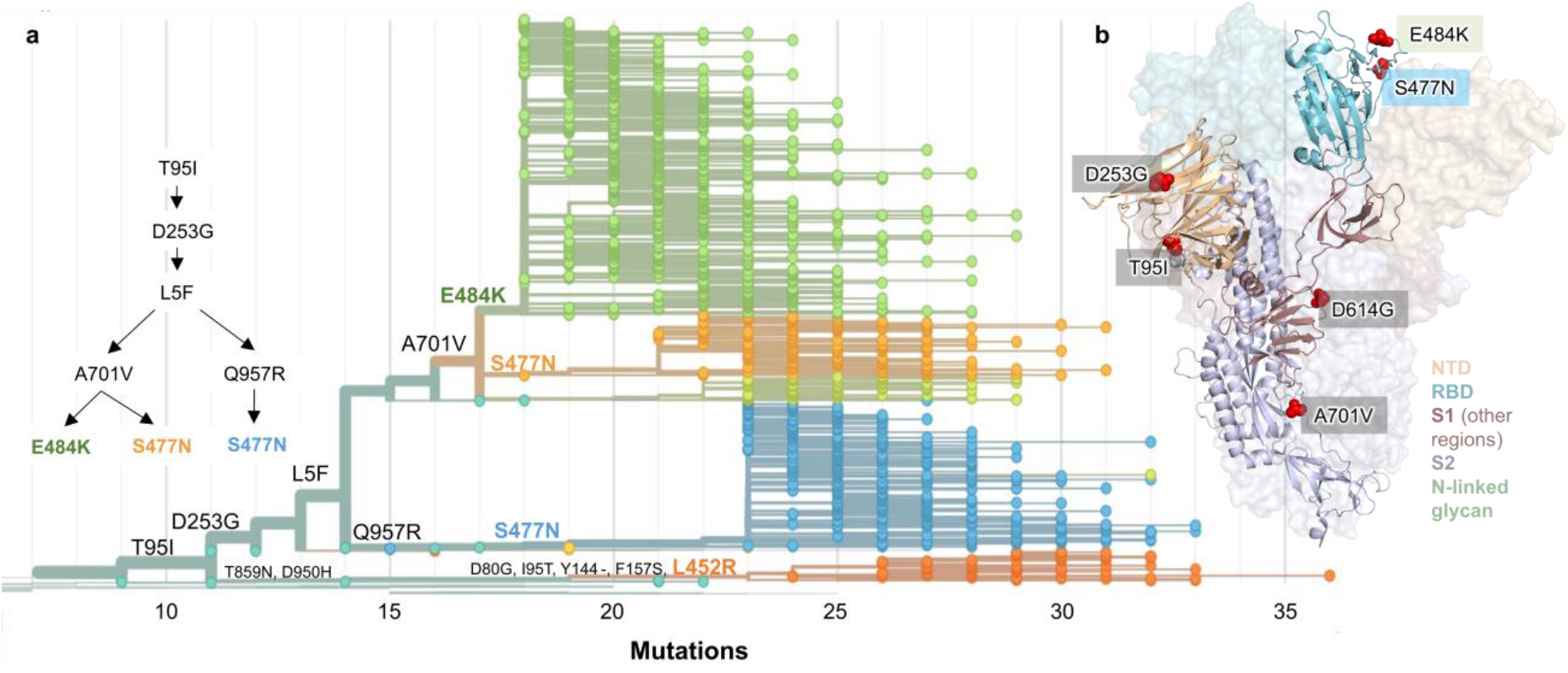
Spike protein amino acid substitutions and structural changes represented in sequenced isolates. **(a)** Maximum-likelihood phylogenetic tree of 2,309 SARS-CoV-2 viruses colored according to spike protein haplotype. Spike protein mutations are labeled on the tree showing the stepwise accumulation of signature B.1.526 mutations T95I, D253G, and L5F, and branching of B.1.526-E484K (green) and two B.1.526-S477N sub-lineages (light orange, blue). The B.1.526-L452R sub-lineage (dark orange) emerged in parallel. An interactive version of this figure is available at https://nextstrain.org/groups/blab/ncov/ny/B.1.526. **(b)** Key mutations of B.1.526 displayed on the spike trimer. The D253G mutation resides in the antigenic supersite within the N-terminal domain (NTD), a target for neutralizing antibodies, E484K and S477N at the receptor binding domain (RBD) interface with the cellular receptor ACE2, and A701V near the furin cleavage site.

Fig. 2b displays the localization of signature B.1.526-E484K and B.1.526-S477N mutations within the S protein. D253G resides in the antigenic supersite within the N-terminal domain^24^, which is a target for neutralizing antibodies^25^, whereas E484K is situated at the RBD interface with the cellular receptor ACE2. The A701V mutation near the furin cleavage site is also shared with variant B.1.351.

### Antibody neutralization of B.1.526

The impact of the signature S protein mutations in B.1.526 on antibody neutralization was first assessed using vesicular stomatitis virus (VSV)-based pseudoviruses as previously described^4,25^. Pseudoviruses containing S477N or E484K alone and all five signature mutations (L5F, T95I, D253G, A701V, and E484K or S477N), termed NYΔ5(E484K) or NYΔ5(S477N), were constructed and subjected to neutralization by 12 monoclonal antibodies including 5 with emergency use authorization, 20 convalescent plasma, and 22 vaccinee sera. The specifics of these monoclonal antibodies and clinical specimens were previously reported^4^. The neutralizing activity of 12 monoclonal antibodies covering a range of epitopes on RBD was essentially unaltered against the S477N and NYΔ5(S477N) pseudoviruses (Extended Data Fig. 3a) showing that this mutation has no discernible antigenic impact, as was confirmed using convalescent plasma and vaccinee sera (Extended Data Fig. 3b). However, against E484K and NYΔ5(E484K) pseudoviruses, the activities of several antibodies were either impaired or lost, including REGN10933 and LY-CoV555 that are already in clinical use (Fig. 3a). Likewise, neutralizing activities of convalescent plasma or vaccinee sera were lowered by 4.1-fold or 3.3-3.6-fold, respectively, against NYΔ5(E484K) (Fig. 3b). Neutralization studies of the authentic B.1.526-E484K virus yielded similar results, although the magnitude of resistance to convalescent plasma or vaccinee sera was slightly lower at 2.6-fold or 1.8-2.0-fold, respectively (Fig. 3b). A comparative analysis with other variants of concern (Fig. 3c) showed that such risks are likely lower than B.1.351 and closer to P.1. Overall, these results demonstrate the need to modify our antibody therapy and to monitor the efficacy of current vaccines in regions where B.1.526-E484K is prevalent.

**Figure 3.**
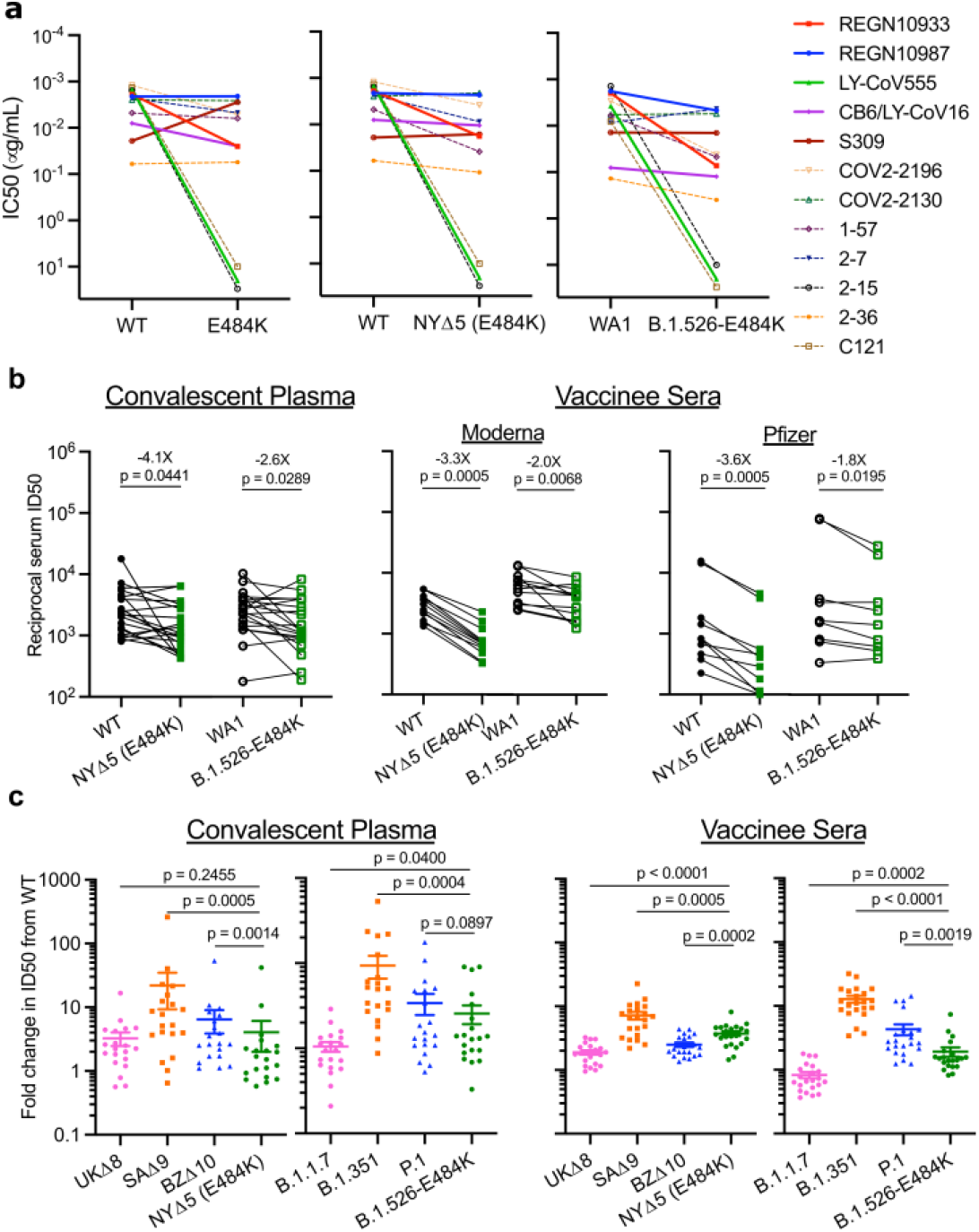
Neutralization studies of B.1.526-E484K and comparative analyses. **(a)** Neutralizing activities of 12 monoclonal antibodies against pseudoviruses containing E484K alone or all five signature B.1.526 mutations (L5F, T95I, D253G, A701V, and E484K), termed NYΔ5(E484K) as well as against the authentic B.1.526-E484K. Antibodies with emergency use authorization are shown in bold solid lines. Data are represented as mean ± SEM of technical triplicates and represent one of two independent experiments. **(b)** Neutralizing activities of convalescent plasma (n=20) and vaccinee sera (n=22) against the NYΔ5(E484K) pseudovirus compared to wildtype pseudovirus as well as against authentic B.1.526-E484K and wildtype virus (WA1). **(c)** Fold change in convalescent plasma and vaccinee sera neutralization ID50 of different variant pseudoviruses and live viruses compared to wildtype counterparts. The data on B.1.1.7, B.1.351 and P.1 were derived from our prior publications^4,18^. Data from 20 convalescent patients or 22 vaccinated individuals were averaged and are represented as arithmetic mean ± SEM (individual data points also shown). Statistical comparisons were made using the Wilcoxon matched-pairs signed rank test; two-tailed p-values are reported.

### B.1.526 surge across New York and the US

Prevalence of the novel variant B.1.526 surged alarmingly in our hospital catchment area (Fig. 4a) and throughout New York State (Fig. 4b) after its emergence in late 2020, replacing other lineages and initially outpacing B.1.1.7. A multinomial logistic regression model describing the concurrent growth rates of these two lineages shows that starting in mid-April 2021, B.1.1.7 surpassed B.1.526 due to a slightly higher fitness, with estimated growth rates in New York State of 5.3% per day for B.1.1.7 (95% CI 5.0–5.7%) and 3.4% per day for B.1.526 (3.2–3.6%) (Fig. 4b). These estimates suggest a fitness advantage of B.1.526 over existing non-variant lineages of 22–25% over a serial interval of 7 days^21,22^ during a period when multiple variants are competing simultaneously. Furthermore, the estimates also suggest a fitness advantage of B.1.1.7 over existing non-variant lineages of 35–40%, as well as a fitness advantage of B.1.1.7 over B.1.526 of 12–15%. Both lineages grew quickly (Fig. 4a,b), but once they reached a high frequency of circulating viruses, the competition between them caused the growth of B.1.1.7 to slow and B.1.526 to decline.

**Figure 4.**
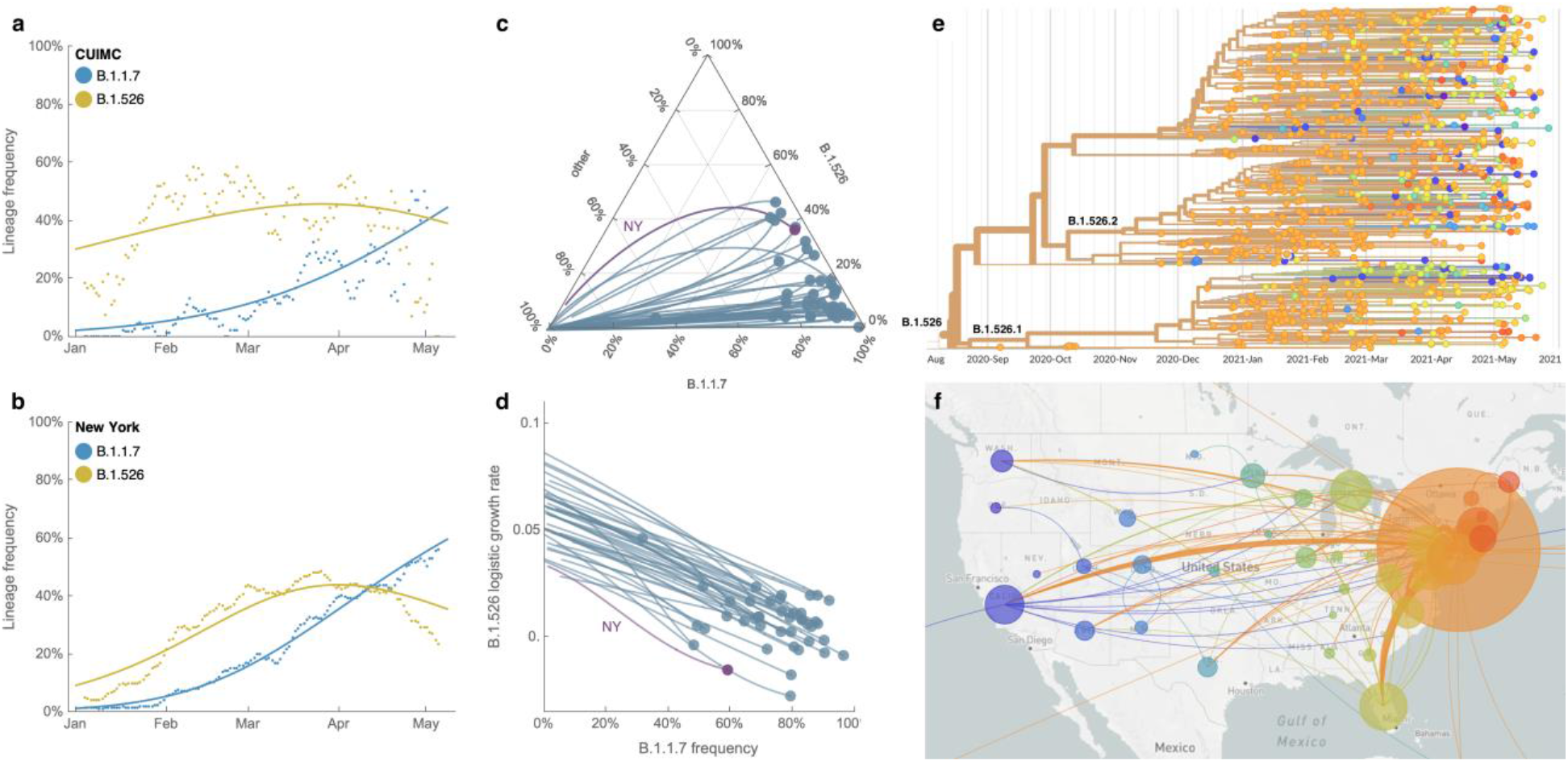
Spread of lineages B.1.1.7 and B.1.526 in New York and the USA. **(a, b)** Frequencies of lineages B.1.1.7 (blue) and B.1.526 (yellow) in the CUIMC catchment area (in panel a) and New York State (in panel b) with dots representing daily 7-day sliding window averages and lines representing fit to a multinomial logistic regression model. **(c)** Ternary plot of state-level frequency trajectories for 42 states separating frequencies of B.1.1.7, B.1.526 and other lineages. Each state-level trajectory is a line in this plot moving from lower left in January 2021 when both B.1.1.7 and B.1.526 were rare, rightward as B.1.1.7 and B.1.526 increase in frequency. The trajectory of New York State is highlighted in purple. **(d)** The same data as in panel c, except plotting frequency of B.1.1.7 against logistic growth rate of B.1.526. (e) Phylogenetic tree of 933 B.1.526 samples from across the US where branch tips are colored based on location of sampling and branches are colored by inferred ancestral location. (f) Phylogeographic view of data from panel e, where each sampling location is represented as a circle with area proportional to sample count and each inferred transition event across the phylogeny is drawn as an arc connecting inferred origin and destination. Most migration events are inferred to be direct dispersals from New York State.

Frequency trajectories of B.1.1.7 and B.1.526 across states (Fig. 4c, Extended Data Fig. 4) show two general patterns: (1) initial rapid increase of both lineages until the proportion of other lineages had been eclipsed, followed by decline of B.1.526 seen in New York and in several neighboring states; and (2) rapid growth and resulting dominance of B.1.1.7 preventing the further rise of B.1.526. The dynamics between these two lineages is further shown in Fig. 4d, which plots the logistic growth rate of B.1.526 against the frequency of B.1.1.7, again at the state-level. At lower frequencies of B.1.1.7, all states have a similarly rapid growth of B.1.526 as it replaces non-variant lineages. As B.1.1.7 increases in frequency, however, it slows the growth of B.1.526, again indicative of a slightly higher fitness for B.1.1.7. At a minimum, B.1.526 rose rapidly where B.1.1.7 was not already dominant and, in several states, continued to grow at a similar pace as B.1.1.7 (Extended Data Fig. 4).

Phylogeographic analysis of the B.1.526 lineage revealed ancestral viruses originating in New York in August 2020, diversifying within the state, and then dispersing to other states (Figs. 4e and 4f). State-level genomic data showed that B.1.526 was concentrated primarily in New York and surrounding states, including New Jersey and Rhode Island (Extended Data Fig. 4). This suggests that B.1.526, and B.1.526-E484K in particular, became widespread in the region, the original epicenter of COVID-19 in the US^26,27^, although the lineage has also grown in states outside the Northeastern US (e.g., North Carolina). By the end of April 2021, the geographic makeup of B.1.526 within the US was quite diverse, and the lineage has emerged and expanded in multiple states across the country (Fig. 4f). The rise of B.1.526 over a short timeframe across the United States (Extended Data Fig. 4), as well as its international spread, are notable.

## Discussion

Here we report the emergence of the SARS-CoV-2 lineage B.1.526, and its surge in New York during the second wave of the COVID-19 pandemic. Neutralization studies on B.1.526-E484K demonstrate that the activities of several antibodies were either impaired or lost, including two (Ly-CoV555 and REGN10933) already in clinical use. Furthermore, neutralizing activities of convalescent plasma or vaccinee sera were lower against B.1.526 harboring E484K (Fig. 3b). The S477N mutation, a key signature of another B.1.526 sub-lineage, on the other hand, did not have an impact on antibody neutralizing.

Several limitations of our study need to be considered. This was a single-center genomic survey representing patients presenting to a hospital system and may not have fully captured patients with milder disease. However, our results are comparable to genomic data released by public health laboratories in the region and incorporate all publicly available data for phylogeographic context and growth rate calculations. As in all genomic surveillance studies, we predominantly sequenced samples with a Ct<30 but covered a high proportion of samples throughout the study period. In addition, our PCR screen allowed us to obtain unbiased estimates of E484K and N501Y prevalence early on in the study. PCR approaches may be increasingly warranted for continued surveillance during non-surge periods, during which Ct values trend higher. Lastly, transmissibility estimates based on observed prevalence are imperfect as they reflect observed growth rates rather than intrinsic transmissibility of the virus.

Taken together, our findings underscore the importance of the E484K mutation, which has emerged in at least 246 different lineages of SARS-CoV-2^28^, a real testament to convergent evolution. This highlights that E484K can rapidly emerge in multiple clonal backgrounds and may warrant targeted screening for this key mutation in addition to robust genomic surveillance programs. However, B.1.526 is one of the few lineages with E484K that has risen to prominence. The greatest threat of B.1.526 appears to be its ease of spread, with an estimated transmissibility of ∼35% greater than non-variant viruses when competing head-to-head. Despite the notable transmissibility of B.1.1.7, B.1.526 was able to spread rapidly in the US to replace other lineages and continued to increase in frequency in several states where both B.1.526 and B.1.1.7 were predominant. Similarly, while B.1.351 may pose the greatest antigenic challenge to antibodies and vaccines, the B.1.526-E484K sub-lineage also exhibits resistance to antibody neutralization. The findings herein present a clear-cut example of SARS-CoV-2 evolution in real time. B.1.526, with its higher transmissibility, appeared suddenly and rose to dominance, only to wane as variants (B.1.1.7, and B.1.617.2 more recently) with even greater fitness emerged. These observations are a stark reminder that if SARS-CoV-2 is allowed to continue its spread, increasingly worrisome variants are to be expected in the future.

## Data Availability

All SARS-CoV-2 genomes generated as part of this study have been submitted to GISAID under submitter ID mka2136 and NCBI GenBank under BioProject PRJNA751551.

https://nextstrain.org/groups/blab/ncov/ny/B.1.526

## Methods

### Clinical cohort

This observational study took place at an academic quaternary care center in New York City. Nasopharyngeal swabs obtained as part of routine clinical care were tested by the Clinical Microbiology laboratory, and positive specimens were transferred to the Columbia University Biobank for inactivation and storage. Electronic health records data extracted for this analysis included demographics, laboratory results, admission, discharge, and transfer dates, current and historical international classification of disease (ICD 9 and 10) codes extracted from the clinical data warehouse. This study was reviewed and approved by the Columbia University Institutional Review Board (protocol number AAAT0123).

### PCR screening

Extended Data Figure 1 describes our overall protocol for variant screening. To enable rapid PCR-based screening, we prepared RNA using the heat inactivation method in place of RNA isolation methods^29^. First, 50 µl of nasal swab sample in VTM solution was transferred into 96-well PCR plates, covered with an adhesive aluminum foil (VWR 60941-076) and incubated at 95°C for 5 min using the PCR instrument. After the centrifugation of the plate at >2,100 x g for 5 min, 5 µl of the supernatant from each sample, which contains viral RNA, was used for the SNP assay.

The SNP assay consists of four steps as follows: reverse transcription (RT) of viral RNA, pre-read of the SNP assay, real-time PCR and post-read of the SNP assay. 5 µl of RNA from the supernatant was added to 15 µl of the single step RT-qPCR reaction mix, which consists of 5 µl of TaqPath 1-step RT-qPCR Master Mix, CG (4x) (ThermoFisher Scientific), 500 nM of forward and reverse primers, 120 nM of VIC-MGB probe, 50 nM of FAM-MGB probe, 1/2000 volume of ROX Reference Dye (Invitrogen) as the final concentration, and nuclease-free water to adjust the total reaction volume of 20 µl. Each reaction plate included 8 control wells, 5×10^6^ and 5×10^3^ copies of WA-1 (wild type), UK variant and South African variant, which were generated by PCR to match the variant sequences, and 2 wells with water as no template controls (NTC).

The primer pairs and probes used are as follows. For the SNP assay for position **501**, a primer pair of 501.F: 5’-GGT TTT AAT TGT TAC TTT CCT TTA CA-3’ and 501.R: 5’-AGT TCA AAA GAA AGT ACT ACT ACT CTG TAT G-3’ were used with two TaqMan probes (ThermoFisher Scientific), one for wild type, VIC.N501MGB: [VIC]-AA CCC ACT AAT GGT-MGBNFQ and the other for variant type, FAM.Y501MGB: [FAM]-AAC CCA CTT ATG GT-MGBNFQ. For position **484**, a primer pair of 484.F: 5’-AGA GAG ATA TTT CAA CTG AAA TCT ATCAGG-3’and 484.R: 5’-GAA ACC ATA TGA TTG TAA AGG AAA GTA AC-3’ were used with two probes, one for wild type, VIC.E484MGB: [VIC]-ATG GTG TTG AAG GT-MGBNFQ and the other for variant type, FAM.K484MGB: [FAM]-ATG GTG TTA AAG GT-MGBNFQ.

The reaction plate was subjected to 1) reverse-transcription reaction (RT) at the condition at 25°C for 2 min, at 50°C for 15 min and a hold at 4°C; 2) SNP assay (pre-read) at 60°C for 30 sec; 3) real-time PCR at 95°C for 20 sec followed by 50 cycles of two-step PCR, at 95°C for 3 sec and at 60°C for 30 sec with the fast 7500 mode; followed by 4) SNP assay (post-read) at 60°C for 30 sec using ABI 7500 Fast Dx Real-Time PCR Instrument with SDS Software (ThermoFisher Scientific). The genotype at each key position for each sample was determined by reading the component signal of the amplification and the allelic discrimination analysis software in the program.

### Whole genome sequencing

Extended Data Fig. 2 displays a flowchart outlining samples available for this study. Isolates with cycle threshold (Ct) values below 35 were selected for sequencing using the ARTIC v3 low-cost protocol targeting 400bp amplicons^30^ or Rapid Barcoding kit protocol targeting 1,200bp amplicons^31^. Briefly, RNA was extracted using the Qiagen RNeasy Mini kit or Zymo DNA/RNA Mini kit. Reverse transcription was performed using LunaScript RT SuperMix (NEB). Tiling PCR was performed on the cDNA, and amplicons were barcoded using the Oxford Nanopore Native Barcoding Expansion 96 kit. Pooled barcoded libraries were then sequenced on an Oxford Nanopore MinION sequencer using R9.4.1 flow cells. Basecalling was performed in the MinKNOW software v21.02.1. Sequencing runs were monitored in real-time using RAMPART (https://artic-network.github.io/rampart/) to ensure sufficient genomic coverage with minimal runtime. Consensus sequence generation was performed using the ARTIC bioinformatics pipeline (https://github.com/artic-network/artic-ncov2019). Genomes were manually curated by visually inspecting sequencing alignment files for verification of key residues in Geneious v10.2.6.

### Phylogenetic analysis

Phylogenetic reconstruction of amino acid changes (Fig. 2A) was conducted using the Nextstrain^32^ workflow at https://github.com/nextstrain/ncov which aligns sequences against the Wuhan-Hu-1 reference via nextalign (https://github.com/nextstrain/nextclade), constructs a maximum-likelihood phylogenetic tree via IQ-TREE^33^, estimates molecular clock branch lengths via TreeTime^34^ and reconstructs nucleotide and amino acid changes also via TreeTime. This workflow was applied to 2309 SARS-CoV-2 genomes possessing the 9bp deletion Δ106-108 in ORF1a-nsp6 along with mutation A20262G which demarcates the parent clade to lineage B.1.526 alongside 688 global reference viruses. This analysis was conducted on data downloaded from gisaid.org^35^ on April 5, 2021. Phylogeographic reconstruction of spread from New York state (Fig. 4E-F) was similarly conducted using the same Nextstrain workflow with the addition of performing ancestral trait reconstruction of the geographic “division” attribute of 933 SARS-CoV-2 genomes downloaded from gisaid.org on Jun 6, 2021.

### Neutralization studies of pseudoviruses

We assayed the neutralizing activity of monoclonal antibodies (mAbs), convalescent plasma, and vaccinee sera against E484K, S477N, and WT (D614G) pseudoviruses, as well as pseudovirus NYΔ5 containing all five signature mutations of B.1.526-E484K (L5F, T95I, D253G, E484K, D614G, A701V), as previously described^25^. We examined four mAbs with emergency use authorization (CB6, REGN10987, REGN10933 and LY-CoV555), plus eight additional RBD mAbs, including ones from our own collection (2-15, 2-7, 1-57, & 2-36)^25^ as well as S309^36^, COV2-2196 & COV2-2130^37^, and C121^38^, We also examined convalescent plasma collected in Spring of 2020 (n=20 patients), and Moderna and Pfizer vaccinee sera (n=22)^4^. Briefly, Vero E6 cells (ATCC) were seeded in 96-well plates (2 ×10^4^ cells per well). Pseudoviruses were incubated with serial dilutions of the test samples in triplicate for 30 min at 37 °C. The mixture was added to cultured cells and incubated for an additional 24 h. Luminescence was measured using a Britelite plus Reporter Gene Assay System (PerkinElmer), and IC_50_ was defined as the dilution at which the relative light units were reduced by 50% compared with the virus control wells (virus + cells) after subtraction of the background in the control groups with cells only. The IC_50_ values were calculated using nonlinear regression in GraphPad Prism 8.0.

Statistical analysis was performed using a Wilcoxon matched-pairs signed rank test. Two-tailed p-values are reported.

### Neutralization of infectious SARS-CoV-2

Infectious SARS-CoV-2 isolate hCoV-19/USA/NY-NP-DOH1/2021 was isolated at the Aaron Diamond AIDS Center (Columbia University Medical Ctr) from nasopharyngeal swab and propagated for one passage in Vero E6 cells (ATCC). Infectious titer of the resulting virus was determined by an end-point dilution and cytopathic effect (CPE) assay on Vero-E6 cells as described previously^25^. The virus has since been deposited at BEI Resources (Cat#NR-55359). SARS-CoV-2 virus USA-WA1/2020 (WA1) obtained from BEI Resources (Cat# NR-52281) served as the control in experiments.

An end-point dilution microplate neutralization assay was performed to measure the neutralization activity of twenty patient convalescent plasma samples and twelve purified monoclonal antibodies. In brief, plasma samples were subjected to successive 5-fold dilutions starting from 1:100. Similarly, antibodies were serially diluted (5-fold dilutions) starting at 50 µg/ml. Triplicates of each dilution were incubated with SARS-CoV-2 at an MOI of 0.1 in EMEM with 7.5% inactivated fetal calf serum (FCS) for 1 hour at 37°C. Post incubation, the virus-antibody mixture was transferred onto a monolayer of Vero-E6 cells grown overnight. The cells were incubated with the mixture for ∼70 hours. Cytopathic effect (CPE) of viral infection was visually scored for each well in a blinded fashion by two independent observers. The results were then converted into percentage neutralization at a given sample dilution or antibody concentration, and the averages ± SEM were plotted using a five-parameter dose-response curve in GraphPad Prism v8.4.

### Growth dynamics

Growth dynamics of B.1.1.7 and B.1.526 were obtained through by downloading “metadata” from gisaid.org on June 6, 2021 for all 422,760 viruses sampled from the USA collected after January 1, 2021. This metadata has PANGO lineages^39^ already assigned to each genome sequence. Daily state-level frequencies (and frequencies for CUIMC) were extracted for plotting via 7-day sliding window averages of the prevalence of B.1.1.7 and B.1.526, calculated as the number of sequence-verified samples from each strain divided by the total number of positive samples with cycle threshold (Ct) values below 35, as this threshold value was used for sequencing. Separately, a multinomial logistic regression model was fit directly to the observation data consisting of individual genomes, their dates of sampling (independent variable *X* in days since January 1, 2021) and their categorical labels (dependent variable *Y*, “B.1.1.7”, “B.1.526” and “other”). This results in a 4-parameter model where both B.1.1.7 and B.1.526 have parameters specified for frequency at day 0 (January 1, 2021) and logistic growth rate. This model was fit to the data using the Classify package of Mathematica v12.2.

### Data availability

All genomes and associated metadata generated as a part of this study have been uploaded to GISAID (gisaid.org) and NCBI GenBank (BioProject Accession PRJNA751551). Biological materials (i.e. variant pseudoviruses) generated as a part of this study will be made available but may require execution of a materials transfer agreement.

### Code availability

Data processing and visualization was performed using publicly available software and packages, primarily RStudio v1.2.5033, GraphPad Prism v8.4, and iTOL (https://itol.embl.de/). The exact workflow used for phylogenetic (Fig. 2A) and phylogeographic analysis of public GISAID data (Fig. 4E-F) is available at https://github.com/blab/ncov-ny. Frequency dynamics were modeled using Mathematica in notebooks also available at https://github.com/blab/ncov-ny.

## Acknowledgements

Biospecimens utilized for this research were obtained from the Columbia University Biobank (CUB) with technical support from Viplan J. Mahadeva, Sebastian Fernando and Sylvia T. Parker-Jones. CUB is supported by the Irving Institute for Clinical and Translational Research (NCATS UL1TR001873). In particular, we thank Muredach Reilly, Eldad Hod, and the CUB COVID-19 Genomics Consortium (CCGC) for facilitating this effort. We are also grateful to Lihong Liu and Sho Iketani for technical support, and Alan Perelson for mathematical input. We gratefully acknowledge all the authors, the originating laboratories responsible for obtaining the specimens, and the submitting laboratories for generating the genetic sequence and metadata and sharing via the GISAID Initiative, on which part of the presented research is based. This work was in part funded by NIH/NIDA grant U01 DA053949 (A.-C.U, M.K.A.) and by support from Andrew & Peggy Cherng, Samuel Yin, Barbara Picower and the JBP Foundation, Brii Biosciences, Roger & David Wu, and the Bill and Melinda Gates Foundation. T.B. is a Pew Biomedical Scholar and is supported by NIH grant no. R35 GM119774-01. Funders and funding agencies had no role in study design, data collection and analysis, decision to publish, or preparation of the manuscript.

## Competing Interests

P.W., M.S.N., Y.H., and D.D.H. are inventors on a provisional patent application on monoclonal antibodies against SARS-CoV-2. D.D.H. is a member of the scientific advisory board of Brii Biosciences, which has provided a grant to Columbia University to support this and other studies on SARS-CoV-2. A.-C.U. and D.D.H. have received funding from Merck & Co. unrelated to this study.

## Author Contributions

**Conceptualization** – A.-C.U., D.D.H., M.K.A., H.M.; **Data curation** – M.K.A., H.M., J.E.Z., P.W., M.S.N., Z.S., T.B., A.G.-S., Y.H., A.L.K., M.T., A.-C.U.; **Formal analysis** – M.K.A., P.W., J.E.Z., T.B., A.G.-S.; **Funding acquisition** – A.-C.U., D.D.H., M.K.A.; **Investigation** – M.K.A., H.M., J.E.Z., P.W., M.S.N., A.L.K., M.T., T.B., Y.H.; **Methodology** – M.K.A., H.M., P.W., M.S.N., T.B., Y.H.; **Supervision** – A.-C.U., D.D.H.; **Visualization** – M.K.A., P.W., T.B.; **Writing – original draft** – A.-C.U., M.K.A., H.M., D.D.H.; **Writing – review and editing** – all authors

## Extended Data

**Extended Data Table 1.** Clinical characteristics of patients infected with SARS-CoV-2 based on viral genotype.

| Clinical Characteristic | E484K (n=170)<sup>1</sup> | Wildtype<sup>1</sup><br>(n=1,180) | P<sup>2</sup> | B.1.526-E484K<sup>1</sup><br>(n=232) | Non-VOI/VOC<br>lineages<sup>1</sup><br>(n=790) | P<sup>2</sup> |
| --- | --- | --- | --- | --- | --- | --- |
| **Demographics** |  |  |  |  |  |  |
| Male sex, n (%) | 80 (47.1) | 539 (45.7) | 0.806 | 97 (41.8) | 373 (47.3) | 0.159 |
| Age, years (median [IQR]) | 56 [36, 68] | 55 [33, 70] | 0.965<sup>3</sup> | 51 [32, 69] | 51 [30, 68] | 0.743<sup>3</sup> |
| Race and ethnicity, n (%) |  |  | 0.288 |  |  | 0.117 |
| Hispanic/Latino | 87 (51.2) | 548 (46.5) |  | 122 (52.6) | 357 (45.3) |  |
| Black | 11 (6.5) | 131 (11.1) |  | 27 (11.6) | 89 (11.3) |  |
| White | 24 (14.1) | 172 (14.6) |  | 23 (9.9) | 119 (15.1) |  |
| Other | 48 (28.2) | 328 (27.8) |  | 60 (25.9) | 223 (28.3) |  |
| Place of residence, n (%) |  |  | 0.008 |  |  | 0.002 |
| NYC | 151 (88.8) | 959 (81.3) |  | 204 (87.9) | 627 (79.4) |  |
| Yonkers | 7 (4.1) | 34 (2.9) |  | 9 (3.9) | 23 (2.9) |  |
| Outside NYC and Yonkers | 12 (7.1) | 187 (15.8) |  | 19 (8.2) | 140 (17.7) |  |
| **Comorbidities** |  |  |  |  |  |  |
| BMI, kg/m<sup>2</sup> (median [IQR]) | 28.9 [25.0, 33.1] | 27.1 [23.5, 31.3] | 0.01<sup>3</sup> | 28.6 [24.7, 33.1] | 26.6 [23.1, 30.5] | 0.001<sup>3</sup> |
| Hypertension, n (%) | 70 (41.7) | 444 (41.2) | 0.981 | 86 (38.4) | 291 (40.6) | 0.602 |
| Diabetes mellitus, n (%) | 51 (30.4) | 265 (24.6) | 0.134 | 63 (28.1) | 166 (23.2) | 0.157 |
| Chronic kidney disease, n (%) | 21 (12.5) | 126 (11.7) | 0.864 | 18 (8.0) | 93 (13.0) | 0.059 |
| Coronary artery disease, n (%) | 13 (7.7) | 108 (10.0) | 0.428 | 16 (7.1) | 72 (10.1) | 0.240 |
| Solid organ transplant, n (%) | 7 (4.2) | 42 (3.9) | 1.00 | 7 (3.1) | 41 (5.7) | 0.171 |
| **Cycle threshold value (mean (SD))<sup>4</sup>** | 29.49 (5.64) | 30.71 (5.66) | 0.013 | 27.65 (4.88) | 28.81 (4.93) | 0.015 |
| **Severity of care and outcomes** |  |  |  |  |  |  |
| Highest level of care, n (%) |  |  | 0.175 |  |  | 0.037 |
| Admitted | 50 (29.6) | 417 (35.4) |  | 81 (35.1) | 228 (29.0) |  |
| Emergency Department | 64 (37.9) | 339 (28.8) |  | 83 (35.9) | 243 (31.0) |  |
| ICU | 13 (7.7) | 86 (7.3) |  | 15 (6.5) | 54 (6.9) |  |
| Outpatient | 42 (24.9) | 335 (28.4) |  | 52 (22.5) | 259 (33.0) |  |
| Supplemental oxygen, n (%) | 48 (85.7) | 348 (77.5) | 0.217 | 67 (81.7) | 187 (73.9) | 0.199 |
| Outcome, n (%) |  |  | 0.454 |  |  | 0.565 |
| Deceased or discharged to hospice | 8 (4.7) | 87 (7.4) |  | 10 (4.4) | 45 (5.7) |  |
| Further care at external facility | 16 (9.5) | 85 (7.2) |  | 16 (7.0) | 44 (5.6) |  |
| Discharged to home | 145 (85.8) | 1003 (85.4) |  | 202 (88.2) | 693 (88.5) |  |

<sup>1</sup> Wildtype isolates are defined as those without E484K or N501Y mutations. For comparisons based on lineage, B.1.526-E484K was compared with non-variant of interest (VOI) and non-variant of concern (VOC) lineages (i.e. B.1.526.1, B.1.526.2, B.1.1.7, P.1, P.2, and B.1.351 were excluded). Comparisons were made using the first genotyped sample per patient to exclude multiple measures.
<sup>2</sup> T-tests (two-sided) were performed for continuous variables and chi-squared tests for categorical variables, unless otherwise indicated as below
<sup>3</sup> Due to non-normal distribution, Kruskal-Wallis non-parametric test was used
<sup>4</sup> Cycle threshold value as determined through our rapid qPCR-based screening assay on heat-inactivated nasopharyngeal swab samples

**Extended Data Figure 1.**
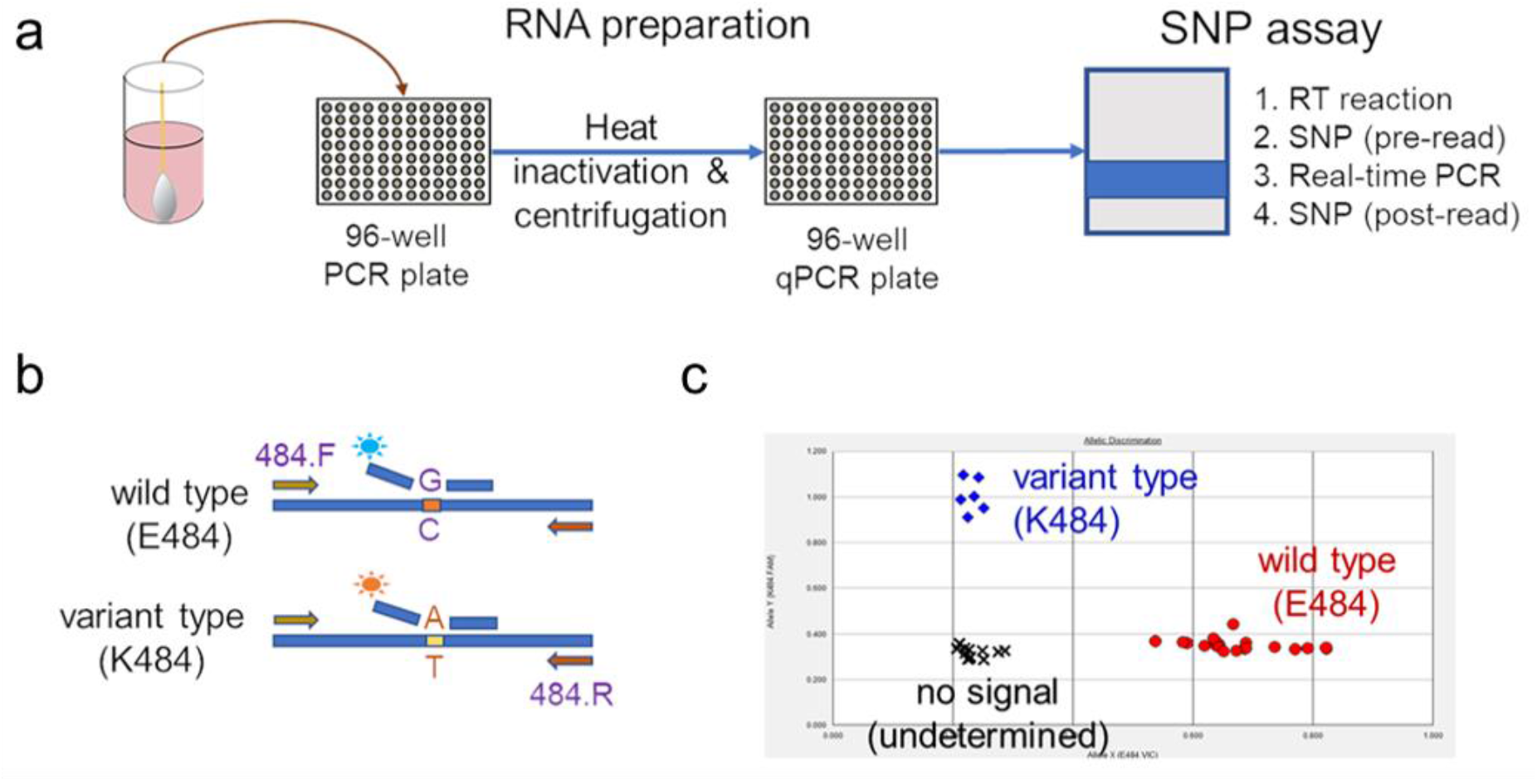
Rapid PCR-based screening assay protocol to identify samples harboring key substitutions. (a) Viral RNA is prepared by heat inactivation and centrifugation. The supernatant is then used for the SNP assay, which entails four steps: the reverse transcription (RT) reaction, pre-PCR reading of the plate to assess background fluorescence (SNP pre-read), real-time PCR, and post-PCR reading of the plate to measure fluorescence (SNP post-read). The runtime for this entire protocol is approximately two hours. (b) Genotype at targeted sites in COVID-19 viral RNA can be determined with two MGB probes, one for wild type (conjugated with VIC) and the other for variant type (conjugated with FAM). (c) Example signals for the variant type (K484; blue), the wild type (E484; red) and samples with no signal (black) are shown.

**Extended Data Figure 2.**
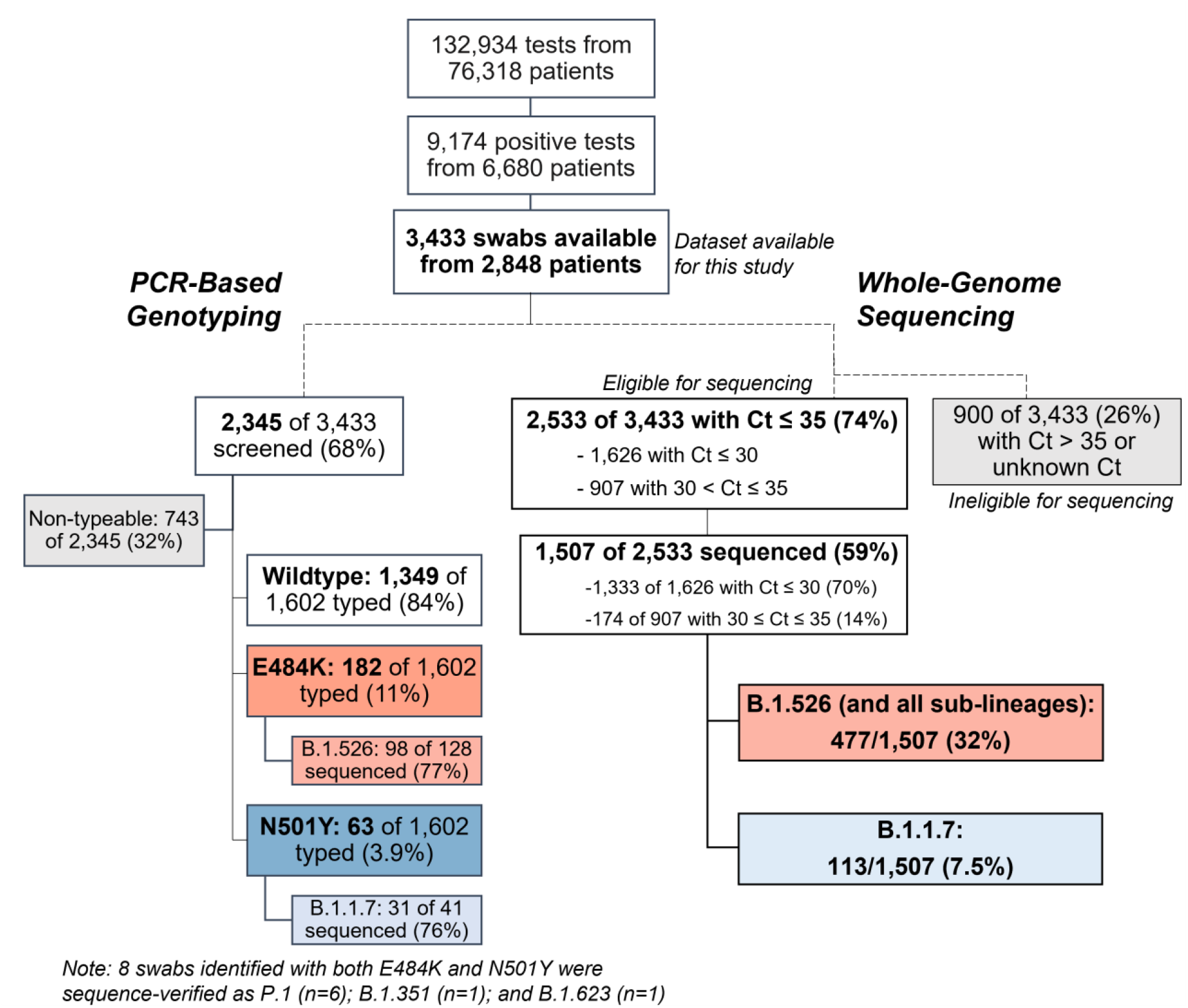
Flowchart for SARS-CoV-2-positive nasopharyngeal swabs included in this study. (**top**) During the study period of November 1, 2020 to May 1, 2021, 6,680 patients tested positive for SARS-CoV-2 at our hospital center and affiliated hospitals. From these 9,174 positive nasopharyngeal swabs, 3,433 swabs were stored as part of the Columbia University Biobank COVID-19 sample repository and available for this study. (**left**) PCR-based genotyping assays for E484K and N501Y (see Extended Data Fig. 1) were performed on 2,345 samples. We identified a significant proportion of samples with E484K (11%), later confirmed through sequencing to primarily fall within the B.1.526 lineage, and a number of samples with N501Y (3.9%), primarily within the B.1.1.7 lineage. (**right**) We performed whole-genome sequencing on 1,507 samples. Of these, 32% belonged to B.1.526 and the sublineages B.1.526.1 and B.1.526.2, while B.1.1.7 constituted a much smaller proportion of samples at our center (7.5%).

**Extended Data Figure 3.**
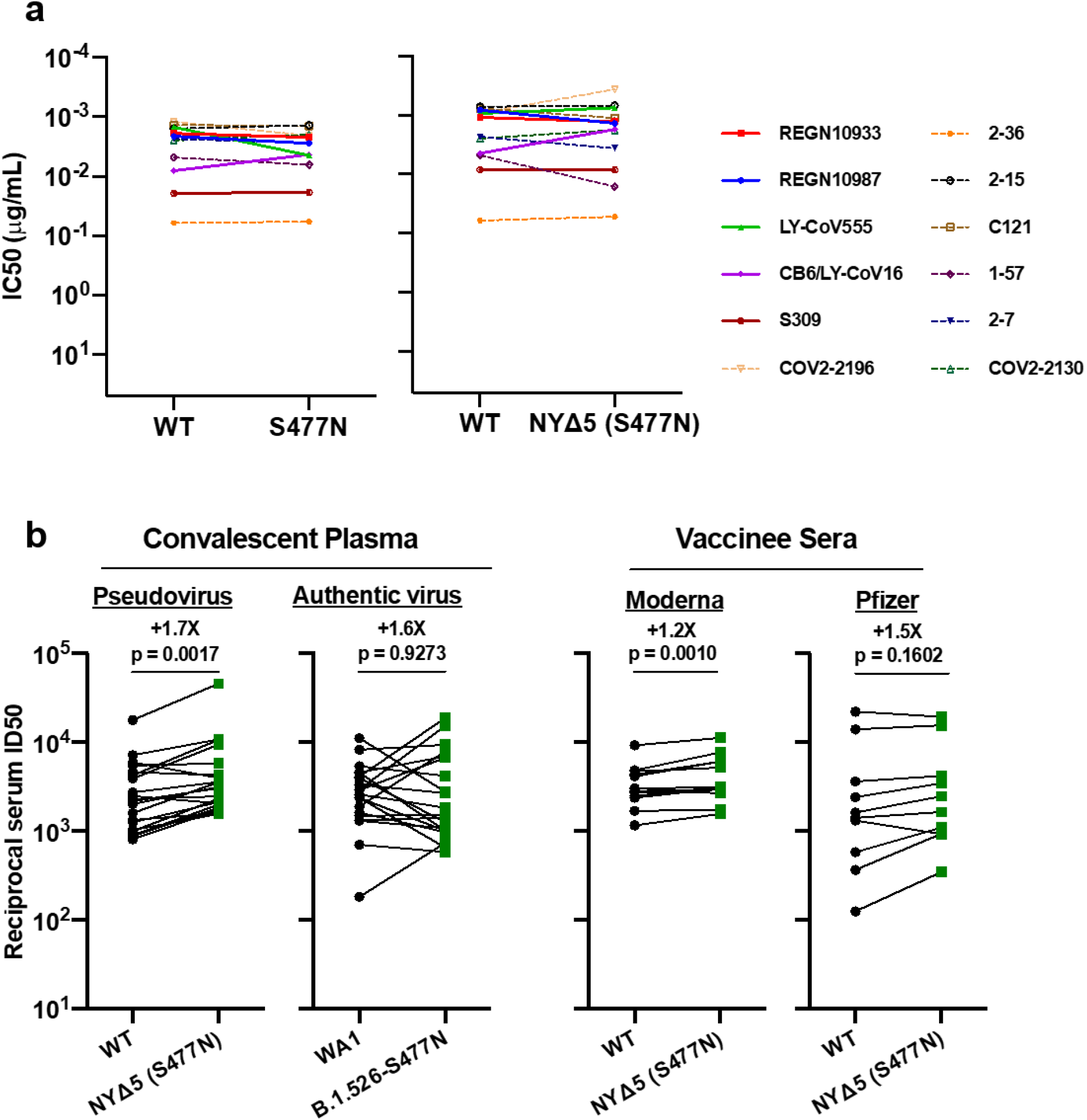
Neutralization studies of B.1.526-S477N. **(a)** Neutralizing activities of 12 monoclonal antibodies against pseudoviruses containing S477N alone or all five signature B.1.526-S477N mutations (L5F, T95I, D253G, A701V, and S477N), termed NYΔ5(S477N). Antibodies with emergency use authorization are shown in bold solid lines. Data are represented as mean ± SEM. of technical triplicates and represent one of two independent experiments. **(b)** Neutralizing activities of convalescent plasma (n=20) against NYΔ5(S477N) as well as against the authentic B.1.526 virus with S477N, and neutralizing activities of vaccinee sera (n=22) against the NYΔ5(S477N) pseudovirus, compared to wildtype counterparts. Statistical comparisons were made using the Wilcoxon matched-pairs signed rank test; two-tailed p-values are reported.

**Extended Data Figure 4.**
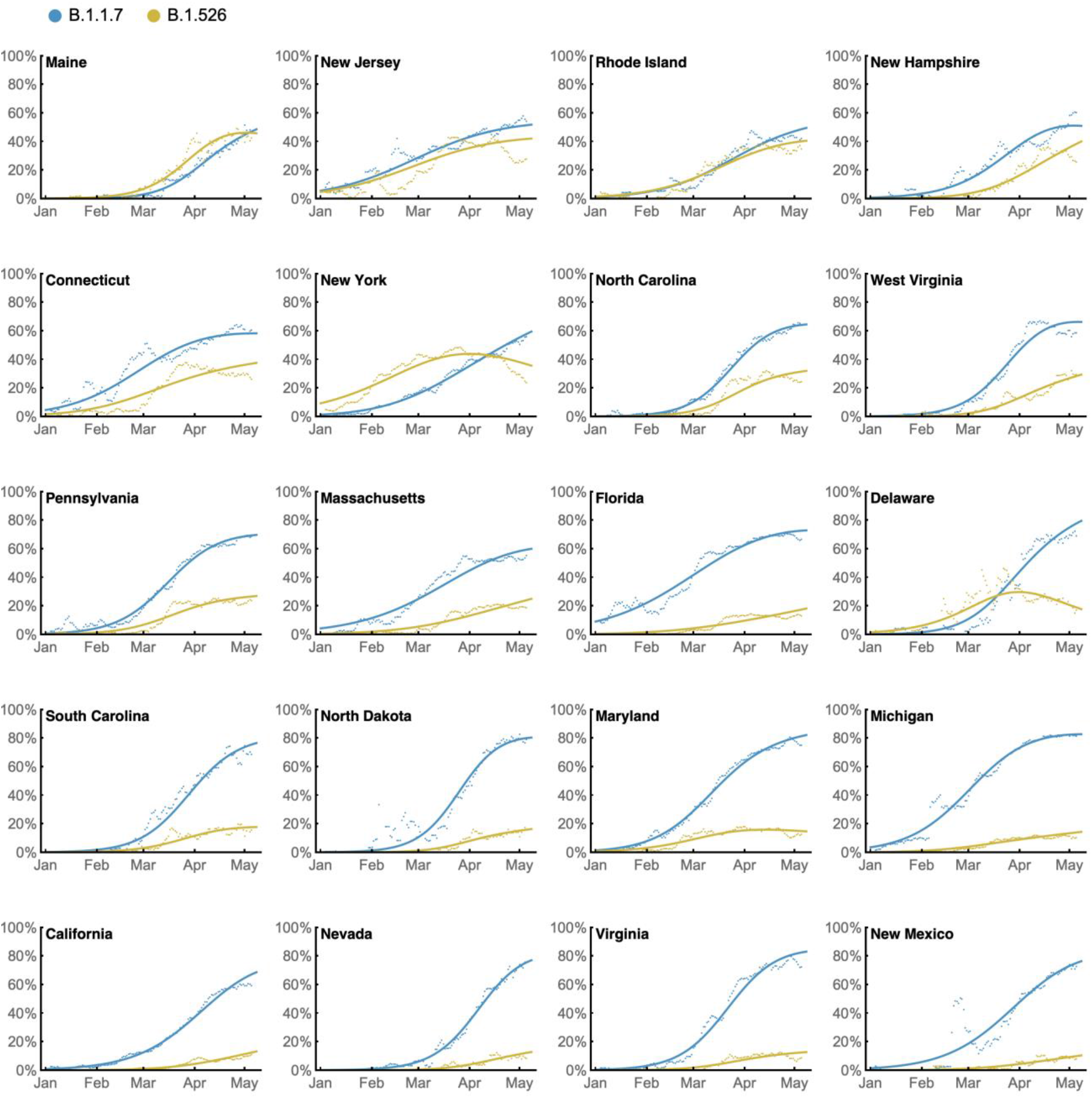
State-level growth dynamics of B.1.526 and B.1.1.7. Daily state-level frequencies of B.1.526 (in yellow) and B.1.1.7 (in blue), based on GISAID data downloaded on June 6, 2021, were used to plot 7-day sliding window averages of the prevalence of each lineage (shown as dots in the figure). A 4-parameter multinomial logistic regression model was fit directly to the observation data, in which both B.1.1.7 and B.1.526 have parameters specified for frequency at day 0 (January 1, 2021) and logistic growth rate (shown as lines in the figure). States are ordered according to frequency of B.1.526 at the final timepoint of May 8, 2021.

## Notes

### Funding Statement

Biospecimens utilized for this research were obtained from the Columbia University Biobank (CUB), supported by the Irving Institute for Clinical and Translational Research, home to Columbia University Clinical and Translational Science Award (CTSA) funded through Grant Number UL1TR001873. This work was in part funded by NIH/NIDA grant U01 DA053949 (ACU, MKA) and by support from Andrew & Peggy Cherng, Samuel Yin, Barbara Picower and the JBP Foundation, Brii Biosciences, Roger & David Wu, and the Bill and Melinda Gates Foundation.

### Author Declarations

This study was reviewed and approved by the Columbia University Institutional Review Board (protocol number AAAT0123).

### Summary of Updates

The manuscript was updated to reflect the emergence and spread of B.1.526 in context with the recent and current landscape of SARS-CoV-2 variants.

## References

1 Rambaut, A. et al. Preliminary genomic characterisation of an emergent SARSCoV-2 lineage in the UK defined by a novel set of spike mutations., <https://virological.org/t/preliminary-genomic-characterisation-of-an-emergent-sars-cov-2-lineage-in-the-uk-defined-by-a-novel-set-of-spike-mutations/563> (2020).

2 Tegally, H. et al. Emergence and rapid spread of a new severe acute respiratory syndrome-related coronavirus 2 (SARS-CoV-2) lineage with multiple spike mutations in South Africa. medRxiv, 2020.2012.2021.20248640, doi:10.1101/2020.12.21.20248640 (2020).

3 Faria, N. R. et al. Genomic characterisation of an emergent SARS-CoV-2 lineage in Manaus: preliminary findings. (2021).

4 Wang, P. et al. Antibody Resistance of SARS-CoV-2 Variants B.1.351 and B.1.1.7. Nature, doi:10.1038/s41586-021-03398-2 (2021).

5 Duchene, S. et al. Temporal signal and the phylodynamic threshold of SARS-CoV-2. Virus Evol 6, veaa061, doi:10.1093/ve/veaa061 (2020).

6 Cherian, S. et al. Convergent evolution of SARS-CoV-2 spike mutations, L452R, E484Q and P681R, in the second wave of COVID-19 in Maharashtra, India. bioRxiv, 2021.2004.2022.440932, doi:10.1101/2021.04.22.440932 (2021).

7 Iacobucci, G. Covid-19: New UK variant may be linked to increased death rate, early data indicate. BMJ 372, n230, doi:10.1136/bmj.n230 (2021).

8 Volz, E. et al. Transmission of SARS-CoV-2 Lineage B.1.1.7 in England: Insights from linking epidemiological and genetic data. medRxiv, 2020.2012.2030.20249034, doi:10.1101/2020.12.30.20249034 (2021).

9 Washington, N. L. et al. Genomic epidemiology identifies emergence and rapid transmission of SARS-CoV-2 B.1.1.7 in the United States. medRxiv, doi:10.1101/2021.02.06.21251159 (2021).

10 Greaney, A. J. et al. Comprehensive mapping of mutations in the SARS-CoV-2 receptor-binding domain that affect recognition by polyclonal human plasma antibodies. Cell Host Microbe 29, 463–476 e466, doi:10.1016/j.chom.2021.02.003 (2021).

11 Thorne, L. G. et al. Evolution of enhanced innate immune evasion by the SARS-CoV-2 B.1.1.7 UK variant. bioRxiv, 2021.2006.2006.446826, doi:10.1101/2021.06.06.446826 (2021).

12 Faria, N. R. et al. Genomics and epidemiology of the P.1 SARS-CoV-2 lineage in Manaus, Brazil. Science, doi:10.1126/science.abh2644 (2021).

13 Sabino, E. C. et al. Resurgence of COVID-19 in Manaus, Brazil, despite high seroprevalence. Lancet 397, 452–455, doi:10.1016/S0140-6736(21)00183-5 (2021).

14 Zucman, N., Uhel, F., Descamps, D., Roux, D. & Ricard, J. D. Severe reinfection with South African SARS-CoV-2 variant 501Y.V2: A case report. Clin Infect Dis, doi:10.1093/cid/ciab129 (2021).

15 Nonaka, C. K. V. et al. Genomic Evidence of SARS-CoV-2 Reinfection Involving E484K Spike Mutation, Brazil. Emerg Infect Dis 27, doi:10.3201/eid2705.210191 (2021).

16 Callaway, E. & Mallapaty, S. Novavax offers first evidence that COVID vaccines protect people against variants. Nature 590, 17, doi:10.1038/d41586-021-00268-9 (2021).

17 Madhi, S. A. et al. Efficacy of the ChAdOx1 nCoV-19 Covid-19 Vaccine against the B.1.351 Variant. N Engl J Med, doi:10.1056/NEJMoa2102214 (2021).

18 Wang, P. et al. Increased Resistance of SARS-CoV-2 Variant P.1 to Antibody Neutralization. bioRxiv, doi:10.1101/2021.03.01.433466 (2021).

19 West, A. P., Barnes, C. O., Yang, Z. & Bjorkman, P. J. SARS-CoV-2 lineage B.1.526 emerging in the New York region detected by software utility created to query the spike mutational landscape. bioRxiv, 2021.2002.2014.431043, doi:10.1101/2021.02.14.431043 (2021).

20 WHO. Tracking SARS-CoV-2 Variants, <https://www.who.int/en/activities/tracking-SARS-CoV-2-variants/> (2021).

21 Goudsmit, J., De Ronde, A., Ho, D. D. & Perelson, A. S. Human immunodeficiency virus fitness in vivo: calculations based on a single zidovudine resistance mutation at codon 215 of reverse transcriptase. J Virol 70, 5662–5664, doi:10.1128/JVI.70.8.5662-5664.1996 (1996).

22 Ali, S. T. et al. Serial interval of SARS-CoV-2 was shortened over time by nonpharmaceutical interventions. Science 369, 1106–1109, doi:10.1126/science.abc9004 (2020).

23 CDC. SARS-CoV-2 Variant Classifications and Definitions, <SARS-CoV-2 Variant Classifications and Definitions> (2021).

24 Cerutti, G. et al. Potent SARS-CoV-2 Neutralizing Antibodies Directed Against Spike N-Terminal Domain Target a Single Supersite. bioRxiv, 2021.2001.2010.426120, doi:10.1101/2021.01.10.426120 (2021).

25 Liu, L. et al. Potent neutralizing antibodies against multiple epitopes on SARS-CoV-2 spike. Nature 584, 450–456, doi:10.1038/s41586-020-2571-7 (2020).

26 Health, N. Y. C. D. o. COVID-19: Data, <https://www1.nyc.gov/site/doh/covid/covid-19-data-trends.page#antibody> (2021).

27 Lasek-Nesselquist, E., Lapierre, P., Schneider, E., George, K. S. & Pata, J. The localized rise of a B.1.526 SARS-CoV-2 variant containing an E484K mutation in New York State. medRxiv, 2021.2002.2026.21251868, doi:10.1101/2021.02.26.21251868 (2021).

28 Alaa Abdel Latif, K. G., Julia L. Mullen, Emily Haag, Ginger Tsueng, Nate Matteson, Mark Zeller, Chunlei Wu, Kristian G. Andersen, Andrew I. Su, Laura D. Hughes, and the Center for Viral Systems. B.1.526 Lineage Report., <https://outbreak.info/situation-reports/S-E484K>> (2021).

29 Smyrlaki, I. et al. Massive and rapid COVID-19 testing is feasible by extraction-free SARS-CoV-2 RT-PCR. Nat Commun 11, 4812, doi:10.1038/s41467-020-18611-5 (2020).

30 Quick, J. Artic Protocol, <https://www.protocols.io/view/ncov-2019-sequencing-protocol-v3-locost-bh42j8ye> (2021).

31 Freed, N., Vlkova, M., Faisal, M. B. & Silander, O. Rapid and inexpensive whole-genome sequencing of SARS-CoV2 using 1200 bp tiled amplicons and Oxford Nanopore rapid barcoding. bioRxiv, doi:10.1101/2020.05.28.122648 (2020).

32 Hadfield, J. et al. Nextstrain: real-time tracking of pathogen evolution. Bioinformatics 34, 4121–4123, doi:10.1093/bioinformatics/bty407 (2018).

33 Minh, B. Q. et al. IQ-TREE 2: New Models and Efficient Methods for Phylogenetic Inference in the Genomic Era. Mol Biol Evol 37, 1530–1534, doi:10.1093/molbev/msaa015 (2020).

34 Sagulenko, P., Puller, V. & Neher, R. A. TreeTime: Maximum-likelihood phylodynamic analysis. Virus Evol 4, vex042, doi:10.1093/ve/vex042 (2018).

35 Shu, Y. & McCauley, J. GISAID: Global initiative on sharing all influenza data - from vision to reality. Euro Surveill 22, doi:10.2807/1560-7917.ES.2017.22.13.30494 (2017).

36 Pinto, D. et al. Cross-neutralization of SARS-CoV-2 by a human monoclonal SARS-CoV antibody. Nature 583, 290–295, doi:10.1038/s41586-020-2349-y (2020).

37 Zost, S. J. et al. Rapid isolation and profiling of a diverse panel of human monoclonal antibodies targeting the SARS-CoV-2 spike protein. Nat Med 26, 1422–1427, doi:10.1038/s41591-020-0998-x (2020).

38 Robbiani, D. F. et al. Convergent antibody responses to SARS-CoV-2 in convalescent individuals. Nature 584, 437–442, doi:10.1038/s41586-020-2456-9 (2020).

39 Rambaut, A. et al. A dynamic nomenclature proposal for SARS-CoV-2 lineages to assist genomic epidemiology. Nat Microbiol 5, 1403–1407, doi:10.1038/s41564-020-0770-5 (2020).

