## Supplementary Table 1 for "Emergence and Expansion of the SARS-CoV-2 Variant B.1.526 Identified in New York" (from another version of this preprint: version 2, 2021-04-15)

We gratefully acknowledge the following Authors from the Originating laboratories responsible for obtaining the specimens, as well as the Submitting laboratories where the genome data were generated and shared via GISAID, on which this research is based.

All Submitters of data may be contacted directly via www.gisaid.org

Authors are sorted alphabetically.

| Accession ID | Originating Laboratory | Submitting Laboratory | Authors |
|---|---|---|---|
| EPI_ISL_1005532 | USC Clinical Lab | Los Angeles County PHL | P. Hemarajata et al. |
| EPI_ISL_1009653, EPI_ISL_1009654 | DPHL | Delaware Public Health Lab | Gregory Hovan |
| EPI_ISL_1009769 | Massachusetts State Public Health Laboratory | Massachusetts State Public Health Laboratory | Andrew Lang, Timelia Fink, Glen Gallagher, Sandra Smole |
| EPI_ISL_1015695, EPI_ISL_1015696, EPI_ISL_1015697, EPI_ISL_1015698, EPI_ISL_1015702, EPI_ISL_1015703, EPI_ISL_1015704, EPI_ISL_1016111, EPI_ISL_1016116 | Columbia University Irving Medical Center | Wadsworth Center, New York State Department of Health | Kirsten St. George, Daryl M. Lamson, Alexis Russel, Matthew Shudt, Melissa A Leisner, Jonathan Plitnick, Navjot Singh, John Kelly, Erasmus Schneider, Erica Lasek-Nesselquist |
| EPI_ISL_1016158 | NORTHWELL HEALTH LABORATORIES | Wadsworth Center, New York State Department of Health | Kirsten St. George, Daryl M. Lamson, Alexis Russel, Matthew Shudt, Melissa A Leisner, Jonathan Plitnick, Navjot Singh, John Kelly, Erasmus Schneider, Erica Lasek-Nesselquist |
| EPI_ISL_1016174, EPI_ISL_1016177, EPI_ISL_1016179, EPI_ISL_1016180, EPI_ISL_1016181, EPI_ISL_1016182, EPI_ISL_1016183, EPI_ISL_1016185, EPI_ISL_1016186, EPI_ISL_1016187, EPI_ISL_1016191, EPI_ISL_1016197, EPI_ISL_1016199, EPI_ISL_1016201, EPI_ISL_1016206, EPI_ISL_1016209, EPI_ISL_1016213, EPI_ISL_1016218 | | | |
| see above | MONTEFIORE MEDICAL CENTER LABORATORIES | Wadsworth Center, New York State Department of Health | Kirsten St. George, Daryl M. Lamson, Alexis Russel, Matthew Shudt, Melissa A Leisner, Jonathan Plitnick, Navjot Singh, John Kelly, Erasmus Schneider, Erica Lasek-Nesselquist |
| EPI_ISL_1016226, EPI_ISL_1016228, EPI_ISL_1016233, EPI_ISL_1016242, EPI_ISL_1016244 | New York Presbyterian Hospital | Wadsworth Center, New York State Department of Health | Kirsten St. George, Daryl M. Lamson, Alexis Russel, Matthew Shudt, Melissa A Leisner, Jonathan Plitnick, Navjot Singh, John Kelly, Erasmus Schneider, Erica Lasek-Nesselquist |
| EPI_ISL_1016260, EPI_ISL_1016270 | Columbia University Irving Medical Center | Wadsworth Center, New York State Department of Health | Kirsten St. George, Daryl M. Lamson, Alexis Russel, Matthew Shudt, Melissa A Leisner, Jonathan Plitnick, Navjot Singh, John Kelly, Erasmus Schneider, Erica Lasek-Nesselquist |
| EPI_ISL_1016271 | New York Presbyterian Hospital | Wadsworth Center, New York State Department of Health | Kirsten St. George, Daryl M. Lamson, Alexis Russel, Matthew Shudt, Melissa A Leisner, Jonathan Plitnick, Navjot Singh, John Kelly, Erasmus Schneider, Erica Lasek-Nesselquist |
| EPI_ISL_1016344, EPI_ISL_1016345 | WESTCHESTER MEDICAL CENTER | Wadsworth Center, New York State Department of Health | Kirsten St. George, Daryl M. Lamson, Alexis Russel, Matthew Shudt, Melissa A Leisner, Jonathan Plitnick, Navjot Singh, John Kelly, Erasmus Schneider, Erica Lasek-Nesselquist |
| EPI_ISL_1017952, EPI_ISL_1017956, EPI_ISL_1017963 | Wyoming Public Health Laboratory | Wyoming Public Health Laboratory | Noah Hull, Taylor Fearing, Lynette Gumbleton, Channing Weber, Ashley Norberg, Bailey Bowcutt, and Wanda Manley |
| EPI_ISL_1017970 | DOHMH PHL | New York City Public Health Laboratory | Jade Wang, et al. |
| EPI_ISL_1017973 | DOHMH Riverside | New York City Public Health Laboratory | Jade Wang, et al. |
| EPI_ISL_1017978 | DOHMH Chelsea | New York City Public Health Laboratory | Jade Wang, et al. |
| EPI_ISL_1017982 | DOHMH Central Harlem | New York City Public Health Laboratory | Jade Wang, et al. |
| EPI_ISL_1017990 | Department of Homeless Services | New York City Public Health Laboratory | Jade Wang, et al. |
| EPI_ISL_1018003, EPI_ISL_1018004 | DOHMH PHL | New York City Public Health Laboratory | Jade Wang, et al. |
| EPI_ISL_1018012, EPI_ISL_1018013 | DOHMH Jamaica | New York City Public Health Laboratory | Jade Wang, et al. |
| EPI_ISL_1018025 | DOHMH Morrisania | New York City Public Health Laboratory | Jade Wang, et al. |
| EPI_ISL_1018028, EPI_ISL_1018029 | DOHMH Corona | New York City Public Health Laboratory | Jade Wang, et al. |
| EPI_ISL_1018033, EPI_ISL_1018034, EPI_ISL_1018037, EPI_ISL_1018047 | OCME Office Of Chief Medical Examiner | New York City Public Health Laboratory | Jade Wang, et al. |
| EPI_ISL_1020489, EPI_ISL_1020490, EPI_ISL_1020508, EPI_ISL_1020509, EPI_ISL_1020514, EPI_ISL_1020515 | Columbia University Irving Medical Center | Wadsworth Center, New York State Department of Health | Kirsten St. George, Daryl M. Lamson, Alexis Russel, Matthew Shudt, Melissa A Leisner, Jonathan Plitnick, Navjot Singh, John Kelly, Erasmus Schneider, Erica Lasek-Nesselquist |
| EPI_ISL_1020948, EPI_ISL_1020964, EPI_ISL_1020994, EPI_ISL_1021057, EPI_ISL_1021524, EPI_ISL_1022027, EPI_ISL_1031584 | Laboratory Corporation of America | Respiratory Viruses Branch, Division of Viral Diseases, Centers for Disease Control and Prevention | Peter W. Cook, Dakota Howard, Dhwani Batra, Ben L. Rambo-Martin, Clinton R. Paden, Suxiang Tong, Duncan MacCannell |
| EPI_ISL_1036298 | Johns Hopkins Hospital Department of Pathology | Johns Hopkins Hospital Department of Pathology | C. Paul Morris, Chun Huai Luo, Adannaya Amadi, Matthew Schwartz, Nicholas Gallagher, Heba H. Mostafa |
| EPI_ISL_1037134, EPI_ISL_1037485, EPI_ISL_1037510, EPI_ISL_1037670, EPI_ISL_1037971 | Laboratory Corporation of America | Respiratory Viruses Branch, Division of Viral Diseases, Centers for Disease Control and Prevention | Peter W. Cook, Dakota Howard, Dhwani Batra, Ben L. Rambo-Martin, Clinton R. Paden, Suxiang Tong, Duncan MacCannell |
| EPI_ISL_1038998 | Yale Pathology Lab | Grubaugh Lab - Yale School of Public Health | Joseph Fauver, Tara Alpert, Anderson Brito, Mallery Breban, Anne Wyllie, Chantal Vogels, Mary Petrone, Annie Watkins, Chaney Kalinich, Isabel Ott, Nathan Grubaugh |
| EPI_ISL_1040855, EPI_ISL_1040857, EPI_ISL_1040858, EPI_ISL_1040861, EPI_ISL_1040865, EPI_ISL_1040868, EPI_ISL_1040875, EPI_ISL_1040878, EPI_ISL_1040884, EPI_ISL_1040895, EPI_ISL_1040898, EPI_ISL_1040899, EPI_ISL_1040900, EPI_ISL_1040901 | | | |
| see above | NYU Langone Health | Departments of Pathology and Medicine, New York University School of Medicine | Adriana Heguy, Dacia Dimartino, Emily Guzman, Christian Marier, Peter Meyn, Sitharam Ramaswami, Gael Westby, Paul Zappile, Yutong Zhang, Paolo Cotzia, Guiqing Wang |
| EPI_ISL_1041215, EPI_ISL_1041222, EPI_ISL_1041223, EPI_ISL_1041226, EPI_ISL_1041227, EPI_ISL_1041228, EPI_ISL_1041233, EPI_ISL_1041234, EPI_ISL_1041238, EPI_ISL_1041239, EPI_ISL_1041240, EPI_ISL_1041241, EPI_ISL_1041242, EPI_ISL_1041245, EPI_ISL_1041247, EPI_ISL_1041251, EPI_ISL_1041262, EPI_ISL_1041266, EPI_ISL_1041267, EPI_ISL_1041268, EPI_ISL_1041269, EPI_ISL_1041273, EPI_ISL_1041274, EPI_ISL_1041283, EPI_ISL_1041284, EPI_ISL_1041291, EPI_ISL_1041292, EPI_ISL_1041296, EPI_ISL_1041297, EPI_ISL_1041300, EPI_ISL_1041306, EPI_ISL_1041308, EPI_ISL_1041315, EPI_ISL_1041318, EPI_ISL_1041333, EPI_ISL_1041336, EPI_ISL_1041337, EPI_ISL_1041338, EPI_ISL_1041340, EPI_ISL_1041341, EPI_ISL_1041353, EPI_ISL_1041361, EPI_ISL_1041364, EPI_ISL_1041369, EPI_ISL_1041372, EPI_ISL_1041380, EPI_ISL_1041383, EPI_ISL_1041386, EPI_ISL_1041388, EPI_ISL_1041397, EPI_ISL_1041398, EPI_ISL_1041400, EPI_ISL_1041403, EPI_ISL_1041405, EPI_ISL_1041414, EPI_ISL_1041415, EPI_ISL_1041416, EPI_ISL_1041421, EPI_ISL_1041422, EPI_ISL_1041424, EPI_ISL_1041426, EPI_ISL_1041443, EPI_ISL_1041444, EPI_ISL_1041454, EPI_ISL_1041458, EPI_ISL_1041461, EPI_ISL_1041467, EPI_ISL_1041475, EPI_ISL_1041476, EPI_ISL_1041480, EPI_ISL_1041483, EPI_ISL_1041484, EPI_ISL_1041487, EPI_ISL_1041488, EPI_ISL_1041493, EPI_ISL_1041495, EPI_ISL_1041499, EPI_ISL_1041502, EPI_ISL_1041503, EPI_ISL_1041504, EPI_ISL_1041505, EPI_ISL_1041507, EPI_ISL_1041512, EPI_ISL_1041513, EPI_ISL_1041514, EPI_ISL_1041516, EPI_ISL_1041519, EPI_ISL_1041521, EPI_ISL_1041523, EPI_ISL_1041525, EPI_ISL_1041526, EPI_ISL_1041538, EPI_ISL_1041544, EPI_ISL_1041545, EPI_ISL_1041553, EPI_ISL_1041554, EPI_ISL_1041558, EPI_ISL_1041559, EPI_ISL_1041560, EPI_ISL_1041561, EPI_ISL_1041563, EPI_ISL_1041569, EPI_ISL_1041574, EPI_ISL_1041580, EPI_ISL_1041581, EPI_ISL_1041583, EPI_ISL_1041584, EPI_ISL_1041585, EPI_ISL_1041586, EPI_ISL_1041587, EPI_ISL_1041594, EPI_ISL_1041595, EPI_ISL_1041596, EPI_ISL_1041597, EPI_ISL_1041599, EPI_ISL_1041600, EPI_ISL_1041604, EPI_ISL_1041605, EPI_ISL_1041608, EPI_ISL_1041610, EPI_ISL_1041611, EPI_ISL_1041614, EPI_ISL_1041616, EPI_ISL_1041625, EPI_ISL_1041634, EPI_ISL_1041635, EPI_ISL_1041641, EPI_ISL_1041642, EPI_ISL_1041645, EPI_ISL_1041649, EPI_ISL_1041650, EPI_ISL_1041652, EPI_ISL_1041657, EPI_ISL_1041661, EPI_ISL_1041667, EPI_ISL_1041668, EPI_ISL_1041670, EPI_ISL_1041672, EPI_ISL_1041673, EPI_ISL_1041675, EPI_ISL_1041676, EPI_ISL_1041682, EPI_ISL_1041686, EPI_ISL_1041689, EPI_ISL_1041695, EPI_ISL_1041698, EPI_ISL_1041701, EPI_ISL_1041704, EPI_ISL_1041710, EPI_ISL_1041714, EPI_ISL_1041715, EPI_ISL_1041718, EPI_ISL_1041727, | | | |

EPI_ISL_1041728, EPI_ISL_1041731, EPI_ISL_1041733, EPI_ISL_1041734, EPI_ISL_1041744, EPI_ISL_1041745, EPI_ISL_1041748, EPI_ISL_1041750, EPI_ISL_1041760, EPI_ISL_1041761, EPI_ISL_1041765, EPI_ISL_1041781, EPI_ISL_1041808, EPI_ISL_1041809, EPI_ISL_1041821, EPI_ISL_1041826, EPI_ISL_1041827, EPI_ISL_1041829, EPI_ISL_1041830, EPI_ISL_1041831, EPI_ISL_1041832, EPI_ISL_1041837, EPI_ISL_1041839, EPI_ISL_1041840, EPI_ISL_1041841, EPI_ISL_1041844, EPI_ISL_1041849, EPI_ISL_1041857, EPI_ISL_1041858, EPI_ISL_1041859, EPI_ISL_1041861, EPI_ISL_1041867, EPI_ISL_1041868, EPI_ISL_1041869, EPI_ISL_1041871, EPI_ISL_1041872, EPI_ISL_1041873, EPI_ISL_1041874, EPI_ISL_1041876, EPI_ISL_1041884, EPI_ISL_1041885, EPI_ISL_1041886, EPI_ISL_1041888, EPI_ISL_1041899, EPI_ISL_1041900, EPI_ISL_1041906, EPI_ISL_1041911, EPI_ISL_1041912, EPI_ISL_1041913, EPI_ISL_1041927, EPI_ISL_1041928, EPI_ISL_1041929, EPI_ISL_1041930

| | | | |
|---|---|---|---|
| see above | Pandemic Response Lab - NYC | Pandemic Response Lab, R&D | Henry Lee, Michael Hammerling, Melissa Hopkins, Cybill del Castillo, William Ward, Pradeep Bugga, Haiping Hao, Jon Laurent |
| EPI_ISL_1046773 | SARS-CoV-2 testing team, National Institute of Infectious Diseases | Pathogen Genomics Center, National Institute of Infectious Diseases | Tsuyoshi Sekizuka, Kentaro Itokawa, Rina Tanaka, Masanori Hashino, Tadashi Kikuchi, Lucky Ronald Runtuwene, Shigeru Kusagawa, Makoto Kuroda |
| EPI_ISL_1050032 | NYU Langone Health | Departments of Pathology and Medicine, New York University School of Medicine | Adriana Heguy, Dacia Dimartino, Emily Guzman, Christian Marier, Peter Meyn, Sitharam Ramaswami, Gael Westby, Paul Zappile, Yutong Zhang, Paolo Cotzia, Guiqing Wang |
| EPI_ISL_1058874 | Michigan Department of Health and Human Services, Bureau of Laboratories | Michigan Department of Health and Human Services, Bureau of Laboratories | Blankenship HM, Riner D, Soehnlen MK |
| EPI_ISL_1059101 | University of Michigan Clinical Microbiology Laboratory | Lauring Lab, University of Michigan, Department of Microbiology and Immunology | Valesano |
| EPI_ISL_1060772, EPI_ISL_1060780, EPI_ISL_1060782, EPI_ISL_1060784, EPI_ISL_1060785, EPI_ISL_1060788, EPI_ISL_1060790 | NJ Rapid Response Teams | New Jersey Public Health Environmental Laboratories (NJ_PHEL) | Lindsey Bodnar, Shiv Verma, Dana Woell, Byeong Jeong |
| EPI_ISL_1060797 | Mary Eliza Mahoney HC | New Jersey Public Health Environmental Laboratories (NJ_PHEL) | Lindsey Bodnar, Shiv Verma, Dana Woell, Byeong Jeong |
| EPI_ISL_1060798, EPI_ISL_1060799, EPI_ISL_1060800, EPI_ISL_1060801, EPI_ISL_1060802, EPI_ISL_1060803, EPI_ISL_1060804, EPI_ISL_1060805 | NJ Rapid Response Teams | New Jersey Public Health Environmental Laboratories (NJ_PHEL) | Lindsey Bodnar, Shiv Verma, Dana Woell, Byeong Jeong |
| EPI_ISL_1064754, EPI_ISL_1064764 | OCME Office Of Chief Medical Examiner | New York City Public Health Laboratory | Jade Wang, et al. |
| EPI_ISL_1064769 | DOHMH PHL | New York City Public Health Laboratory | Jade Wang, et al. |
| EPI_ISL_1064774 | OCME Office Of Chief Medical Examiner | New York City Public Health Laboratory | Jade Wang, et al. |
| EPI_ISL_1064775 | DOHMH Central Harlem | New York City Public Health Laboratory | Jade Wang, et al. |
| EPI_ISL_1064777 | DOHMH Crown Heights | New York City Public Health Laboratory | Jade Wang, et al. |
| EPI_ISL_1064803 | DOHMH Central Harlem | New York City Public Health Laboratory | Jade Wang, et al. |
| EPI_ISL_1064804 | OCME Office Of Chief Medical Examiner | New York City Public Health Laboratory | Jade Wang, et al. |
| EPI_ISL_1064805, EPI_ISL_1064806 | DOHMH PHL | New York City Public Health Laboratory | Jade Wang, et al. |
| EPI_ISL_1064807 | DOHMH Jamaica | New York City Public Health Laboratory | Jade Wang, et al. |
| EPI_ISL_1064808 | DOHMH Corona | New York City Public Health Laboratory | Jade Wang, et al. |
| EPI_ISL_1064809 | DOHMH Central Harlem | New York City Public Health Laboratory | Jade Wang, et al. |
| EPI_ISL_1064810 | Department of Homeless Services | New York City Public Health Laboratory | Jade Wang, et al. |
| EPI_ISL_1064811 | DOHMH Corona | New York City Public Health Laboratory | Jade Wang, et al. |
| EPI_ISL_1064812 | DOHMH Morrisania | New York City Public Health Laboratory | Jade Wang, et al. |
| EPI_ISL_1064813, EPI_ISL_1064814, EPI_ISL_1064815, EPI_ISL_1064816, EPI_ISL_1064817, EPI_ISL_1064818 | OCME Office Of Chief Medical Examiner | New York City Public Health Laboratory | Jade Wang, et al. |
| EPI_ISL_1064823 | DOHMH Riverside | New York City Public Health Laboratory | Jade Wang, et al. |
| EPI_ISL_1064830 | DOHMH Jamaica | New York City Public Health Laboratory | Jade Wang, et al. |
| EPI_ISL_1064831 | DOHMH Central Harlem | New York City Public Health Laboratory | Jade Wang, et al. |
| EPI_ISL_1064832, EPI_ISL_1064833 | OCME Office Of Chief Medical Examiner | New York City Public Health Laboratory | Jade Wang, et al. |
| EPI_ISL_1064834 | DOHMH Morrisania | New York City Public Health Laboratory | Jade Wang, et al. |
| EPI_ISL_1064835 | DOHMH Crown Heights | New York City Public Health Laboratory | Jade Wang, et al. |
| EPI_ISL_1064836 | DOHMH Fort Greene | New York City Public Health Laboratory | Jade Wang, et al. |
| EPI_ISL_1064837, EPI_ISL_1064838, EPI_ISL_1064839, EPI_ISL_1064840 | DOHMH Jamaica | New York City Public Health Laboratory | Jade Wang, et al. |
| EPI_ISL_1064841, EPI_ISL_1064842 | DOHMH Morrisania | New York City Public Health Laboratory | Jade Wang, et al. |
| EPI_ISL_1064843, EPI_ISL_1064844, EPI_ISL_1064845, EPI_ISL_1064846 | OCME Office Of Chief Medical Examiner | New York City Public Health Laboratory | Jade Wang, et al. |
| EPI_ISL_1064847 | DOHMH Corona | New York City Public Health Laboratory | Jade Wang, et al. |
| EPI_ISL_1064848 | DOHMH Jamaica | New York City Public Health Laboratory | Jade Wang, et al. |
| EPI_ISL_1064849, EPI_ISL_1064850 | DOHMH Morrisania | New York City Public Health Laboratory | Jade Wang, et al. |
| EPI_ISL_1064851, EPI_ISL_1064852 | Department of Homeless Services | New York City Public Health Laboratory | Jade Wang, et al. |
| EPI_ISL_1064853, EPI_ISL_1064854, EPI_ISL_1064855, EPI_ISL_1064856, EPI_ISL_1064857 | OCME Office Of Chief Medical Examiner | New York City Public Health Laboratory | Jade Wang, et al. |
| EPI_ISL_1064858 | DOHMH Jamaica | New York City Public Health Laboratory | Jade Wang, et al. |
| EPI_ISL_1064859, EPI_ISL_1064860 | DOHMH PHL | New York City Public Health Laboratory | Jade Wang, et al. |
| EPI_ISL_1064861 | DOHMH Morrisania | New York City Public Health Laboratory | Jade Wang, et al. |
| EPI_ISL_1064863, EPI_ISL_1064864 | OCME Office Of Chief Medical Examiner | New York City Public Health Laboratory | Jade Wang, et al. |
| EPI_ISL_1069268 | UW Virology Lab | UW Virology Lab | Pavitra Roychoudhury, Hong Xie, Lasata Shrestha, Shah Mohamed Bakhash, Robert Livingston, Michelle Lin, Noah Baker, Meei-Li Huang, Keith R Jerome, Alexander Greninger |
| EPI_ISL_1076611, EPI_ISL_1077830, | Houston Methodist Hospital | Houston Methodist Hospital | S. Wesley Long, Randall J. Olsen, Paul A. Christensen, Sishir Subedi, Robert Olson, James J. Davis, Matthew Ojeda Saavedra, Prasanti Yerramilli, Layne |

| | | | |
|---|---|---|---|
| EPI_ISL_1079178, EPI_ISL_1079543, EPI_ISL_1079930, EPI_ISL_1080059, EPI_ISL_1080164 | | | Pruitt, Kristina Reppond, Madison N. Shyer, Jessica Cambric, Ilya J. Finkelstein, Jimmy Gollihar, and James M. Musser |
| EPI_ISL_1080339 | New Jersey Department of Health, Public Health Laboratories | NJ_PHEL | Lindsey Bodnar, Shiv Verma, Dana Woell, Byeong Jeong |
| EPI_ISL_1080350 | NJ Rapid Response Teams | NJ_PHEL | Lindsey Bodnar, Shiv Verma, Dana Woell, Byeong Jeong |
| EPI_ISL_1080351 | New Jersey Department of Health, Public Health Laboratories | NJ_PHEL | Lindsey Bodnar, Shiv Verma, Dana Woell, Byeong Jeong |

EPI_ISL_1080752, EPI_ISL_1080753, EPI_ISL_1080755, EPI_ISL_1080756, EPI_ISL_1080757, EPI_ISL_1080758, EPI_ISL_1080759, EPI_ISL_1080761, EPI_ISL_1080763, EPI_ISL_1080764, EPI_ISL_1080765, EPI_ISL_1080766, EPI_ISL_1080767, EPI_ISL_1080768, EPI_ISL_1080769, EPI_ISL_1080770, EPI_ISL_1080771, EPI_ISL_1080772, EPI_ISL_1080773, EPI_ISL_1080774, EPI_ISL_1080775, EPI_ISL_1080780, EPI_ISL_1080782, EPI_ISL_1080784, EPI_ISL_1080785, EPI_ISL_1080786, EPI_ISL_1080787, EPI_ISL_1080788, EPI_ISL_1080789, EPI_ISL_1080790, EPI_ISL_1080791, EPI_ISL_1080792, EPI_ISL_1080793, EPI_ISL_1080794, EPI_ISL_1080795, EPI_ISL_1080796, EPI_ISL_1080798, EPI_ISL_1080799, EPI_ISL_1080800, EPI_ISL_1080801, EPI_ISL_1080802, EPI_ISL_1080803, EPI_ISL_1080804, EPI_ISL_1080805, EPI_ISL_1080806, EPI_ISL_1080807, EPI_ISL_1080808

| | | | |
|---|---|---|---|
| see above | Clinical Microbiology Laboratory, NewYork Presbyterian Hospital/Columbia University Irving Medical Center | Uhlemann Laboratory, Columbia University Irving Medical Center | Medini K. Annavajhala, Anne-Catrin Uhlemann, Hiroshi Mohri, David Ho |
| EPI_ISL_1081256 | Maryland Public Health Laboratory | Maryland Public Health Laboratory | Maryland Department of Health Laboratories Administration |
| EPI_ISL_1081297, EPI_ISL_1081298, EPI_ISL_1081299, EPI_ISL_1081302, EPI_ISL_1081304 | Wyoming Public Health Laboratory | Wyoming Public Health Laboratory | Jim Mildenberger, Wanda Manley, Noah Hull, Taylor Fearing, Lynette Gumbleton, Channing Weber, Ashley Norberg, Chayse Rowley, Marley Goetz, Brian Dominguez, Elliot Thomasson, Cari Sloma, and Rob Christensen |

EPI_ISL_1087276, EPI_ISL_1087277, EPI_ISL_1087278, EPI_ISL_1087292, EPI_ISL_1087293, EPI_ISL_1087294, EPI_ISL_1087295, EPI_ISL_1087296, EPI_ISL_1087297, EPI_ISL_1087298, EPI_ISL_1087299, EPI_ISL_1087300, EPI_ISL_1087301

| | | | |
|---|---|---|---|
| see above | NYU Langone Health | Departments of Pathology and Medicine, New York University School of Medicine | Adriana Heguy, Dacia Dimartino, Emily Guzman, Christian Marier, Peter Meyn, Sitharam Ramaswami, Gael Westby, Paul Zappile, Yutong Zhang, Paolo Cotzia, Guiqing Wang |
| EPI_ISL_1089817, EPI_ISL_1089818 | Dutch COVID-19 response team | National Institute for Public Health and the Environment (RIVM) | Adam Meijer, Harry Vennema, Dirk Eggink, Jeroen Cremer, Sharon van den Brink, Bas van der Veer, AnneMarie van den Brandt, Florian Zwagemaker, Dennis Schmitz, Chantal Reusken, on behalf of the national COVID-19 response team |
| EPI_ISL_1091224 | Croatian Institute of Public Health | Croatian Institute of Public Health | Irena Tabain, Ivana Ferenak |
| EPI_ISL_1091942, EPI_ISL_1091958, EPI_ISL_1091959, EPI_ISL_1091982 | Johns Hopkins Hospital Department of Pathology | Johns Hopkins Hospital Department of Pathology | C. Paul Morris, Chun Huai Luo, Adannaya Amadi, Matthew Schwartz, Nicholas Gallagher, Heba H. Mostafa |
| EPI_ISL_1092375, EPI_ISL_1092979, EPI_ISL_1092980 | National Virus Reference Laboratory | National Virus Reference Laboratory | Zoe Yandle, Charlene Bennet, Gabriel Gonzalez, Michael Carr, Jonathan Dean, Cillian F De Gascun |
| EPI_ISL_1094322, EPI_ISL_1094368 | NYSDOH Wadsworth Center, Virology Lab | Respiratory Viruses Branch, Division of Viral Diseases, Centers for Disease Control and Prevention | Krista Queen, Yan Li, Ying Tao, Jing Zhang, Anna Uehara, Anna Montmayeur, Clinton R. Paden, Peter W. Cook, Rachel Marine, Mili Sheth, Jasmine Padilla, Sarah Nobles, Mark Burroughs, Lori Rowe, Haibin Wang, Ben L. Rambo-Martin, Dhwani Batra, Justin Lee, Suxiang Tong |
| EPI_ISL_1094369 | MT Public Health Laboratory | Respiratory Viruses Branch, Division of Viral Diseases, Centers for Disease Control and Prevention | Krista Queen, Yan Li, Ying Tao, Jing Zhang, Anna Uehara, Anna Montmayeur, Clinton R. Paden, Peter W. Cook, Rachel Marine, Mili Sheth, Jasmine Padilla, Sarah Nobles, Mark Burroughs, Lori Rowe, Haibin Wang, Ben L. Rambo-Martin, Dhwani Batra, Justin Lee, Suxiang Tong |
| EPI_ISL_1094370 | NYSDOH Wadsworth Center, Virology Lab | Respiratory Viruses Branch, Division of Viral Diseases, Centers for Disease Control and Prevention | Krista Queen, Yan Li, Ying Tao, Jing Zhang, Anna Uehara, Anna Montmayeur, Clinton R. Paden, Peter W. Cook, Rachel Marine, Mili Sheth, Jasmine Padilla, Sarah Nobles, Mark Burroughs, Lori Rowe, Haibin Wang, Ben L. Rambo-Martin, Dhwani Batra, Justin Lee, Suxiang Tong |
| EPI_ISL_1094371 | WA State Department of Health | Respiratory Viruses Branch, Division of Viral Diseases, Centers for Disease Control and Prevention | Krista Queen, Yan Li, Ying Tao, Jing Zhang, Anna Uehara, Anna Montmayeur, Clinton R. Paden, Peter W. Cook, Rachel Marine, Mili Sheth, Jasmine Padilla, Sarah Nobles, Mark Burroughs, Lori Rowe, Haibin Wang, Ben L. Rambo-Martin, Dhwani Batra, Justin Lee, Suxiang Tong |
| EPI_ISL_1095660 | Fulgent Genetics | Fulgent Genetics | Harry Gao, Mickey Li, John Gao, Joseph Fierro, Benafsh Sapra, Becky Tsai, Yan Meng, Doreen Ng, James Xie |
| EPI_ISL_1097712 | Eurofins Diatherix | Hudsonalpha Genome Sequencing Center | Jane Grimwood, Melissa Williams, Lori H. Handley, Joshua Stough, Leslie Malone, Stefan Brzezinski, Ada Stewart, Teresa Jones, Jenell Webber, John Lovell, Jennifer Cart, and Jeremy Schmutz |

EPI_ISL_1097781, EPI_ISL_1097785, EPI_ISL_1097789, EPI_ISL_1097791, EPI_ISL_1097792, EPI_ISL_1097794, EPI_ISL_1097795, EPI_ISL_1097796, EPI_ISL_1097799, EPI_ISL_1097803, EPI_ISL_1097804, EPI_ISL_1097807, EPI_ISL_1097810, EPI_ISL_1097815, EPI_ISL_1097817, EPI_ISL_1097822, EPI_ISL_1097823, EPI_ISL_1097826, EPI_ISL_1097827, EPI_ISL_1097829, EPI_ISL_1097831, EPI_ISL_1097833, EPI_ISL_1097834, EPI_ISL_1097835, EPI_ISL_1097837, EPI_ISL_1097841, EPI_ISL_1097842, EPI_ISL_1097843, EPI_ISL_1097845, EPI_ISL_1097846, EPI_ISL_1097849, EPI_ISL_1097850, EPI_ISL_1097854, EPI_ISL_1097856, EPI_ISL_1097859, EPI_ISL_1097863, EPI_ISL_1097864, EPI_ISL_1097868, EPI_ISL_1097871, EPI_ISL_1097873, EPI_ISL_1097875, EPI_ISL_1097876, EPI_ISL_1097878, EPI_ISL_1097879, EPI_ISL_1097880, EPI_ISL_1097882, EPI_ISL_1097883, EPI_ISL_1097887, EPI_ISL_1097889, EPI_ISL_1097891, EPI_ISL_1097894, EPI_ISL_1097896, EPI_ISL_1097901, EPI_ISL_1097906, EPI_ISL_1097907, EPI_ISL_1097908, EPI_ISL_1097909, EPI_ISL_1097914, EPI_ISL_1097921, EPI_ISL_1097922, EPI_ISL_1097923, EPI_ISL_1097924, EPI_ISL_1097925, EPI_ISL_1097927, EPI_ISL_1097930, EPI_ISL_1097932, EPI_ISL_1097938, EPI_ISL_1097940, EPI_ISL_1097941, EPI_ISL_1097942, EPI_ISL_1097947, EPI_ISL_1097949, EPI_ISL_1097950, EPI_ISL_1097953, EPI_ISL_1097954, EPI_ISL_1097957, EPI_ISL_1097959, EPI_ISL_1097960, EPI_ISL_1097965, EPI_ISL_1097966, EPI_ISL_1097967, EPI_ISL_1097969, EPI_ISL_1097976, EPI_ISL_1097981, EPI_ISL_1097986, EPI_ISL_1097989, EPI_ISL_1097993, EPI_ISL_1097994, EPI_ISL_1097996, EPI_ISL_1098002, EPI_ISL_1098003, EPI_ISL_1098005, EPI_ISL_1098011, EPI_ISL_1098012, EPI_ISL_1098020, EPI_ISL_1098026, EPI_ISL_1098029, EPI_ISL_1098030, EPI_ISL_1098043, EPI_ISL_1098051, EPI_ISL_1098064, EPI_ISL_1098067, EPI_ISL_1098070, EPI_ISL_1098077, EPI_ISL_1098081, EPI_ISL_1098085, EPI_ISL_1098086, EPI_ISL_1098097, EPI_ISL_1098101, EPI_ISL_1098105, EPI_ISL_1098107, EPI_ISL_1098109, EPI_ISL_1098110, EPI_ISL_1098111, EPI_ISL_1098112, EPI_ISL_1098125, EPI_ISL_1098126, EPI_ISL_1098128, EPI_ISL_1098130, EPI_ISL_1098132, EPI_ISL_1098137, EPI_ISL_1098138, EPI_ISL_1098139, EPI_ISL_1098140, EPI_ISL_1098149, EPI_ISL_1098151, EPI_ISL_1098153, EPI_ISL_1098154, EPI_ISL_1098155, EPI_ISL_1098163, EPI_ISL_1098164, EPI_ISL_1098167, EPI_ISL_1098176, EPI_ISL_1098180, EPI_ISL_1098181, EPI_ISL_1098186, EPI_ISL_1098190, EPI_ISL_1098191, EPI_ISL_1098192, EPI_ISL_1098194, EPI_ISL_1098199, EPI_ISL_1098202, EPI_ISL_1098203, EPI_ISL_1098204, EPI_ISL_1098209, EPI_ISL_1098218, EPI_ISL_1098224, EPI_ISL_1098225, EPI_ISL_1098235, EPI_ISL_1098238, EPI_ISL_1098240, EPI_ISL_1098241, EPI_ISL_1098257, EPI_ISL_1098258, EPI_ISL_1098259, EPI_ISL_1098263, EPI_ISL_1098267, EPI_ISL_1098281, EPI_ISL_1098285, EPI_ISL_1098290, EPI_ISL_1098301, EPI_ISL_1098302, EPI_ISL_1098303, EPI_ISL_1098305, EPI_ISL_1098306, EPI_ISL_1098309, EPI_ISL_1098310, EPI_ISL_1098311, EPI_ISL_1098312, EPI_ISL_1098324, EPI_ISL_1098325, EPI_ISL_1098326, EPI_ISL_1098327, EPI_ISL_1098335, EPI_ISL_1098339, EPI_ISL_1098344, EPI_ISL_1098346, EPI_ISL_1098347, EPI_ISL_1098348, EPI_ISL_1098349, EPI_ISL_1098355, EPI_ISL_1098356, EPI_ISL_1098357, EPI_ISL_1098358, EPI_ISL_1098360, EPI_ISL_1098363, EPI_ISL_1098365, EPI_ISL_1098378, EPI_ISL_1098379, EPI_ISL_1098381, EPI_ISL_1098383, EPI_ISL_1098387, EPI_ISL_1098389, EPI_ISL_1098390, EPI_ISL_1098391, EPI_ISL_1098394, EPI_ISL_1098399, EPI_ISL_1098400, EPI_ISL_1098403, EPI_ISL_1098405, EPI_ISL_1098406, EPI_ISL_1098421, EPI_ISL_1098425, EPI_ISL_1098426, EPI_ISL_1098428, EPI_ISL_1098429, EPI_ISL_1098431, EPI_ISL_1098432, EPI_ISL_1098433, EPI_ISL_1098437, EPI_ISL_1098438, EPI_ISL_1098441, EPI_ISL_1098445, EPI_ISL_1098446, EPI_ISL_1098447, EPI_ISL_1098448, EPI_ISL_1098450, EPI_ISL_1098455, EPI_ISL_1098456, EPI_ISL_1098459, EPI_ISL_1098460, EPI_ISL_1098463, EPI_ISL_1098465, EPI_ISL_1098466, EPI_ISL_1098469, EPI_ISL_1098470, EPI_ISL_1098472, EPI_ISL_1098473, EPI_ISL_1098476, EPI_ISL_1098482, EPI_ISL_1098487, EPI_ISL_1098489, EPI_ISL_1098493, EPI_ISL_1098494, EPI_ISL_1098495, EPI_ISL_1098497, EPI_ISL_1098500, EPI_ISL_1098503, EPI_ISL_1098504, EPI_ISL_1098506, EPI_ISL_1098510, EPI_ISL_1098511, EPI_ISL_1098514, EPI_ISL_1098519, EPI_ISL_1098522, EPI_ISL_1098523, EPI_ISL_1098524, EPI_ISL_1098525, EPI_ISL_1098532, EPI_ISL_1098534, EPI_ISL_1098535, EPI_ISL_1098536, EPI_ISL_1098538, EPI_ISL_1098541, EPI_ISL_1098542, EPI_ISL_1098543, EPI_ISL_1098550, EPI_ISL_1098555, EPI_ISL_1098564, EPI_ISL_1098565, EPI_ISL_1098567, EPI_ISL_1098568, EPI_ISL_1098570, EPI_ISL_1098571, EPI_ISL_1098574, EPI_ISL_1098576, EPI_ISL_1098578, EPI_ISL_1098583, EPI_ISL_1098586, EPI_ISL_1098587, EPI_ISL_1098594, EPI_ISL_1098595, EPI_ISL_1098596

| | | | |
|---|---|---|---|
| see above | Pandemic Response Lab - NYC | Pandemic Response Lab, R&D | Henry Lee, Michael Hammerling, Melissa Hopkins, Cybill del Castillo, Shinyoung Clair Kang, William Ward, Pradeep Bugga, Haiping Hao, Jon Laurent |
| EPI_ISL_1105341 | University College London Hospital | COVID-19 Genomics UK (COG-UK) Consortium | Judith Heaney, Matthew Byott, Catherine Houlihan, Dan Frampton, Stuart Kirk, Moira Spyer and Eleni Nastouli |
| EPI_ISL_1109521 | University Hospital for Infectious Diseases | Croatian Institute of Public Health | Irena Tabain, Ivana Ferenak |
| EPI_ISL_1109557 | "Dr. Andrija Stampar" Teaching Institute of Public Health | Croatian Institute of Public Health | Irena Tabain, Ivana Ferenak |
| EPI_ISL_1111536, EPI_ISL_1111542, EPI_ISL_1111543, EPI_ISL_1111544, EPI_ISL_1111546 | Wyoming Public Health Laboratory | Wyoming Public Health Laboratory | Jim Mildenberger, Wanda Manley, Noah Hull, Taylor Fearing, Lynette Gumbleton, Channing Weber, Ashley Norberg, Chayse Rowley, Marley Goetz, Brian Dominguez, Elliot Thomasson, Cari Sloma, and Rob Christensen |
| EPI_ISL_1113142, EPI_ISL_1113147, EPI_ISL_1113148, EPI_ISL_1113149 | NJ Rapid Response Teams | New Jersey Public Health and Environmental Laboratories (PHEL) | Lindsey Bodnar, Shiv Verma, Dana Woell, Byeong Jeong |
| EPI_ISL_1113151 | Mary Eliza Mahoney HC | New Jersey Public Health and Environmental Laboratories (PHEL) | Lindsey Bodnar, Shiv Verma, Dana Woell, Byeong Jeong |
| EPI_ISL_1113155 | NJ Rapid Response Teams | New Jersey Public Health and Environmental Laboratories (PHEL) | Lindsey Bodnar, Shiv Verma, Dana Woell, Byeong Jeong |
| EPI_ISL_1113156 | New Jersey Department of Health, Public Health Laboratories | New Jersey Public Health and Environmental Laboratories (PHEL) | Lindsey Bodnar, Shiv Verma, Dana Woell, Byeong Jeong |
| EPI_ISL_1113170 | Mary Eliza Mahoney HC | New Jersey Public Health and Environmental Laboratories (PHEL) | Lindsey Bodnar, Shiv Verma, Dana Woell, Byeong Jeong |
| EPI_ISL_1113171 | New Jersey Department of Health, Public Health Laboratories | New Jersey Public Health and Environmental Laboratories (PHEL) | Lindsey Bodnar, Shiv Verma, Dana Woell, Byeong Jeong |

| | | | |
|---|---|---|---|
| EPI_ISL_1114471 | Department of Health and Mental Hygiene Central Harlem | New York City Public Health Laboratory | Jade Wang, et al. |
| EPI_ISL_1114472, EPI_ISL_1114474 | Department of Health and Mental Hygiene Corona | New York City Public Health Laboratory | Jade Wang, et al. |
| EPI_ISL_1114478, EPI_ISL_1114481 | Department of Health and Mental Hygiene Jamaica | New York City Public Health Laboratory | Jade Wang, et al. |
| EPI_ISL_1114482, EPI_ISL_1114483 | Department of Health and Mental Hygiene Public Health Laboratory | New York City Public Health Laboratory | Jade Wang, et al. |
| EPI_ISL_1114488, EPI_ISL_1114491 | Department of Health and Mental Hygiene Morrisania | New York City Public Health Laboratory | Jade Wang, et al. |
| EPI_ISL_1114493 | Department of Health and Mental Hygiene Central Harlem | New York City Public Health Laboratory | Jade Wang, et al. |
| EPI_ISL_1114495, EPI_ISL_1114496 | Department of Health and Mental Hygiene Fort Greene | New York City Public Health Laboratory | Jade Wang, et al. |
| EPI_ISL_1114500, EPI_ISL_1114501 | Department of Health and Mental Hygiene Chelsea | New York City Public Health Laboratory | Jade Wang, et al. |
| EPI_ISL_1114502 | Department of Health and Mental Hygiene Crown Heights | New York City Public Health Laboratory | Jade Wang, et al. |
| EPI_ISL_1114504, EPI_ISL_1114505, EPI_ISL_1114507 | Department of Health and Mental Hygiene Corona | New York City Public Health Laboratory | Jade Wang, et al. |
| EPI_ISL_1114510 | Department of Health and Mental Hygiene Public Health Laboratory | New York City Public Health Laboratory | Jade Wang, et al. |
| EPI_ISL_1114514, EPI_ISL_1114515 | Department of Health and Mental Hygiene Corona | New York City Public Health Laboratory | Jade Wang, et al. |
| EPI_ISL_1114516, EPI_ISL_1114517 | Department of Health and Mental Hygiene Crown Heights | New York City Public Health Laboratory | Jade Wang, et al. |
| EPI_ISL_1114520, EPI_ISL_1114521, EPI_ISL_1114522, EPI_ISL_1114523 | Department of Health and Mental Hygiene Public Health Laboratory | New York City Public Health Laboratory | Jade Wang, et al. |
| EPI_ISL_1114524 | Department of Health and Mental Hygiene Chelsea | New York City Public Health Laboratory | Jade Wang, et al. |
| EPI_ISL_1114525, EPI_ISL_1114527, EPI_ISL_1114530 | OCME Office Of Chief Medical Examiner | New York City Public Health Laboratory | Jade Wang, et al. |
| EPI_ISL_1116479, EPI_ISL_1116483, EPI_ISL_1116488, EPI_ISL_1116516, EPI_ISL_1116517, EPI_ISL_1116518, EPI_ISL_1116519, EPI_ISL_1116520, EPI_ISL_1116521, EPI_ISL_1116522, EPI_ISL_1116523, EPI_ISL_1116525, EPI_ISL_1116526, EPI_ISL_1116527, EPI_ISL_1116529, EPI_ISL_1116530, EPI_ISL_1116531, EPI_ISL_1116532 | | | |
| see above | NYU Langone Health | Departments of Pathology and Medicine, New York University School of Medicine | Adriana Heguy, Dacia Dimartino, Emily Guzman, Christian Marier, Peter Meyn, Sitharam Ramaswami, Gael Westby, Paul Zappile, Yutong Zhang, Paolo Cotzia, Guiqing Wang |
| EPI_ISL_1121020 | Wyoming Public Health Laboratory | Wyoming Public Health Laboratory | Jim Mildenberger, Wanda Manley, Noah Hull, Taylor Fearing, Lynette Gumbleton, Channing Weber, Ashley Norberg, Chayse Rowley, Marley Goetz, Brian Dominguez, Elliot Thomasson, Cari Sloma, and Rob Christensen |
| EPI_ISL_1121888 | Department of Health & Mental Hygiene Morrisania | New York City Public Health Laboratory | Jade Wang, et al. |
| EPI_ISL_1121892 | Department of Health & Mental Hygiene Crown Heights | New York City Public Health Laboratory | Jade Wang, et al. |
| EPI_ISL_1121893 | Department of Health & Mental Hygiene Central Harlem | New York City Public Health Laboratory | Jade Wang, et al. |
| EPI_ISL_1121895 | OCME Office Of Chief Medical Examiner | New York City Public Health Laboratory | Jade Wang, et al. |
| EPI_ISL_1121899 | Department of Health & Mental Hygiene Corona | New York City Public Health Laboratory | Jade Wang, et al. |
| EPI_ISL_1121900 | Department of Health & Mental Hygiene Central Harlem | New York City Public Health Laboratory | Jade Wang, et al. |
| EPI_ISL_1121902 | Department of Health & Mental Hygiene Fort Greene | New York City Public Health Laboratory | Jade Wang, et al. |
| EPI_ISL_1121904 | Department of Health & Mental Hygiene Crown Heights | New York City Public Health Laboratory | Jade Wang, et al. |
| EPI_ISL_1121905, EPI_ISL_1121906, EPI_ISL_1121907 | Department of Health & Mental Hygiene Corona | New York City Public Health Laboratory | Jade Wang, et al. |
| EPI_ISL_1121909, EPI_ISL_1121910, EPI_ISL_1121911 | Department of Health & Mental Hygiene PHL | New York City Public Health Laboratory | Jade Wang, et al. |
| EPI_ISL_1121914 | Department of Health & Mental Hygiene Riverside | New York City Public Health Laboratory | Jade Wang, et al. |
| EPI_ISL_1121916, EPI_ISL_1121918 | Department of Health & Mental Hygiene Jamaica | New York City Public Health Laboratory | Jade Wang, et al. |
| EPI_ISL_1121919, EPI_ISL_1121921 | Department of Health & Mental Hygiene Morrisania | New York City Public Health Laboratory | Jade Wang, et al. |
| EPI_ISL_1121929, EPI_ISL_1121931 | Department of Health & Mental Hygiene PHL | New York City Public Health Laboratory | Jade Wang, et al. |
| EPI_ISL_1121935 | Department of Health & Mental Hygiene Corona | New York City Public Health Laboratory | Jade Wang, et al. |
| EPI_ISL_1121938 | Department of Health & Mental Hygiene Crown Heights | New York City Public Health Laboratory | Jade Wang, et al. |
| EPI_ISL_1121940, EPI_ISL_1121941, EPI_ISL_1121942, EPI_ISL_1121947, EPI_ISL_1121948, EPI_ISL_1121949 | Department of Health & Mental Hygiene Morrisania | New York City Public Health Laboratory | Jade Wang, et al. |
| EPI_ISL_1121950 | Department of Health & Mental Hygiene Central Harlem | New York City Public Health Laboratory | Jade Wang, et al. |
| EPI_ISL_1121953, EPI_ISL_1121957, EPI_ISL_1121959 | Department of Health & Mental Hygiene Jamaica | New York City Public Health Laboratory | Jade Wang, et al. |
| EPI_ISL_1122114 | OHSU Lab Services Molecular Microbiology Lab | Oregon SARS-CoV-2 Genome Sequencing Center | Brendan L. O'Connell, Sally Grindstaff, Kayla Carter, Ruth V. Nichols, Alec J. Hirsch, Donna Hansel, Guang Fan, Xuan, Qin, Daniel N. Streblow, William B. Messer, Andrew C. Adey, Benjamin N. Bimber, Brian J. O'Roak |
| EPI_ISL_1132286 | Florida Bureau of Public Health Laboratories | Florida Bureau of Public Health Laboratories | Sarah Schmedes, Jason Blanton |
| EPI_ISL_1135892 | Nevada State Public Health Laboratory | Nevada State Public Health Laboratory | Andrew Gorzalski, Mark Pandori |
| EPI_ISL_1137004, EPI_ISL_1137005, EPI_ISL_1137006, EPI_ISL_1137008 | Delaware Public Health Lab | Delaware Public Health Lab | Gregory Hovan |
| EPI_ISL_1137499, EPI_ISL_1137500 | New Jersey Department of Health, Public Health and Environmental Laboratories | New Jersey Public Health and Environmental Laboratories | Lindsey Bodnar, Shiv K. Verma, Dana Woell, Byeong Jeong |
| EPI_ISL_1153659 | MVZ Labor Dr. Fenner und Kollegen (Standort Hamburg) | Robert Koch Institute | unknown |
| EPI_ISL_1157425 | Northwestern Memorial Hospital | Northwestern University - Ozer Lab | Ramon Lorenzo-Redondo, Lacy M. Simons, Taylor J. Dean, Chad J. Achenbach, Lawrence J. Jennings, Chao Qi, Michael G. Ison, Judd F. Hultquist, Egon A. Ozer |
| EPI_ISL_1158456 | Michigan Department of Health and Human Services, Bureau of Laboratories | Michigan Department of Health and Human Services, Bureau of Laboratories | Blankenship HM, Riner D, Soehnlen MK |
| EPI_ISL_1160097, EPI_ISL_1160290, EPI_ISL_1160303, EPI_ISL_1160377, EPI_ISL_1160396, EPI_ISL_1160432, EPI_ISL_1160452, EPI_ISL_1160486, EPI_ISL_1160533, EPI_ISL_1160546, EPI_ISL_1160604, EPI_ISL_1160628, EPI_ISL_1160662, EPI_ISL_1160708, EPI_ISL_1162091, EPI_ISL_1162118, EPI_ISL_1162141, | | | |

| | | | |
|---|---|---|---|
| EPI_ISL_1162166, EPI_ISL_1162184, EPI_ISL_1162216, EPI_ISL_1162237, EPI_ISL_1162246, EPI_ISL_1162268, EPI_ISL_1162347, EPI_ISL_1162392, EPI_ISL_1162393, EPI_ISL_1162403, EPI_ISL_1162570, EPI_ISL_1162589, EPI_ISL_1162682 | | | |
| see above | Laboratory Corporation of America | Respiratory Viruses Branch, Division of Viral Diseases, Centers for Disease Control and Prevention | Peter W. Cook, Dakota Howard, Dhwani Batra, Ben L. Rambo-Martin, Minoo Agarwal, Eyad Almasri Debbie Boles, Ayla Burns, Nuthawin Charoensri, Oren Cohen, Susan Countryman, Mary Ann Cristobal, Bobbi Croy, Suzanne Dale, Hrushikesh Deshmukh, Amanda Douglas, Vincent Drouillon, Marcia Eisenberg, Howard Engler, Rama Ghatti, Prashant Gupta, Susan Hicks, Jake Humphrey, Lax Iyer, Manoj Jain, Mohan Kolli, Brian Krueger, Tim Kuphal, Stanley Letovsky, Michael Levandoski, Craig Lukasik, Jonathan Meltzer, Brian Norvell, Mindy Nye, Scott Parker, Christos Petropoulos, John Pruitt, Steven Ragan, Scott Ryan, Mike Sapeta, Jana Schroth, Suresh Babu Selvaraju, Goran Stevovic, Amanda Suchanek, Andrea Throop, Lyndon Tilson, Thomas Urban, Joe Voshell, Kimberly Wagner, Jonathan Williams, Mary Williamson, Qian Zeng, Tricia Zwiefelhofer, Clinton R. Paden, Suxiang Tong, Duncan MacCannell |
| EPI_ISL_1163563, EPI_ISL_1163565, EPI_ISL_1163616, EPI_ISL_1163637 | Maine HETL | Tewhey Lab, The Jackson Laboratory | Matluk,N., Dewey,H., Iosue,F., Barter,M., Lynch,R., Munger,H. and Tewhey,R. |
| EPI_ISL_1163665 | Tempus | Grubaugh Lab - Yale School of Public Health | Joseph Fauver, Tara Alpert, Anderson Brito, Mallery Breban, Anne Wyllie, Chantal Vogels, Mary Petrone, Annie Watkins, Chaney Kalinich, Isabel Ott, Nathan Grubaugh |
| EPI_ISL_1163684, EPI_ISL_1163685, EPI_ISL_1163686 | Yale Clinical Virology Lab | Grubaugh Lab - Yale School of Public Health | Joseph Fauver, Tara Alpert, Anderson Brito, Mallery Breban, Anne Wyllie, Chantal Vogels, Mary Petrone, Annie Watkins, Chaney Kalinich, Isabel Ott, Nathan Grubaugh |
| EPI_ISL_1164075 | National Virus Reference Laboratory | National Virus Reference Laboratory | Zoe Yandle, Charlene Bennet, Gabriel Gonzalez, Michael Carr, Jonathan Dean, Cillian F De Gascun |
| EPI_ISL_1164855 | Diagnostic Genomics Lab and Functional Genomics Core University of South Carolina | Diagnostic Genomics Lab and Functional Genomics Core, University of South Carolina | Carolyn Banister, Hao Ji, Diego Altomare, Herr Doktor Professor Genomiklaborsdirektor Universitätkovidenpandemieretter Phillip Buckhaults, Michael Shtutman, Helmut Albrecht |
| EPI_ISL_1168237 | TXDSHS | TXDSHS | Rashmi Tuladhar, Bonnie Oh, Jenny Zhang, Maliha Rahman, Mayela Pedrueza, Anita Pokharel, Myong Koag, Chun Wang, Rachel Lee, Grace Kubin |
| EPI_ISL_1168763, EPI_ISL_1168764 | Yale Pathology Lab | Grubaugh Lab - Yale School of Public Health | Tara Alpert, Joseph Fauver, Chen Liu, Pei Hui, Jianhui Wang, Susan Bell and Han Zhou, Anderson Brito, Mallery Breban, Anne Wyllie, Chantal Vogels, Mary Petrone, Chaney Kalinich, Isabel Ott, Arnau Casanovas, Catherine Muenker, Adam Moore, Alice Lu, Maria Tokuyama, Patrick Wong, Peiwen Lu, Saad Omer, Richard Martinello, Allison Nelson, Shelli Farhadian, Akiko Iwasaki, Charlese Dela Cruz, Albert Ko, Nathan Grubaugh |
| EPI_ISL_1171780, EPI_ISL_1171797, EPI_ISL_1171798, EPI_ISL_1171813, EPI_ISL_1171834, EPI_ISL_1171836, EPI_ISL_1171837 | Johns Hopkins Hospital Department of Pathology | Johns Hopkins Hospital Department of Pathology | C. Paul Morris, Chun Huai Luo, Adannaya Amadi, Matthew Schwartz, Nicholas Gallagher, Heba H. Mostafa |
| EPI_ISL_1172111, EPI_ISL_1172112, EPI_ISL_1172115, EPI_ISL_1172116, EPI_ISL_1172121, EPI_ISL_1172122, EPI_ISL_1172128, EPI_ISL_1172132, EPI_ISL_1172134, EPI_ISL_1172135, EPI_ISL_1172137, EPI_ISL_1172139, EPI_ISL_1172140, EPI_ISL_1172141, EPI_ISL_1172144, EPI_ISL_1172145, EPI_ISL_1172146, EPI_ISL_1172147, EPI_ISL_1172149, EPI_ISL_1172154, EPI_ISL_1172157, EPI_ISL_1172158, EPI_ISL_1172160, EPI_ISL_1172161, EPI_ISL_1172164, EPI_ISL_1172166, EPI_ISL_1172169, EPI_ISL_1172170, EPI_ISL_1172174, EPI_ISL_1172178, EPI_ISL_1172181, EPI_ISL_1172189, EPI_ISL_1172190, EPI_ISL_1172193, EPI_ISL_1172194, EPI_ISL_1172196, EPI_ISL_1172200, EPI_ISL_1172201, EPI_ISL_1172205, EPI_ISL_1172208, EPI_ISL_1172209, EPI_ISL_1172211, EPI_ISL_1172212, EPI_ISL_1172213, EPI_ISL_1172216, EPI_ISL_1172217, EPI_ISL_1172218, EPI_ISL_1172220, EPI_ISL_1172228, EPI_ISL_1172229, EPI_ISL_1172234, EPI_ISL_1172239, EPI_ISL_1172242, EPI_ISL_1172247, EPI_ISL_1172250, EPI_ISL_1172253, EPI_ISL_1172254, EPI_ISL_1172256, EPI_ISL_1172261, EPI_ISL_1172263, EPI_ISL_1172264, EPI_ISL_1172268, EPI_ISL_1172273, EPI_ISL_1172278, EPI_ISL_1172284, EPI_ISL_1172286, EPI_ISL_1172287, EPI_ISL_1172288, EPI_ISL_1172295, EPI_ISL_1172300, EPI_ISL_1172303, EPI_ISL_1172304, EPI_ISL_1172305, EPI_ISL_1172306, EPI_ISL_1172308, EPI_ISL_1172309, EPI_ISL_1172313, EPI_ISL_1172316, EPI_ISL_1172321, EPI_ISL_1172327, EPI_ISL_1172328, EPI_ISL_1172330, EPI_ISL_1172331, EPI_ISL_1172332, EPI_ISL_1172333, EPI_ISL_1172335, EPI_ISL_1172336, EPI_ISL_1172338, EPI_ISL_1172340, EPI_ISL_1172347, EPI_ISL_1172354, EPI_ISL_1172359, EPI_ISL_1172360, EPI_ISL_1172363, EPI_ISL_1172366, EPI_ISL_1172373, EPI_ISL_1172377, EPI_ISL_1172378, EPI_ISL_1172381, EPI_ISL_1172382, EPI_ISL_1172388, EPI_ISL_1172389, EPI_ISL_1172390, EPI_ISL_1172391, EPI_ISL_1172392, EPI_ISL_1172393, EPI_ISL_1172397, EPI_ISL_1172399, EPI_ISL_1172405, EPI_ISL_1172406, EPI_ISL_1172409, EPI_ISL_1172410, EPI_ISL_1172412, EPI_ISL_1172414, EPI_ISL_1172419, EPI_ISL_1172425, EPI_ISL_1172427, EPI_ISL_1172429, EPI_ISL_1172435, EPI_ISL_1172439, EPI_ISL_1172449, EPI_ISL_1172450, EPI_ISL_1172451, EPI_ISL_1172452, EPI_ISL_1172453, EPI_ISL_1172454, EPI_ISL_1172456, EPI_ISL_1172460, EPI_ISL_1172461, EPI_ISL_1172469, EPI_ISL_1172471, EPI_ISL_1172472, EPI_ISL_1172476, EPI_ISL_1172480, EPI_ISL_1172482, EPI_ISL_1172486, EPI_ISL_1172490, EPI_ISL_1172493, EPI_ISL_1172494, EPI_ISL_1172498, EPI_ISL_1172502, EPI_ISL_1172505, EPI_ISL_1172508, EPI_ISL_1172510, EPI_ISL_1172514, EPI_ISL_1172515, EPI_ISL_1172516, EPI_ISL_1172518, EPI_ISL_1172523, EPI_ISL_1172525, EPI_ISL_1172530, EPI_ISL_1172533, EPI_ISL_1172534, EPI_ISL_1172538, EPI_ISL_1172541, EPI_ISL_1172542, EPI_ISL_1172543, EPI_ISL_1172544, EPI_ISL_1172548, EPI_ISL_1172552, EPI_ISL_1172554, EPI_ISL_1172557, EPI_ISL_1172558, EPI_ISL_1172559, EPI_ISL_1172560, EPI_ISL_1172562, EPI_ISL_1172564, EPI_ISL_1172566, EPI_ISL_1172573, EPI_ISL_1172575, EPI_ISL_1172577, EPI_ISL_1172581, EPI_ISL_1172582, EPI_ISL_1172586, EPI_ISL_1172588, EPI_ISL_1172591, EPI_ISL_1172592, EPI_ISL_1172593, EPI_ISL_1172594, EPI_ISL_1172595, EPI_ISL_1172596, EPI_ISL_1172597, EPI_ISL_1172600, EPI_ISL_1172601, EPI_ISL_1172605, EPI_ISL_1172606, EPI_ISL_1172612, EPI_ISL_1172613, EPI_ISL_1172615, EPI_ISL_1172616, EPI_ISL_1172620, EPI_ISL_1172621, EPI_ISL_1172622, EPI_ISL_1172626, EPI_ISL_1172627, EPI_ISL_1172640, EPI_ISL_1172644, EPI_ISL_1172651, EPI_ISL_1172652, EPI_ISL_1172654, EPI_ISL_1172655, EPI_ISL_1172659, EPI_ISL_1172660, EPI_ISL_1172662, EPI_ISL_1172663, EPI_ISL_1172664, EPI_ISL_1172665, EPI_ISL_1172669, EPI_ISL_1172676, EPI_ISL_1172677, EPI_ISL_1172678, EPI_ISL_1172679, EPI_ISL_1172680, EPI_ISL_1172681, EPI_ISL_1172682, EPI_ISL_1172683, EPI_ISL_1172684, EPI_ISL_1172686, EPI_ISL_1172687, EPI_ISL_1172689, EPI_ISL_1172694, EPI_ISL_1172701, EPI_ISL_1172702, EPI_ISL_1172713, EPI_ISL_1172714, EPI_ISL_1172715, EPI_ISL_1172717, EPI_ISL_1172718, EPI_ISL_1172721, EPI_ISL_1172722, EPI_ISL_1172725, EPI_ISL_1172727, EPI_ISL_1172728, EPI_ISL_1172730, EPI_ISL_1172731, EPI_ISL_1172732, EPI_ISL_1172735, EPI_ISL_1172736, EPI_ISL_1172737, EPI_ISL_1172738, EPI_ISL_1172739, EPI_ISL_1172741, EPI_ISL_1172742, EPI_ISL_1172746, EPI_ISL_1172747, EPI_ISL_1172748, EPI_ISL_1172749, EPI_ISL_1172750, EPI_ISL_1172753, EPI_ISL_1172756, EPI_ISL_1172762, EPI_ISL_1172763, EPI_ISL_1172766, EPI_ISL_1172768, EPI_ISL_1172769, EPI_ISL_1172770, EPI_ISL_1172775, EPI_ISL_1172777, EPI_ISL_1172778, EPI_ISL_1172779, EPI_ISL_1172780, EPI_ISL_1172781, EPI_ISL_1172782, EPI_ISL_1172783, EPI_ISL_1172789, EPI_ISL_1172791, EPI_ISL_1172796, EPI_ISL_1172799, EPI_ISL_1172800, EPI_ISL_1172801, EPI_ISL_1172802, EPI_ISL_1172804, EPI_ISL_1172806, EPI_ISL_1172808, EPI_ISL_1172810, EPI_ISL_1172811, EPI_ISL_1172815, EPI_ISL_1172816, EPI_ISL_1172817, EPI_ISL_1172818, EPI_ISL_1172821, EPI_ISL_1172823, EPI_ISL_1172824, EPI_ISL_1172825, EPI_ISL_1172826, EPI_ISL_1172829, EPI_ISL_1172830, EPI_ISL_1172831, EPI_ISL_1172833, EPI_ISL_1172834, EPI_ISL_1172835, EPI_ISL_1172836, EPI_ISL_1172838, EPI_ISL_1172843, EPI_ISL_1172846, EPI_ISL_1172847, EPI_ISL_1172848, EPI_ISL_1172850, EPI_ISL_1172852, EPI_ISL_1172853, EPI_ISL_1172855, EPI_ISL_1172856, EPI_ISL_1172860, EPI_ISL_1172861, EPI_ISL_1172863, EPI_ISL_1172867, EPI_ISL_1172868, EPI_ISL_1172869, EPI_ISL_1172871, EPI_ISL_1172878, EPI_ISL_1172881, EPI_ISL_1172882, EPI_ISL_1172886, EPI_ISL_1172888, EPI_ISL_1172890, EPI_ISL_1172892, EPI_ISL_1172894, EPI_ISL_1172897, EPI_ISL_1172904, EPI_ISL_1172905, EPI_ISL_1172909, EPI_ISL_1172913, EPI_ISL_1172917, EPI_ISL_1172918, EPI_ISL_1172919, EPI_ISL_1172920, EPI_ISL_1172924, EPI_ISL_1172925, EPI_ISL_1172926, EPI_ISL_1172929, EPI_ISL_1172931, EPI_ISL_1172932, EPI_ISL_1172933, EPI_ISL_1172940, EPI_ISL_1172942, EPI_ISL_1172944, EPI_ISL_1172949, EPI_ISL_1172950, EPI_ISL_1172951, EPI_ISL_1172952, EPI_ISL_1172953, EPI_ISL_1172954, EPI_ISL_1172955, EPI_ISL_1172956, EPI_ISL_1172958, EPI_ISL_1172959, EPI_ISL_1172962, EPI_ISL_1172963, EPI_ISL_1172966, EPI_ISL_1172970, EPI_ISL_1172973, EPI_ISL_1172975, EPI_ISL_1172981, EPI_ISL_1172983, EPI_ISL_1172986, EPI_ISL_1172988, EPI_ISL_1172989, EPI_ISL_1172995, EPI_ISL_1173001, EPI_ISL_1173002, EPI_ISL_1173006, EPI_ISL_1173009, EPI_ISL_1173015, EPI_ISL_1173016, EPI_ISL_1173021, EPI_ISL_1173024, EPI_ISL_1173025, EPI_ISL_1173030, EPI_ISL_1173034, EPI_ISL_1173036, EPI_ISL_1173037, EPI_ISL_1173038, EPI_ISL_1173040, EPI_ISL_1173043, EPI_ISL_1173044, EPI_ISL_1173045, EPI_ISL_1173046, EPI_ISL_1173049, EPI_ISL_1173051, EPI_ISL_1173053, EPI_ISL_1173054, EPI_ISL_1173058, EPI_ISL_1173061, EPI_ISL_1173062, EPI_ISL_1173070, EPI_ISL_1173071, EPI_ISL_1173072, EPI_ISL_1173075, EPI_ISL_1173078, EPI_ISL_1173079, EPI_ISL_1173081, EPI_ISL_1173082, EPI_ISL_1173083, EPI_ISL_1173089, EPI_ISL_1173090, EPI_ISL_1173091, EPI_ISL_1173092, EPI_ISL_1173093, EPI_ISL_1173098, EPI_ISL_1173100, EPI_ISL_1173103, EPI_ISL_1173104, EPI_ISL_1173105, EPI_ISL_1173107, EPI_ISL_1173109, EPI_ISL_1173110, EPI_ISL_1173111, EPI_ISL_1173116, EPI_ISL_1173120, EPI_ISL_1173125, EPI_ISL_1173126, EPI_ISL_1173127, EPI_ISL_1173128, EPI_ISL_1173129, EPI_ISL_1173130, EPI_ISL_1173133, EPI_ISL_1173134, EPI_ISL_1173138, EPI_ISL_1173139, EPI_ISL_1173140, EPI_ISL_1173141, EPI_ISL_1173142, EPI_ISL_1173143, EPI_ISL_1173144, EPI_ISL_1173148, EPI_ISL_1173149, EPI_ISL_1173150, EPI_ISL_1173151, EPI_ISL_1173152, EPI_ISL_1173153, EPI_ISL_1173157 | | | |
| see above | Pandemic Response Lab - NYC | Pandemic Response Lab, R&D | Henry Lee, Michael Hammerling, Melissa Hopkins, Cybill del Castillo, Shinyoung Clair Kang, William Ward, Pradeep Bugga, Haiping Hao, Jon Laurent |
| EPI_ISL_1181662, EPI_ISL_1181669, EPI_ISL_1181674, EPI_ISL_1181675, EPI_ISL_1181677, EPI_ISL_1181688 | DPHL | Delaware Public Health Lab | Gregory Hovan |
| EPI_ISL_1182008 | Maryland Public Health Laboratory | Maryland Public Health Laboratory | Maryland Department of Health Laboratories Administration |
| EPI_ISL_1182022 | Yale Clinical Virology Lab | Grubaugh Lab - Yale School of Public Health | Joseph Fauver, Mallery Breban, Isabell Ott, Tara Alpert, Mary Petrone, Anderson Brito, Chantal Vogels, Annie Watkins, Chaney Kalinich, Marie L. Landry, Nathan Grubaugh |
| EPI_ISL_1184099, EPI_ISL_1184102, EPI_ISL_1184103 | New Jersey Department of Health, Public Health and Environmental Laboratories | New Jersey Public Health and Environmental Laboratories | Lindsey Bodnar, Shiv K. Verma, Dana Woell, Byeong Jeong |
| EPI_ISL_1191460 | National Virus Reference Laboratory | National Virus Reference Laboratory | Zoe Yandle, Charlene Bennet, Gabriel Gonzalez, Michael Carr, Jonathan Dean, Cillian F De Gascun |
| EPI_ISL_1191574 | Florida Bureau of Public Health Laboratories | Florida Bureau of Public Health Laboratories | Sarah Schmedes, Jason Blanton |
| EPI_ISL_1195745, EPI_ISL_1195749, EPI_ISL_1195753, EPI_ISL_1195757, EPI_ISL_1195759, EPI_ISL_1195761, EPI_ISL_1195762, EPI_ISL_1195767, EPI_ISL_1195768, EPI_ISL_1195775, EPI_ISL_1195776, EPI_ISL_1195778, EPI_ISL_1195780, EPI_ISL_1195785, EPI_ISL_1195789, EPI_ISL_1195800, EPI_ISL_1195810, EPI_ISL_1195811, EPI_ISL_1195814, EPI_ISL_1195825, EPI_ISL_1195827, EPI_ISL_1195830, EPI_ISL_1195831, EPI_ISL_1195843, EPI_ISL_1195845, EPI_ISL_1195846, EPI_ISL_1195856, EPI_ISL_1195857, EPI_ISL_1195864, EPI_ISL_1195866, EPI_ISL_1195871, EPI_ISL_1195880, EPI_ISL_1195887, EPI_ISL_1195888, EPI_ISL_1195897, EPI_ISL_1195898 | | | |
| see above | NYU Langone Health | Departments of Pathology and Medicine, New York University School of Medicine | Adriana Heguy, Dacia Dimartino, Emily Guzman, Christian Marier, Peter Meyn, Sitharam Ramaswami, Gael Westby, Paul Zappile, Yutong Zhang, Paolo Cotzia, Guiqing Wang |
| EPI_ISL_1196067 | Maryland Genomics, Institute for Genome Sciences, University of Maryland School of Medicine | Maryland Genomics, Institute for Genome Sciences, University of Maryland School of Medicine | Tallon, Luke J; Sadzewicz, Lisa D; Humphrys, Mike; Ott, Sandra; Roussey, Holly; Mehta, Aditya; Vavikolanu, Kranthi; Fraser, Claire M; Ravel, Jacques |
| EPI_ISL_1197131, EPI_ISL_1197134, EPI_ISL_1197135, EPI_ISL_1197136, EPI_ISL_1197137, EPI_ISL_1197138, EPI_ISL_1197141, EPI_ISL_1197144, EPI_ISL_1197147 | USC Clinical Lab | Los Angeles County PHL | P. Hemarajata et al. |
| EPI_ISL_1200526, EPI_ISL_1200527 | OCME Office Of Chief Medical Examiner | New York City Public Health Laboratory | Jade Wang, et al. |
| EPI_ISL_1200533 | DOHMH Corona | New York City Public Health Laboratory | Jade Wang, et al. |

| | | | |
|---|---|---|---|
| EPI_ISL_1200535 | DOHMH Jamaica | New York City Public Health Laboratory | Jade Wang, et al. |
| EPI_ISL_1200537 | DOHMH Riverside | New York City Public Health Laboratory | Jade Wang, et al. |
| EPI_ISL_1200542 | DOHMH Jamaica | New York City Public Health Laboratory | Jade Wang, et al. |
| EPI_ISL_1200543 | DOHMH Crown Heights | New York City Public Health Laboratory | Jade Wang, et al. |
| EPI_ISL_1200562 | Department of Homeless Services | New York City Public Health Laboratory | Jade Wang, et al. |
| EPI_ISL_1200563 | DOHMH Jamaica | New York City Public Health Laboratory | Jade Wang, et al. |
| EPI_ISL_1200564 | DOHMH Chelsea | New York City Public Health Laboratory | Jade Wang, et al. |
| EPI_ISL_1200565 | DOHMH Corona | New York City Public Health Laboratory | Jade Wang, et al. |
| EPI_ISL_1200566 | DOHMH PHL | New York City Public Health Laboratory | Jade Wang, et al. |
| EPI_ISL_1200567 | DOHMH Morrisania | New York City Public Health Laboratory | Jade Wang, et al. |
| EPI_ISL_1200568, EPI_ISL_1200569, EPI_ISL_1200570 | DOHMH Corona | New York City Public Health Laboratory | Jade Wang, et al. |
| EPI_ISL_1200571 | DOHMH Crown Heights | New York City Public Health Laboratory | Jade Wang, et al. |
| EPI_ISL_1200572, EPI_ISL_1200573 | DOHMH Jamaica | New York City Public Health Laboratory | Jade Wang, et al. |
| EPI_ISL_1200574 | DOHMH Chelsea | New York City Public Health Laboratory | Jade Wang, et al. |
| EPI_ISL_1200575, EPI_ISL_1200582 | DOHMH Morrisania | New York City Public Health Laboratory | Jade Wang, et al. |
| EPI_ISL_1200583 | DOHMH Jamaica | New York City Public Health Laboratory | Jade Wang, et al. |
| EPI_ISL_1200584 | DOHMH Chelsea | New York City Public Health Laboratory | Jade Wang, et al. |
| EPI_ISL_1200585 | DOHMH Jamaica | New York City Public Health Laboratory | Jade Wang, et al. |
| EPI_ISL_1200586, EPI_ISL_1200587 | DOHMH PHL | New York City Public Health Laboratory | Jade Wang, et al. |
| EPI_ISL_1200588, EPI_ISL_1200589 | DOHMH Morrisania | New York City Public Health Laboratory | Jade Wang, et al. |
| EPI_ISL_1200590 | DOHMH Jamaica | New York City Public Health Laboratory | Jade Wang, et al. |
| EPI_ISL_1200591, EPI_ISL_1200592, EPI_ISL_1200593 | DOHMH Corona | New York City Public Health Laboratory | Jade Wang, et al. |
| EPI_ISL_1200594 | DOHMH Central Harlem | New York City Public Health Laboratory | Jade Wang, et al. |
| EPI_ISL_1200595, EPI_ISL_1200596 | DOHMH Jamaica | New York City Public Health Laboratory | Jade Wang, et al. |
| EPI_ISL_1200597, EPI_ISL_1200598 | DOHMH Corona | New York City Public Health Laboratory | Jade Wang, et al. |
| EPI_ISL_1200599 | Department of Homeless Services | New York City Public Health Laboratory | Jade Wang, et al. |
| EPI_ISL_1200600 | DOHMH Morrisania | New York City Public Health Laboratory | Jade Wang, et al. |
| EPI_ISL_1200601, EPI_ISL_1200602 | DOHMH PHL | New York City Public Health Laboratory | Jade Wang, et al. |
| EPI_ISL_1200603 | DOHMH Corona | New York City Public Health Laboratory | Jade Wang, et al. |
| EPI_ISL_1200604 | DOHMH Morrisania | New York City Public Health Laboratory | Jade Wang, et al. |
| EPI_ISL_1200605 | DOHMH Crown Heights | New York City Public Health Laboratory | Jade Wang, et al. |
| EPI_ISL_1200606 | DOHMH Riverside | New York City Public Health Laboratory | Jade Wang, et al. |
| EPI_ISL_1200607, EPI_ISL_1200608, EPI_ISL_1200609, EPI_ISL_1200610 | OCME Office Of Chief Medical Examiner | New York City Public Health Laboratory | Jade Wang, et al. |
| EPI_ISL_1200920 | University of Michigan Clinical Microbiology Laboratory | Lauring Lab, University of Michigan, Department of Microbiology and Immunology | Valesano |
| EPI_ISL_1203819, EPI_ISL_1203826, EPI_ISL_1203827, EPI_ISL_1203828 | The Jackson Laboratory | The Jackson Laboratory | Lloyd M, Sanderson B, Srivastava A, Maurya R, Renzette N, Omerza G, Kelly K, Li L, Wei C L, Adams M |
| EPI_ISL_1209430, EPI_ISL_1209451, EPI_ISL_1209452 | Division of Emerging Infectious Diseases, Bureau of Infectious Diseases Diagnosis Control, Korea Disease Control and Prevention Agency | Division of Emerging Infectious Diseases, Bureau of Infectious Diseases Diagnosis Control, Korea Disease Control and Prevention Agency | Ae Kyung Park, Il-Hwan Kim, Heui Man Kim, Jeong-Min Kim, Chae Young Lee, Jin Sun No, Namjoo Lee, Sang Hee Woo, Eun-Jin Kim |
| EPI_ISL_1218741, EPI_ISL_1218814, EPI_ISL_1218935, EPI_ISL_1218971 | Florida Bureau of Public Health Laboratories | Florida Bureau of Public Health Laboratories | Sarah Schmedes, Jason Blanton |
| EPI_ISL_1220946, EPI_ISL_1220953, EPI_ISL_1221011, EPI_ISL_1221015, EPI_ISL_1221114, EPI_ISL_1221125, EPI_ISL_1221137, EPI_ISL_1221138, EPI_ISL_1221160, EPI_ISL_1221173, EPI_ISL_1221175, EPI_ISL_1221226, EPI_ISL_1221280, EPI_ISL_1221424, EPI_ISL_1221426, EPI_ISL_1221521, EPI_ISL_1221549, EPI_ISL_1221618, EPI_ISL_1221748, EPI_ISL_1221749, EPI_ISL_1221770, EPI_ISL_1221782, EPI_ISL_1221958, EPI_ISL_1221982, EPI_ISL_1222052, EPI_ISL_1222083, EPI_ISL_1222140, EPI_ISL_1222143, EPI_ISL_1222144, EPI_ISL_1222270, EPI_ISL_1222380, EPI_ISL_1222413 | | | |
| see above | Laboratory Corporation of America | Centers for Disease Control and Prevention Division of Viral Diseases, Pathogen Discovery | Peter W. Cook, Dakota Howard, Dhwani Batra, Ben L. Rambo-Martin, Minoo Agarwal, Eyad Almasri, Debbie Boles, Ayla Burns, Nuthawin Charoensri, Oren Cohen, Susan Countryman, Mary Ann Cristobal, Bobbi Croy, Suzanne Dale, Hrushikesh Deshmukh, Amanda Douglas, Vincent Drouillon, Marcia Eisenberg, Howard Engler, Rama Ghatti, Prashant Gupta, Susan Hicks, Jake Humphrey, Lax Iyer, Manoj Jain, Mohan Kolli, Brian Krueger, Tim Kuphal, Stanley Letovsky, Michael Levandoski, Craig Lukasik, Jonathan Meltzer, Brian Norvell, Mindy Nye, Scott Parker, Christos Petropoulos, John Pruitt, Steven Ragan, Scott Ryan, Mike Sapeta, Jana Schroth, Suresh Babu Selvaraju, Goran Stevovic, Amanda Suchanek, Andrea Throop, Lyndon Tilson, Thomas Urban, Joe Voshell, Kimberly Wagner, Jonathan Williams, Mary Williamson, Qian Zeng, Tricia Zwiefelhofer, Clinton R. Paden, Suxiang Tong, Duncan MacCannell |
| EPI_ISL_1224790 | M Health Fairview | Minnesota Department of Health, Public Health Laboratory | Alexandra Lorentz, Jacob Garfin, Matt Plumb, and Xiong Wang |
| EPI_ISL_1225608, EPI_ISL_1225609, EPI_ISL_1225668, EPI_ISL_1225670 | Yale Clinical Virology Lab | Grubaugh Lab - Yale School of Public Health | Joseph Fauver, Mallery Breban, Isabell Ott, Tara Alpert, Mary Petrone, Anderson Brito, Chantal Vogels, Annie Watkins, Chaney Kalinich, Marie L. Landry, Nathan Grubaugh |
| EPI_ISL_1225678, EPI_ISL_1225701, EPI_ISL_1225713 | Murphy Medical Association | Grubaugh Lab - Yale School of Public Health | Joseph Fauver, Mallery Breban, Isabell Ott, Tara Alpert, Mary Petrone, Anderson Brito, Chantal Vogels, Annie Watkins, Chaney Kalinich, Caleb Neal, Eva Laszlo, Steven Murphy, Nathan Grubaugh |
| EPI_ISL_1225729, EPI_ISL_1225730 | Yale Clinical Virology Lab | Grubaugh Lab - Yale School of Public Health | Joseph Fauver, Mallery Breban, Isabell Ott, Tara Alpert, Mary Petrone, Anderson Brito, Chantal Vogels, Annie Watkins, Chaney Kalinich, Marie L. Landry, Nathan Grubaugh |
| EPI_ISL_1226430 | NYC Pandemic Response Lab | Wadsworth Center, New York State Department of Health | Kirsten St. George, Daryl M. Lamson, Alexis Russel, Matthew Shudt, Melissa A Leisner, Jonathan Plitnick, Navjot Singh, John Kelly, Erasmus Schneider, Erica Lasek-Nesselquist |
| EPI_ISL_1226433 | MEMORIAL SLOAN KETTERING CANCER CENTER | Wadsworth Center, New York State Department of Health | Kirsten St. George, Daryl M. Lamson, Alexis Russel, Matthew Shudt, Melissa A Leisner, Jonathan Plitnick, Navjot Singh, John Kelly, Erasmus Schneider, Erica Lasek-Nesselquist |

| | | | |
|---|---|---|---|
| EPI_ISL_1226439 | Columbia University Irving Medical Center | Wadsworth Center, New York State Department of Health | Kirsten St. George, Daryl M. Lamson, Alexis Russel, Matthew Shudt, Melissa A Leisner, Jonathan Plitnick, Navjot Singh, John Kelly, Erasmus Schneider, Erica Lasek-Nesselquist |
| EPI_ISL_1226440 | SUNY UPSTATE MEDICAL UNIVERSITY | Wadsworth Center, New York State Department of Health | Kirsten St. George, Daryl M. Lamson, Alexis Russel, Matthew Shudt, Melissa A Leisner, Jonathan Plitnick, Navjot Singh, John Kelly, Erasmus Schneider, Erica Lasek-Nesselquist |
| EPI_ISL_1226456 | URMC LABS | Wadsworth Center, New York State Department of Health | Kirsten St. George, Daryl M. Lamson, Alexis Russel, Matthew Shudt, Melissa A Leisner, Jonathan Plitnick, Navjot Singh, John Kelly, Erasmus Schneider, Erica Lasek-Nesselquist |
| EPI_ISL_1226461 | MONTEFIORE MEDICAL CENTER LABORATORIES | Wadsworth Center, New York State Department of Health | Kirsten St. George, Daryl M. Lamson, Alexis Russel, Matthew Shudt, Melissa A Leisner, Jonathan Plitnick, Navjot Singh, John Kelly, Erasmus Schneider, Erica Lasek-Nesselquist |
| EPI_ISL_1226465 | NYC Pandemic Response Lab | Wadsworth Center, New York State Department of Health | Kirsten St. George, Daryl M. Lamson, Alexis Russel, Matthew Shudt, Melissa A Leisner, Jonathan Plitnick, Navjot Singh, John Kelly, Erasmus Schneider, Erica Lasek-Nesselquist |
| EPI_ISL_1226493 | MONTEFIORE MEDICAL CENTER LABORATORIES | Wadsworth Center, New York State Department of Health | Kirsten St. George, Daryl M. Lamson, Alexis Russel, Matthew Shudt, Melissa A Leisner, Jonathan Plitnick, Navjot Singh, John Kelly, Erasmus Schneider, Erica Lasek-Nesselquist |
| EPI_ISL_1226521 | TEMPUS LABS INC | Wadsworth Center, New York State Department of Health | Kirsten St. George, Daryl M. Lamson, Alexis Russel, Matthew Shudt, Melissa A Leisner, Jonathan Plitnick, Navjot Singh, John Kelly, Erasmus Schneider, Erica Lasek-Nesselquist |
| EPI_ISL_1226595, EPI_ISL_1226708 | MONTEFIORE MEDICAL CENTER LABORATORIES | Wadsworth Center, New York State Department of Health | Kirsten St. George, Daryl M. Lamson, Alexis Russel, Matthew Shudt, Melissa A Leisner, Jonathan Plitnick, Navjot Singh, John Kelly, Erasmus Schneider, Erica Lasek-Nesselquist |
| EPI_ISL_1226734 | URMC LABS | Wadsworth Center, New York State Department of Health | Kirsten St. George, Daryl M. Lamson, Alexis Russel, Matthew Shudt, Melissa A Leisner, Jonathan Plitnick, Navjot Singh, John Kelly, Erasmus Schneider, Erica Lasek-Nesselquist |
| EPI_ISL_1226735 | NYC Pandemic Response Lab | Wadsworth Center, New York State Department of Health | Kirsten St. George, Daryl M. Lamson, Alexis Russel, Matthew Shudt, Melissa A Leisner, Jonathan Plitnick, Navjot Singh, John Kelly, Erasmus Schneider, Erica Lasek-Nesselquist |
| EPI_ISL_1226737 | NORTHWELL HEALTH LABORATORIES | Wadsworth Center, New York State Department of Health | Kirsten St. George, Daryl M. Lamson, Alexis Russel, Matthew Shudt, Melissa A Leisner, Jonathan Plitnick, Navjot Singh, John Kelly, Erasmus Schneider, Erica Lasek-Nesselquist |
| EPI_ISL_1226738, EPI_ISL_1226739 | NYC Pandemic Response Lab | Wadsworth Center, New York State Department of Health | Kirsten St. George, Daryl M. Lamson, Alexis Russel, Matthew Shudt, Melissa A Leisner, Jonathan Plitnick, Navjot Singh, John Kelly, Erasmus Schneider, Erica Lasek-Nesselquist |
| EPI_ISL_1226740 | Wadsworth Center, New York State Department of Health | Wadsworth Center, New York State Department of Health | Kirsten St. George, Daryl M. Lamson, Alexis Russel, Matthew Shudt, Melissa A Leisner, Jonathan Plitnick, Navjot Singh, John Kelly, Erasmus Schneider, Erica Lasek-Nesselquist |
| EPI_ISL_1226741 | THE MARY IMOGENE BASSETT HOSPITAL | Wadsworth Center, New York State Department of Health | Kirsten St. George, Daryl M. Lamson, Alexis Russel, Matthew Shudt, Melissa A Leisner, Jonathan Plitnick, Navjot Singh, John Kelly, Erasmus Schneider, Erica Lasek-Nesselquist |
| EPI_ISL_1226742 | NORTHWELL HEALTH LABORATORIES | Wadsworth Center, New York State Department of Health | Kirsten St. George, Daryl M. Lamson, Alexis Russel, Matthew Shudt, Melissa A Leisner, Jonathan Plitnick, Navjot Singh, John Kelly, Erasmus Schneider, Erica Lasek-Nesselquist |
| EPI_ISL_1226743 | MEMORIAL SLOAN KETTERING CANCER CENTER | Wadsworth Center, New York State Department of Health | Kirsten St. George, Daryl M. Lamson, Alexis Russel, Matthew Shudt, Melissa A Leisner, Jonathan Plitnick, Navjot Singh, John Kelly, Erasmus Schneider, Erica Lasek-Nesselquist |
| EPI_ISL_1226744, EPI_ISL_1226745 | MONTEFIORE MEDICAL CENTER LABORATORIES | Wadsworth Center, New York State Department of Health | Kirsten St. George, Daryl M. Lamson, Alexis Russel, Matthew Shudt, Melissa A Leisner, Jonathan Plitnick, Navjot Singh, John Kelly, Erasmus Schneider, Erica Lasek-Nesselquist |
| EPI_ISL_1226746 | Masonic Care Community of NY | Wadsworth Center, New York State Department of Health | Kirsten St. George, Daryl M. Lamson, Alexis Russel, Matthew Shudt, Melissa A Leisner, Jonathan Plitnick, Navjot Singh, John Kelly, Erasmus Schneider, Erica Lasek-Nesselquist |
| EPI_ISL_1226747 | MEMORIAL SLOAN KETTERING CANCER CENTER | Wadsworth Center, New York State Department of Health | Kirsten St. George, Daryl M. Lamson, Alexis Russel, Matthew Shudt, Melissa A Leisner, Jonathan Plitnick, Navjot Singh, John Kelly, Erasmus Schneider, Erica Lasek-Nesselquist |
| EPI_ISL_1226748 | Masonic Care Community of NY | Wadsworth Center, New York State Department of Health | Kirsten St. George, Daryl M. Lamson, Alexis Russel, Matthew Shudt, Melissa A Leisner, Jonathan Plitnick, Navjot Singh, John Kelly, Erasmus Schneider, Erica Lasek-Nesselquist |
| EPI_ISL_1226749 | MEMORIAL SLOAN KETTERING CANCER CENTER | Wadsworth Center, New York State Department of Health | Kirsten St. George, Daryl M. Lamson, Alexis Russel, Matthew Shudt, Melissa A Leisner, Jonathan Plitnick, Navjot Singh, John Kelly, Erasmus Schneider, Erica Lasek-Nesselquist |
| EPI_ISL_1226750 | URMC LABS | Wadsworth Center, New York State Department of Health | Kirsten St. George, Daryl M. Lamson, Alexis Russel, Matthew Shudt, Melissa A Leisner, Jonathan Plitnick, Navjot Singh, John Kelly, Erasmus Schneider, Erica Lasek-Nesselquist |
| EPI_ISL_1226751, EPI_ISL_1226752 | Wadsworth Center, New York State Department of Health | Wadsworth Center, New York State Department of Health | Kirsten St. George, Daryl M. Lamson, Alexis Russel, Matthew Shudt, Melissa A Leisner, Jonathan Plitnick, Navjot Singh, John Kelly, Erasmus Schneider, Erica Lasek-Nesselquist |
| EPI_ISL_1226753 | SUNY UPSTATE MEDICAL UNIVERSITY | Wadsworth Center, New York State Department of Health | Kirsten St. George, Daryl M. Lamson, Alexis Russel, Matthew Shudt, Melissa A Leisner, Jonathan Plitnick, Navjot Singh, John Kelly, Erasmus Schneider, Erica Lasek-Nesselquist |
| EPI_ISL_1226754 | KALEIDA CENTER FOR LABORATORY MEDICINE | Wadsworth Center, New York State Department of Health | Kirsten St. George, Daryl M. Lamson, Alexis Russel, Matthew Shudt, Melissa A Leisner, Jonathan Plitnick, Navjot Singh, John Kelly, Erasmus Schneider, Erica Lasek-Nesselquist |
| EPI_ISL_1226755, EPI_ISL_1226756 | MONTEFIORE MEDICAL CENTER LABORATORIES | Wadsworth Center, New York State Department of Health | Kirsten St. George, Daryl M. Lamson, Alexis Russel, Matthew Shudt, Melissa A Leisner, Jonathan Plitnick, Navjot Singh, John Kelly, Erasmus Schneider, Erica Lasek-Nesselquist |
| EPI_ISL_1226757 | WESTCHESTER MEDICAL CENTER | Wadsworth Center, New York State Department of Health | Kirsten St. George, Daryl M. Lamson, Alexis Russel, Matthew Shudt, Melissa A Leisner, Jonathan Plitnick, Navjot Singh, John Kelly, Erasmus Schneider, Erica Lasek-Nesselquist |
| EPI_ISL_1226758, EPI_ISL_1226759 | MONTEFIORE MEDICAL CENTER LABORATORIES | Wadsworth Center, New York State Department of Health | Kirsten St. George, Daryl M. Lamson, Alexis Russel, Matthew Shudt, Melissa A Leisner, Jonathan Plitnick, Navjot Singh, John Kelly, Erasmus Schneider, Erica Lasek-Nesselquist |
| EPI_ISL_1226760, EPI_ISL_1226761, EPI_ISL_1226762 | URMC LABS | Wadsworth Center, New York State Department of Health | Kirsten St. George, Daryl M. Lamson, Alexis Russel, Matthew Shudt, Melissa A Leisner, Jonathan Plitnick, Navjot Singh, John Kelly, Erasmus Schneider, Erica Lasek-Nesselquist |
| EPI_ISL_1226763 | BOSTON HEART DIAGNOSTICS CORP | Wadsworth Center, New York State Department of Health | Kirsten St. George, Daryl M. Lamson, Alexis Russel, Matthew Shudt, Melissa A Leisner, Jonathan Plitnick, Navjot Singh, John Kelly, Erasmus Schneider, Erica Lasek-Nesselquist |
| EPI_ISL_1226764 | ALBANY MEDICAL CENTER | Wadsworth Center, New York State Department of Health | Kirsten St. George, Daryl M. Lamson, Alexis Russel, Matthew Shudt, Melissa A Leisner, Jonathan Plitnick, Navjot Singh, John Kelly, Erasmus Schneider, Erica Lasek-Nesselquist |
| EPI_ISL_1226765, EPI_ISL_1226766, EPI_ISL_1226767, EPI_ISL_1226768, EPI_ISL_1226769, EPI_ISL_1226770 | NYC Pandemic Response Lab | Wadsworth Center, New York State Department of Health | Kirsten St. George, Daryl M. Lamson, Alexis Russel, Matthew Shudt, Melissa A Leisner, Jonathan Plitnick, Navjot Singh, John Kelly, Erasmus Schneider, Erica Lasek-Nesselquist |
| EPI_ISL_1226771, EPI_ISL_1226772 | MEMORIAL SLOAN KETTERING CANCER CENTER | Wadsworth Center, New York State Department of Health | Kirsten St. George, Daryl M. Lamson, Alexis Russel, Matthew Shudt, Melissa A Leisner, Jonathan Plitnick, Navjot Singh, John Kelly, Erasmus Schneider, Erica Lasek-Nesselquist |
| EPI_ISL_1226773 | SUNY UPSTATE MEDICAL UNIVERSITY | Wadsworth Center, New York State Department of Health | Kirsten St. George, Daryl M. Lamson, Alexis Russel, Matthew Shudt, Melissa A Leisner, Jonathan Plitnick, Navjot Singh, John Kelly, Erasmus Schneider, Erica Lasek-Nesselquist |
| EPI_ISL_1226774, EPI_ISL_1226775 | ALBANY MEDICAL CENTER | Wadsworth Center, New York State Department of Health | Kirsten St. George, Daryl M. Lamson, Alexis Russel, Matthew Shudt, Melissa A Leisner, Jonathan Plitnick, Navjot Singh, John Kelly, Erasmus Schneider, Erica Lasek-Nesselquist |

| | | | |
|---|---|---|---|
| EPI_ISL_1226776 | SUNY UPSTATE MEDICAL UNIVERSITY | Wadsworth Center, New York State Department of Health | Kirsten St. George, Daryl M. Lamson, Alexis Russel, Matthew Shudt, Melissa A Leisner, Jonathan Plitnick, Navjot Singh, John Kelly, Erasmus Schneider, Erica Lasek-Nesselquist |
| EPI_ISL_1226777, EPI_ISL_1226778, EPI_ISL_1226779, EPI_ISL_1226780, EPI_ISL_1226781 | NYC Pandemic Response Lab | Wadsworth Center, New York State Department of Health | Kirsten St. George, Daryl M. Lamson, Alexis Russel, Matthew Shudt, Melissa A Leisner, Jonathan Plitnick, Navjot Singh, John Kelly, Erasmus Schneider, Erica Lasek-Nesselquist |
| EPI_ISL_1226782 | MEMORIAL SLOAN KETTERING CANCER CENTER | Wadsworth Center, New York State Department of Health | Kirsten St. George, Daryl M. Lamson, Alexis Russel, Matthew Shudt, Melissa A Leisner, Jonathan Plitnick, Navjot Singh, John Kelly, Erasmus Schneider, Erica Lasek-Nesselquist |
| EPI_ISL_1226783 | MONTEFIORE MEDICAL CENTER LABORATORIES | Wadsworth Center, New York State Department of Health | Kirsten St. George, Daryl M. Lamson, Alexis Russel, Matthew Shudt, Melissa A Leisner, Jonathan Plitnick, Navjot Singh, John Kelly, Erasmus Schneider, Erica Lasek-Nesselquist |
| EPI_ISL_1226784 | MEMORIAL SLOAN KETTERING CANCER CENTER | Wadsworth Center, New York State Department of Health | Kirsten St. George, Daryl M. Lamson, Alexis Russel, Matthew Shudt, Melissa A Leisner, Jonathan Plitnick, Navjot Singh, John Kelly, Erasmus Schneider, Erica Lasek-Nesselquist |
| EPI_ISL_1226785 | NYC Pandemic Response Lab | Wadsworth Center, New York State Department of Health | Kirsten St. George, Daryl M. Lamson, Alexis Russel, Matthew Shudt, Melissa A Leisner, Jonathan Plitnick, Navjot Singh, John Kelly, Erasmus Schneider, Erica Lasek-Nesselquist |
| EPI_ISL_1226786 | MONTEFIORE MEDICAL CENTER LABORATORIES | Wadsworth Center, New York State Department of Health | Kirsten St. George, Daryl M. Lamson, Alexis Russel, Matthew Shudt, Melissa A Leisner, Jonathan Plitnick, Navjot Singh, John Kelly, Erasmus Schneider, Erica Lasek-Nesselquist |
| EPI_ISL_1226787, EPI_ISL_1226788 | NYC Pandemic Response Lab | Wadsworth Center, New York State Department of Health | Kirsten St. George, Daryl M. Lamson, Alexis Russel, Matthew Shudt, Melissa A Leisner, Jonathan Plitnick, Navjot Singh, John Kelly, Erasmus Schneider, Erica Lasek-Nesselquist |
| EPI_ISL_1226789 | Columbia University Irving Medical Center | Wadsworth Center, New York State Department of Health | Kirsten St. George, Daryl M. Lamson, Alexis Russel, Matthew Shudt, Melissa A Leisner, Jonathan Plitnick, Navjot Singh, John Kelly, Erasmus Schneider, Erica Lasek-Nesselquist |
| EPI_ISL_1226790, EPI_ISL_1226791, EPI_ISL_1226792 | ALBANY MEDICAL CENTER | Wadsworth Center, New York State Department of Health | Kirsten St. George, Daryl M. Lamson, Alexis Russel, Matthew Shudt, Melissa A Leisner, Jonathan Plitnick, Navjot Singh, John Kelly, Erasmus Schneider, Erica Lasek-Nesselquist |
| EPI_ISL_1226793 | TEMPUS LABS INC | Wadsworth Center, New York State Department of Health | Kirsten St. George, Daryl M. Lamson, Alexis Russel, Matthew Shudt, Melissa A Leisner, Jonathan Plitnick, Navjot Singh, John Kelly, Erasmus Schneider, Erica Lasek-Nesselquist |
| EPI_ISL_1226794 | NYC Pandemic Response Lab | Wadsworth Center, New York State Department of Health | Kirsten St. George, Daryl M. Lamson, Alexis Russel, Matthew Shudt, Melissa A Leisner, Jonathan Plitnick, Navjot Singh, John Kelly, Erasmus Schneider, Erica Lasek-Nesselquist |
| EPI_ISL_1226795 | WESTCHESTER MEDICAL CENTER | Wadsworth Center, New York State Department of Health | Kirsten St. George, Daryl M. Lamson, Alexis Russel, Matthew Shudt, Melissa A Leisner, Jonathan Plitnick, Navjot Singh, John Kelly, Erasmus Schneider, Erica Lasek-Nesselquist |
| EPI_ISL_1226796 | MONTEFIORE MEDICAL CENTER LABORATORIES | Wadsworth Center, New York State Department of Health | Kirsten St. George, Daryl M. Lamson, Alexis Russel, Matthew Shudt, Melissa A Leisner, Jonathan Plitnick, Navjot Singh, John Kelly, Erasmus Schneider, Erica Lasek-Nesselquist |
| EPI_ISL_1226797 | ALBANY MEDICAL CENTER | Wadsworth Center, New York State Department of Health | Kirsten St. George, Daryl M. Lamson, Alexis Russel, Matthew Shudt, Melissa A Leisner, Jonathan Plitnick, Navjot Singh, John Kelly, Erasmus Schneider, Erica Lasek-Nesselquist |
| EPI_ISL_1226798, EPI_ISL_1226800, EPI_ISL_1226801 | NYC Pandemic Response Lab | Wadsworth Center, New York State Department of Health | Kirsten St. George, Daryl M. Lamson, Alexis Russel, Matthew Shudt, Melissa A Leisner, Jonathan Plitnick, Navjot Singh, John Kelly, Erasmus Schneider, Erica Lasek-Nesselquist |
| EPI_ISL_1226802, EPI_ISL_1226803 | MEMORIAL SLOAN KETTERING CANCER CENTER | Wadsworth Center, New York State Department of Health | Kirsten St. George, Daryl M. Lamson, Alexis Russel, Matthew Shudt, Melissa A Leisner, Jonathan Plitnick, Navjot Singh, John Kelly, Erasmus Schneider, Erica Lasek-Nesselquist |
| EPI_ISL_1226804, EPI_ISL_1226805, EPI_ISL_1226806, EPI_ISL_1226807 | MONTEFIORE MEDICAL CENTER LABORATORIES | Wadsworth Center, New York State Department of Health | Kirsten St. George, Daryl M. Lamson, Alexis Russel, Matthew Shudt, Melissa A Leisner, Jonathan Plitnick, Navjot Singh, John Kelly, Erasmus Schneider, Erica Lasek-Nesselquist |
| EPI_ISL_1226808 | NYC Pandemic Response Lab | Wadsworth Center, New York State Department of Health | Kirsten St. George, Daryl M. Lamson, Alexis Russel, Matthew Shudt, Melissa A Leisner, Jonathan Plitnick, Navjot Singh, John Kelly, Erasmus Schneider, Erica Lasek-Nesselquist |
| EPI_ISL_1226809 | WESTCHESTER MEDICAL CENTER | Wadsworth Center, New York State Department of Health | Kirsten St. George, Daryl M. Lamson, Alexis Russel, Matthew Shudt, Melissa A Leisner, Jonathan Plitnick, Navjot Singh, John Kelly, Erasmus Schneider, Erica Lasek-Nesselquist |
| EPI_ISL_1226810 | MONTEFIORE MEDICAL CENTER LABORATORIES | Wadsworth Center, New York State Department of Health | Kirsten St. George, Daryl M. Lamson, Alexis Russel, Matthew Shudt, Melissa A Leisner, Jonathan Plitnick, Navjot Singh, John Kelly, Erasmus Schneider, Erica Lasek-Nesselquist |
| EPI_ISL_1226811, EPI_ISL_1226812 | URMC LABS | Wadsworth Center, New York State Department of Health | Kirsten St. George, Daryl M. Lamson, Alexis Russel, Matthew Shudt, Melissa A Leisner, Jonathan Plitnick, Navjot Singh, John Kelly, Erasmus Schneider, Erica Lasek-Nesselquist |
| EPI_ISL_1226813 | SUNY UPSTATE MEDICAL UNIVERSITY | Wadsworth Center, New York State Department of Health | Kirsten St. George, Daryl M. Lamson, Alexis Russel, Matthew Shudt, Melissa A Leisner, Jonathan Plitnick, Navjot Singh, John Kelly, Erasmus Schneider, Erica Lasek-Nesselquist |
| EPI_ISL_1226814 | NYC Pandemic Response Lab | Wadsworth Center, New York State Department of Health | Kirsten St. George, Daryl M. Lamson, Alexis Russel, Matthew Shudt, Melissa A Leisner, Jonathan Plitnick, Navjot Singh, John Kelly, Erasmus Schneider, Erica Lasek-Nesselquist |
| EPI_ISL_1226815 | TEMPUS LABS INC | Wadsworth Center, New York State Department of Health | Kirsten St. George, Daryl M. Lamson, Alexis Russel, Matthew Shudt, Melissa A Leisner, Jonathan Plitnick, Navjot Singh, John Kelly, Erasmus Schneider, Erica Lasek-Nesselquist |
| EPI_ISL_1226816 | WESTCHESTER MEDICAL CENTER | Wadsworth Center, New York State Department of Health | Kirsten St. George, Daryl M. Lamson, Alexis Russel, Matthew Shudt, Melissa A Leisner, Jonathan Plitnick, Navjot Singh, John Kelly, Erasmus Schneider, Erica Lasek-Nesselquist |
| EPI_ISL_1226817 | MONTEFIORE MEDICAL CENTER LABORATORIES | Wadsworth Center, New York State Department of Health | Kirsten St. George, Daryl M. Lamson, Alexis Russel, Matthew Shudt, Melissa A Leisner, Jonathan Plitnick, Navjot Singh, John Kelly, Erasmus Schneider, Erica Lasek-Nesselquist |
| EPI_ISL_1226818 | NYC Pandemic Response Lab | Wadsworth Center, New York State Department of Health | Kirsten St. George, Daryl M. Lamson, Alexis Russel, Matthew Shudt, Melissa A Leisner, Jonathan Plitnick, Navjot Singh, John Kelly, Erasmus Schneider, Erica Lasek-Nesselquist |
| EPI_ISL_1226819 | THE MARY IMOGENE BASSETT HOSPITAL | Wadsworth Center, New York State Department of Health | Kirsten St. George, Daryl M. Lamson, Alexis Russel, Matthew Shudt, Melissa A Leisner, Jonathan Plitnick, Navjot Singh, John Kelly, Erasmus Schneider, Erica Lasek-Nesselquist |
| EPI_ISL_1226820 | MEMORIAL SLOAN KETTERING CANCER CENTER | Wadsworth Center, New York State Department of Health | Kirsten St. George, Daryl M. Lamson, Alexis Russel, Matthew Shudt, Melissa A Leisner, Jonathan Plitnick, Navjot Singh, John Kelly, Erasmus Schneider, Erica Lasek-Nesselquist |
| EPI_ISL_1226821 | TEMPUS LABS INC | Wadsworth Center, New York State Department of Health | Kirsten St. George, Daryl M. Lamson, Alexis Russel, Matthew Shudt, Melissa A Leisner, Jonathan Plitnick, Navjot Singh, John Kelly, Erasmus Schneider, Erica Lasek-Nesselquist |
| EPI_ISL_1226822 | WESTCHESTER MEDICAL CENTER | Wadsworth Center, New York State Department of Health | Kirsten St. George, Daryl M. Lamson, Alexis Russel, Matthew Shudt, Melissa A Leisner, Jonathan Plitnick, Navjot Singh, John Kelly, Erasmus Schneider, Erica Lasek-Nesselquist |
| EPI_ISL_1226823, EPI_ISL_1226824 | MEMORIAL SLOAN KETTERING CANCER CENTER | Wadsworth Center, New York State Department of Health | Kirsten St. George, Daryl M. Lamson, Alexis Russel, Matthew Shudt, Melissa A Leisner, Jonathan Plitnick, Navjot Singh, John Kelly, Erasmus Schneider, Erica Lasek-Nesselquist |
| EPI_ISL_1226825, EPI_ISL_1226826 | Wadsworth Center, New York State Department of Health | Wadsworth Center, New York State Department of Health | Kirsten St. George, Daryl M. Lamson, Alexis Russel, Matthew Shudt, Melissa A Leisner, Jonathan Plitnick, Navjot Singh, John Kelly, Erasmus Schneider, Erica Lasek-Nesselquist |
| EPI_ISL_1226827, EPI_ISL_1226828 | MEMORIAL SLOAN KETTERING CANCER CENTER | Wadsworth Center, New York State Department of Health | Kirsten St. George, Daryl M. Lamson, Alexis Russel, Matthew Shudt, Melissa A Leisner, Jonathan Plitnick, Navjot Singh, John Kelly, Erasmus Schneider, |

| | | | |
|---|---|---|---|
| | | | Erica Lasek-Nesselquist |
| EPI_ISL_1226829, EPI_ISL_1226830 | MONTEFIORE MEDICAL CENTER LABORATORIES | Wadsworth Center, New York State Department of Health | Kirsten St. George, Daryl M. Lamson, Alexis Russel, Matthew Shudt, Melissa A Leisner, Jonathan Plitnick, Navjot Singh, John Kelly, Erasmus Schneider, Erica Lasek-Nesselquist |
| EPI_ISL_1226831 | ALBANY MEDICAL CENTER | Wadsworth Center, New York State Department of Health | Kirsten St. George, Daryl M. Lamson, Alexis Russel, Matthew Shudt, Melissa A Leisner, Jonathan Plitnick, Navjot Singh, John Kelly, Erasmus Schneider, Erica Lasek-Nesselquist |
| EPI_ISL_1226832 | MONTEFIORE MEDICAL CENTER LABORATORIES | Wadsworth Center, New York State Department of Health | Kirsten St. George, Daryl M. Lamson, Alexis Russel, Matthew Shudt, Melissa A Leisner, Jonathan Plitnick, Navjot Singh, John Kelly, Erasmus Schneider, Erica Lasek-Nesselquist |
| EPI_ISL_1226833 | NORTH SHORE UNIVERSITY HOSPITAL | Wadsworth Center, New York State Department of Health | Kirsten St. George, Daryl M. Lamson, Alexis Russel, Matthew Shudt, Melissa A Leisner, Jonathan Plitnick, Navjot Singh, John Kelly, Erasmus Schneider, Erica Lasek-Nesselquist |
| EPI_ISL_1226834 | Columbia University Irving Medical Center | Wadsworth Center, New York State Department of Health | Kirsten St. George, Daryl M. Lamson, Alexis Russel, Matthew Shudt, Melissa A Leisner, Jonathan Plitnick, Navjot Singh, John Kelly, Erasmus Schneider, Erica Lasek-Nesselquist |
| EPI_ISL_1226835, EPI_ISL_1226836 | NYC Pandemic Response Lab | Wadsworth Center, New York State Department of Health | Kirsten St. George, Daryl M. Lamson, Alexis Russel, Matthew Shudt, Melissa A Leisner, Jonathan Plitnick, Navjot Singh, John Kelly, Erasmus Schneider, Erica Lasek-Nesselquist |
| EPI_ISL_1226837, EPI_ISL_1226838 | MONTEFIORE MEDICAL CENTER LABORATORIES | Wadsworth Center, New York State Department of Health | Kirsten St. George, Daryl M. Lamson, Alexis Russel, Matthew Shudt, Melissa A Leisner, Jonathan Plitnick, Navjot Singh, John Kelly, Erasmus Schneider, Erica Lasek-Nesselquist |
| EPI_ISL_1226839, EPI_ISL_1226840, EPI_ISL_1226841 | NYC Pandemic Response Lab | Wadsworth Center, New York State Department of Health | Kirsten St. George, Daryl M. Lamson, Alexis Russel, Matthew Shudt, Melissa A Leisner, Jonathan Plitnick, Navjot Singh, John Kelly, Erasmus Schneider, Erica Lasek-Nesselquist |
| EPI_ISL_1227029 | BIO-REFERENCE LABORATORIES | Wadsworth Center, New York State Department of Health | Kirsten St. George, Daryl M. Lamson, Alexis Russel, Matthew Shudt, Melissa A Leisner, Jonathan Plitnick, Navjot Singh, John Kelly, Erasmus Schneider, Erica Lasek-Nesselquist |
| EPI_ISL_1227030 | GLENS FALLS HOSPITAL LABORATORY | Wadsworth Center, New York State Department of Health | Kirsten St. George, Daryl M. Lamson, Alexis Russel, Matthew Shudt, Melissa A Leisner, Jonathan Plitnick, Navjot Singh, John Kelly, Erasmus Schneider, Erica Lasek-Nesselquist |
| EPI_ISL_1227031 | NYC Pandemic Response Lab | Wadsworth Center, New York State Department of Health | Kirsten St. George, Daryl M. Lamson, Alexis Russel, Matthew Shudt, Melissa A Leisner, Jonathan Plitnick, Navjot Singh, John Kelly, Erasmus Schneider, Erica Lasek-Nesselquist |
| EPI_ISL_1227032 | BIO-REFERENCE LABORATORIES | Wadsworth Center, New York State Department of Health | Kirsten St. George, Daryl M. Lamson, Alexis Russel, Matthew Shudt, Melissa A Leisner, Jonathan Plitnick, Navjot Singh, John Kelly, Erasmus Schneider, Erica Lasek-Nesselquist |
| EPI_ISL_1227033 | WESTCHESTER MEDICAL CENTER | Wadsworth Center, New York State Department of Health | Kirsten St. George, Daryl M. Lamson, Alexis Russel, Matthew Shudt, Melissa A Leisner, Jonathan Plitnick, Navjot Singh, John Kelly, Erasmus Schneider, Erica Lasek-Nesselquist |
| EPI_ISL_1227034 | Columbia University Irving Medical Center | Wadsworth Center, New York State Department of Health | Kirsten St. George, Daryl M. Lamson, Alexis Russel, Matthew Shudt, Melissa A Leisner, Jonathan Plitnick, Navjot Singh, John Kelly, Erasmus Schneider, Erica Lasek-Nesselquist |
| EPI_ISL_1227035, EPI_ISL_1227036, EPI_ISL_1227037, EPI_ISL_1227038 | MEMORIAL SLOAN KETTERING CANCER CENTER | Wadsworth Center, New York State Department of Health | Kirsten St. George, Daryl M. Lamson, Alexis Russel, Matthew Shudt, Melissa A Leisner, Jonathan Plitnick, Navjot Singh, John Kelly, Erasmus Schneider, Erica Lasek-Nesselquist |
| EPI_ISL_1227039 | NORTHWELL HEALTH LABORATORIES | Wadsworth Center, New York State Department of Health | Kirsten St. George, Daryl M. Lamson, Alexis Russel, Matthew Shudt, Melissa A Leisner, Jonathan Plitnick, Navjot Singh, John Kelly, Erasmus Schneider, Erica Lasek-Nesselquist |
| EPI_ISL_1227040 | MEMORIAL SLOAN KETTERING CANCER CENTER | Wadsworth Center, New York State Department of Health | Kirsten St. George, Daryl M. Lamson, Alexis Russel, Matthew Shudt, Melissa A Leisner, Jonathan Plitnick, Navjot Singh, John Kelly, Erasmus Schneider, Erica Lasek-Nesselquist |
| EPI_ISL_1227041 | MONTEFIORE MEDICAL CENTER LABORATORIES | Wadsworth Center, New York State Department of Health | Kirsten St. George, Daryl M. Lamson, Alexis Russel, Matthew Shudt, Melissa A Leisner, Jonathan Plitnick, Navjot Singh, John Kelly, Erasmus Schneider, Erica Lasek-Nesselquist |
| EPI_ISL_1227042 | Masonic Care Community of NY | Wadsworth Center, New York State Department of Health | Kirsten St. George, Daryl M. Lamson, Alexis Russel, Matthew Shudt, Melissa A Leisner, Jonathan Plitnick, Navjot Singh, John Kelly, Erasmus Schneider, Erica Lasek-Nesselquist |
| EPI_ISL_1227043 | MEMORIAL SLOAN KETTERING CANCER CENTER | Wadsworth Center, New York State Department of Health | Kirsten St. George, Daryl M. Lamson, Alexis Russel, Matthew Shudt, Melissa A Leisner, Jonathan Plitnick, Navjot Singh, John Kelly, Erasmus Schneider, Erica Lasek-Nesselquist |
| EPI_ISL_1227044, EPI_ISL_1227045, EPI_ISL_1227046, EPI_ISL_1227047, EPI_ISL_1227048, EPI_ISL_1227049 | URMC LABS | Wadsworth Center, New York State Department of Health | Kirsten St. George, Daryl M. Lamson, Alexis Russel, Matthew Shudt, Melissa A Leisner, Jonathan Plitnick, Navjot Singh, John Kelly, Erasmus Schneider, Erica Lasek-Nesselquist |
| EPI_ISL_1227050, EPI_ISL_1227051, EPI_ISL_1227052, EPI_ISL_1227053, EPI_ISL_1227054 | Wadsworth Center, New York State Department of Health | Wadsworth Center, New York State Department of Health | Kirsten St. George, Daryl M. Lamson, Alexis Russel, Matthew Shudt, Melissa A Leisner, Jonathan Plitnick, Navjot Singh, John Kelly, Erasmus Schneider, Erica Lasek-Nesselquist |
| EPI_ISL_1227055 | SUNY UPSTATE MEDICAL UNIVERSITY | Wadsworth Center, New York State Department of Health | Kirsten St. George, Daryl M. Lamson, Alexis Russel, Matthew Shudt, Melissa A Leisner, Jonathan Plitnick, Navjot Singh, John Kelly, Erasmus Schneider, Erica Lasek-Nesselquist |
| EPI_ISL_1227056, EPI_ISL_1227057, EPI_ISL_1227058 | KALEIDA CENTER FOR LABORATORY MEDICINE | Wadsworth Center, New York State Department of Health | Kirsten St. George, Daryl M. Lamson, Alexis Russel, Matthew Shudt, Melissa A Leisner, Jonathan Plitnick, Navjot Singh, John Kelly, Erasmus Schneider, Erica Lasek-Nesselquist |
| EPI_ISL_1227059 | New York Presbyterian Hospital | Wadsworth Center, New York State Department of Health | Kirsten St. George, Daryl M. Lamson, Alexis Russel, Matthew Shudt, Melissa A Leisner, Jonathan Plitnick, Navjot Singh, John Kelly, Erasmus Schneider, Erica Lasek-Nesselquist |
| EPI_ISL_1227060 | WESTCHESTER MEDICAL CENTER | Wadsworth Center, New York State Department of Health | Kirsten St. George, Daryl M. Lamson, Alexis Russel, Matthew Shudt, Melissa A Leisner, Jonathan Plitnick, Navjot Singh, John Kelly, Erasmus Schneider, Erica Lasek-Nesselquist |
| EPI_ISL_1227061, EPI_ISL_1227062, EPI_ISL_1227063, EPI_ISL_1227064, EPI_ISL_1227065, EPI_ISL_1227067, EPI_ISL_1227068, EPI_ISL_1227069 | MONTEFIORE MEDICAL CENTER LABORATORIES | Wadsworth Center, New York State Department of Health | Kirsten St. George, Daryl M. Lamson, Alexis Russel, Matthew Shudt, Melissa A Leisner, Jonathan Plitnick, Navjot Singh, John Kelly, Erasmus Schneider, Erica Lasek-Nesselquist |
| EPI_ISL_1227070, EPI_ISL_1227071 | URMC LABS | Wadsworth Center, New York State Department of Health | Kirsten St. George, Daryl M. Lamson, Alexis Russel, Matthew Shudt, Melissa A Leisner, Jonathan Plitnick, Navjot Singh, John Kelly, Erasmus Schneider, Erica Lasek-Nesselquist |
| EPI_ISL_1227072 | SARATOGA HOSPITAL LABORATORY | Wadsworth Center, New York State Department of Health | Kirsten St. George, Daryl M. Lamson, Alexis Russel, Matthew Shudt, Melissa A Leisner, Jonathan Plitnick, Navjot Singh, John Kelly, Erasmus Schneider, Erica Lasek-Nesselquist |
| EPI_ISL_1227073, EPI_ISL_1227074, EPI_ISL_1227075, EPI_ISL_1227076, EPI_ISL_1227077, EPI_ISL_1227078, EPI_ISL_1227079, EPI_ISL_1227080, EPI_ISL_1227081, EPI_ISL_1227082, EPI_ISL_1227083, EPI_ISL_1227084, EPI_ISL_1227085, EPI_ISL_1227086 | | | |
| see above | NYC Pandemic Response Lab | Wadsworth Center, New York State Department of Health | Kirsten St. George, Daryl M. Lamson, Alexis Russel, Matthew Shudt, Melissa A Leisner, Jonathan Plitnick, Navjot Singh, John Kelly, Erasmus Schneider, Erica Lasek-Nesselquist |
| EPI_ISL_1227087 | ALBANY MEDICAL CENTER | Wadsworth Center, New York State Department of Health | Kirsten St. George, Daryl M. Lamson, Alexis Russel, Matthew Shudt, Melissa A Leisner, Jonathan Plitnick, Navjot Singh, John Kelly, Erasmus Schneider, Erica Lasek-Nesselquist |
| EPI_ISL_1227088, EPI_ISL_1227089, EPI_ISL_1227090, EPI_ISL_1227091, EPI_ISL_1227092, EPI_ISL_1227093, | MEMORIAL SLOAN KETTERING CANCER CENTER | Wadsworth Center, New York State Department of Health | Kirsten St. George, Daryl M. Lamson, Alexis Russel, Matthew Shudt, Melissa A Leisner, Jonathan Plitnick, Navjot Singh, John Kelly, Erasmus Schneider, Erica Lasek-Nesselquist |

| | | | |
|---|---|---|---|
| EPI_ISL_1227094 | | | |
| EPI_ISL_1227095 | SUNY UPSTATE MEDICAL UNIVERSITY | Wadsworth Center, New York State Department of Health | Kirsten St. George, Daryl M. Lamson, Alexis Russel, Matthew Shudt, Melissa A Leisner, Jonathan Plitnick, Navjot Singh, John Kelly, Erasmus Schneider, Erica Lasek-Nesselquist |
| EPI_ISL_1227096 | URMC LABS | Wadsworth Center, New York State Department of Health | Kirsten St. George, Daryl M. Lamson, Alexis Russel, Matthew Shudt, Melissa A Leisner, Jonathan Plitnick, Navjot Singh, John Kelly, Erasmus Schneider, Erica Lasek-Nesselquist |
| EPI_ISL_1227097, EPI_ISL_1227098, EPI_ISL_1227099 | NYC Pandemic Response Lab | Wadsworth Center, New York State Department of Health | Kirsten St. George, Daryl M. Lamson, Alexis Russel, Matthew Shudt, Melissa A Leisner, Jonathan Plitnick, Navjot Singh, John Kelly, Erasmus Schneider, Erica Lasek-Nesselquist |
| EPI_ISL_1227100, EPI_ISL_1227101, EPI_ISL_1227102 | WESTCHESTER MEDICAL CENTER | Wadsworth Center, New York State Department of Health | Kirsten St. George, Daryl M. Lamson, Alexis Russel, Matthew Shudt, Melissa A Leisner, Jonathan Plitnick, Navjot Singh, John Kelly, Erasmus Schneider, Erica Lasek-Nesselquist |
| EPI_ISL_1227103, EPI_ISL_1227104 | ALBANY MEDICAL CENTER | Wadsworth Center, New York State Department of Health | Kirsten St. George, Daryl M. Lamson, Alexis Russel, Matthew Shudt, Melissa A Leisner, Jonathan Plitnick, Navjot Singh, John Kelly, Erasmus Schneider, Erica Lasek-Nesselquist |
| EPI_ISL_1227105, EPI_ISL_1227106 | KALEIDA CENTER FOR LABORATORY MEDICINE | Wadsworth Center, New York State Department of Health | Kirsten St. George, Daryl M. Lamson, Alexis Russel, Matthew Shudt, Melissa A Leisner, Jonathan Plitnick, Navjot Singh, John Kelly, Erasmus Schneider, Erica Lasek-Nesselquist |
| EPI_ISL_1227107 | SUNY UPSTATE MEDICAL UNIVERSITY | Wadsworth Center, New York State Department of Health | Kirsten St. George, Daryl M. Lamson, Alexis Russel, Matthew Shudt, Melissa A Leisner, Jonathan Plitnick, Navjot Singh, John Kelly, Erasmus Schneider, Erica Lasek-Nesselquist |
| EPI_ISL_1227108, EPI_ISL_1227109 | URMC LABS | Wadsworth Center, New York State Department of Health | Kirsten St. George, Daryl M. Lamson, Alexis Russel, Matthew Shudt, Melissa A Leisner, Jonathan Plitnick, Navjot Singh, John Kelly, Erasmus Schneider, Erica Lasek-Nesselquist |
| EPI_ISL_1227110, EPI_ISL_1227111, EPI_ISL_1227112 | TEMPUS LABS INC | Wadsworth Center, New York State Department of Health | Kirsten St. George, Daryl M. Lamson, Alexis Russel, Matthew Shudt, Melissa A Leisner, Jonathan Plitnick, Navjot Singh, John Kelly, Erasmus Schneider, Erica Lasek-Nesselquist |
| EPI_ISL_1227113, EPI_ISL_1227114, EPI_ISL_1227115, EPI_ISL_1227116, EPI_ISL_1227117, EPI_ISL_1227118, EPI_ISL_1227119, EPI_ISL_1227120, EPI_ISL_1227121, EPI_ISL_1227122, EPI_ISL_1227123, EPI_ISL_1227124, EPI_ISL_1227125, EPI_ISL_1227126, EPI_ISL_1227127, EPI_ISL_1227128 | | | |
| see above | NYC Pandemic Response Lab | Wadsworth Center, New York State Department of Health | Kirsten St. George, Daryl M. Lamson, Alexis Russel, Matthew Shudt, Melissa A Leisner, Jonathan Plitnick, Navjot Singh, John Kelly, Erasmus Schneider, Erica Lasek-Nesselquist |
| EPI_ISL_1227129, EPI_ISL_1227130 | ALBANY MEDICAL CENTER | Wadsworth Center, New York State Department of Health | Kirsten St. George, Daryl M. Lamson, Alexis Russel, Matthew Shudt, Melissa A Leisner, Jonathan Plitnick, Navjot Singh, John Kelly, Erasmus Schneider, Erica Lasek-Nesselquist |
| EPI_ISL_1227131, EPI_ISL_1227132, EPI_ISL_1227133 | MEMORIAL SLOAN KETTERING CANCER CENTER | Wadsworth Center, New York State Department of Health | Kirsten St. George, Daryl M. Lamson, Alexis Russel, Matthew Shudt, Melissa A Leisner, Jonathan Plitnick, Navjot Singh, John Kelly, Erasmus Schneider, Erica Lasek-Nesselquist |
| EPI_ISL_1227134, EPI_ISL_1227135, EPI_ISL_1227136 | WESTCHESTER MEDICAL CENTER | Wadsworth Center, New York State Department of Health | Kirsten St. George, Daryl M. Lamson, Alexis Russel, Matthew Shudt, Melissa A Leisner, Jonathan Plitnick, Navjot Singh, John Kelly, Erasmus Schneider, Erica Lasek-Nesselquist |
| EPI_ISL_1227137, EPI_ISL_1227138, EPI_ISL_1227139 | MONTEFIORE MEDICAL CENTER LABORATORIES | Wadsworth Center, New York State Department of Health | Kirsten St. George, Daryl M. Lamson, Alexis Russel, Matthew Shudt, Melissa A Leisner, Jonathan Plitnick, Navjot Singh, John Kelly, Erasmus Schneider, Erica Lasek-Nesselquist |
| EPI_ISL_1227140 | ALBANY MEDICAL CENTER | Wadsworth Center, New York State Department of Health | Kirsten St. George, Daryl M. Lamson, Alexis Russel, Matthew Shudt, Melissa A Leisner, Jonathan Plitnick, Navjot Singh, John Kelly, Erasmus Schneider, Erica Lasek-Nesselquist |
| EPI_ISL_1227142, EPI_ISL_1227143 | NYC Pandemic Response Lab | Wadsworth Center, New York State Department of Health | Kirsten St. George, Daryl M. Lamson, Alexis Russel, Matthew Shudt, Melissa A Leisner, Jonathan Plitnick, Navjot Singh, John Kelly, Erasmus Schneider, Erica Lasek-Nesselquist |
| EPI_ISL_1227145, EPI_ISL_1227146 | URMC LABS | Wadsworth Center, New York State Department of Health | Kirsten St. George, Daryl M. Lamson, Alexis Russel, Matthew Shudt, Melissa A Leisner, Jonathan Plitnick, Navjot Singh, John Kelly, Erasmus Schneider, Erica Lasek-Nesselquist |
| EPI_ISL_1227147, EPI_ISL_1227148 | NYC Pandemic Response Lab | Wadsworth Center, New York State Department of Health | Kirsten St. George, Daryl M. Lamson, Alexis Russel, Matthew Shudt, Melissa A Leisner, Jonathan Plitnick, Navjot Singh, John Kelly, Erasmus Schneider, Erica Lasek-Nesselquist |
| EPI_ISL_1227149 | WESTCHESTER MEDICAL CENTER | Wadsworth Center, New York State Department of Health | Kirsten St. George, Daryl M. Lamson, Alexis Russel, Matthew Shudt, Melissa A Leisner, Jonathan Plitnick, Navjot Singh, John Kelly, Erasmus Schneider, Erica Lasek-Nesselquist |
| EPI_ISL_1227150 | SUNY UPSTATE MEDICAL UNIVERSITY | Wadsworth Center, New York State Department of Health | Kirsten St. George, Daryl M. Lamson, Alexis Russel, Matthew Shudt, Melissa A Leisner, Jonathan Plitnick, Navjot Singh, John Kelly, Erasmus Schneider, Erica Lasek-Nesselquist |
| EPI_ISL_1227151 | MONTEFIORE MEDICAL CENTER LABORATORIES | Wadsworth Center, New York State Department of Health | Kirsten St. George, Daryl M. Lamson, Alexis Russel, Matthew Shudt, Melissa A Leisner, Jonathan Plitnick, Navjot Singh, John Kelly, Erasmus Schneider, Erica Lasek-Nesselquist |
| EPI_ISL_1227152 | WESTCHESTER MEDICAL CENTER | Wadsworth Center, New York State Department of Health | Kirsten St. George, Daryl M. Lamson, Alexis Russel, Matthew Shudt, Melissa A Leisner, Jonathan Plitnick, Navjot Singh, John Kelly, Erasmus Schneider, Erica Lasek-Nesselquist |
| EPI_ISL_1227153, EPI_ISL_1227154 | MEMORIAL SLOAN KETTERING CANCER CENTER | Wadsworth Center, New York State Department of Health | Kirsten St. George, Daryl M. Lamson, Alexis Russel, Matthew Shudt, Melissa A Leisner, Jonathan Plitnick, Navjot Singh, John Kelly, Erasmus Schneider, Erica Lasek-Nesselquist |
| EPI_ISL_1227155, EPI_ISL_1227156, EPI_ISL_1227157 | MONTEFIORE MEDICAL CENTER LABORATORIES | Wadsworth Center, New York State Department of Health | Kirsten St. George, Daryl M. Lamson, Alexis Russel, Matthew Shudt, Melissa A Leisner, Jonathan Plitnick, Navjot Singh, John Kelly, Erasmus Schneider, Erica Lasek-Nesselquist |
| EPI_ISL_1227158 | NORTH SHORE UNIVERSITY HOSPITAL | Wadsworth Center, New York State Department of Health | Kirsten St. George, Daryl M. Lamson, Alexis Russel, Matthew Shudt, Melissa A Leisner, Jonathan Plitnick, Navjot Singh, John Kelly, Erasmus Schneider, Erica Lasek-Nesselquist |
| EPI_ISL_1227159 | Columbia University Irving Medical Center | Wadsworth Center, New York State Department of Health | Kirsten St. George, Daryl M. Lamson, Alexis Russel, Matthew Shudt, Melissa A Leisner, Jonathan Plitnick, Navjot Singh, John Kelly, Erasmus Schneider, Erica Lasek-Nesselquist |
| EPI_ISL_1227160, EPI_ISL_1227161 | WESTCHESTER MEDICAL CENTER | Wadsworth Center, New York State Department of Health | Kirsten St. George, Daryl M. Lamson, Alexis Russel, Matthew Shudt, Melissa A Leisner, Jonathan Plitnick, Navjot Singh, John Kelly, Erasmus Schneider, Erica Lasek-Nesselquist |
| EPI_ISL_1227162 | SUNY UPSTATE MEDICAL UNIVERSITY | Wadsworth Center, New York State Department of Health | Kirsten St. George, Daryl M. Lamson, Alexis Russel, Matthew Shudt, Melissa A Leisner, Jonathan Plitnick, Navjot Singh, John Kelly, Erasmus Schneider, Erica Lasek-Nesselquist |
| EPI_ISL_1227163 | MEMORIAL SLOAN KETTERING CANCER CENTER | Wadsworth Center, New York State Department of Health | Kirsten St. George, Daryl M. Lamson, Alexis Russel, Matthew Shudt, Melissa A Leisner, Jonathan Plitnick, Navjot Singh, John Kelly, Erasmus Schneider, Erica Lasek-Nesselquist |
| EPI_ISL_1227164 | WESTCHESTER MEDICAL CENTER | Wadsworth Center, New York State Department of Health | Kirsten St. George, Daryl M. Lamson, Alexis Russel, Matthew Shudt, Melissa A Leisner, Jonathan Plitnick, Navjot Singh, John Kelly, Erasmus Schneider, Erica Lasek-Nesselquist |
| EPI_ISL_1227165 | URMC LABS | Wadsworth Center, New York State Department of Health | Kirsten St. George, Daryl M. Lamson, Alexis Russel, Matthew Shudt, Melissa A Leisner, Jonathan Plitnick, Navjot Singh, John Kelly, Erasmus Schneider, Erica Lasek-Nesselquist |
| EPI_ISL_1227166 | NYC Pandemic Response Lab | Wadsworth Center, New York State Department of Health | Kirsten St. George, Daryl M. Lamson, Alexis Russel, Matthew Shudt, Melissa A Leisner, Jonathan Plitnick, Navjot Singh, John Kelly, Erasmus Schneider, Erica Lasek-Nesselquist |
| EPI_ISL_1227167 | MEMORIAL SLOAN KETTERING CANCER CENTER | Wadsworth Center, New York State Department of Health | Kirsten St. George, Daryl M. Lamson, Alexis Russel, Matthew Shudt, Melissa A Leisner, Jonathan Plitnick, Navjot Singh, John Kelly, Erasmus Schneider, Erica Lasek-Nesselquist |
| EPI_ISL_1227168 | MONTEFIORE MEDICAL CENTER LABORATORIES | Wadsworth Center, New York State Department of Health | Kirsten St. George, Daryl M. Lamson, Alexis Russel, Matthew Shudt, Melissa A Leisner, Jonathan Plitnick, Navjot Singh, John Kelly, Erasmus Schneider, Erica Lasek-Nesselquist |

| | | | |
|---|---|---|---|
| EPI_ISL_1227169 | TEMPUS LABS INC | Wadsworth Center, New York State Department of Health | Kirsten St. George, Daryl M. Lamson, Alexis Russel, Matthew Shudt, Melissa A Leisner, Jonathan Plitnick, Navjot Singh, John Kelly, Erasmus Schneider, Erica Lasek-Nesselquist |
| EPI_ISL_1227170, EPI_ISL_1227171 | NYC Pandemic Response Lab | Wadsworth Center, New York State Department of Health | Kirsten St. George, Daryl M. Lamson, Alexis Russel, Matthew Shudt, Melissa A Leisner, Jonathan Plitnick, Navjot Singh, John Kelly, Erasmus Schneider, Erica Lasek-Nesselquist |
| EPI_ISL_1227172 | MONTEFIORE MEDICAL CENTER LABORATORIES | Wadsworth Center, New York State Department of Health | Kirsten St. George, Daryl M. Lamson, Alexis Russel, Matthew Shudt, Melissa A Leisner, Jonathan Plitnick, Navjot Singh, John Kelly, Erasmus Schneider, Erica Lasek-Nesselquist |
| EPI_ISL_1227173 | NYC Pandemic Response Lab | Wadsworth Center, New York State Department of Health | Kirsten St. George, Daryl M. Lamson, Alexis Russel, Matthew Shudt, Melissa A Leisner, Jonathan Plitnick, Navjot Singh, John Kelly, Erasmus Schneider, Erica Lasek-Nesselquist |
| EPI_ISL_1227174, EPI_ISL_1227175 | MEMORIAL SLOAN KETTERING CANCER CENTER | Wadsworth Center, New York State Department of Health | Kirsten St. George, Daryl M. Lamson, Alexis Russel, Matthew Shudt, Melissa A Leisner, Jonathan Plitnick, Navjot Singh, John Kelly, Erasmus Schneider, Erica Lasek-Nesselquist |
| EPI_ISL_1227176 | URMC LABS | Wadsworth Center, New York State Department of Health | Kirsten St. George, Daryl M. Lamson, Alexis Russel, Matthew Shudt, Melissa A Leisner, Jonathan Plitnick, Navjot Singh, John Kelly, Erasmus Schneider, Erica Lasek-Nesselquist |
| EPI_ISL_1227177 | NYC Pandemic Response Lab | Wadsworth Center, New York State Department of Health | Kirsten St. George, Daryl M. Lamson, Alexis Russel, Matthew Shudt, Melissa A Leisner, Jonathan Plitnick, Navjot Singh, John Kelly, Erasmus Schneider, Erica Lasek-Nesselquist |
| EPI_ISL_1227179, EPI_ISL_1227180 | WESTCHESTER MEDICAL CENTER | Wadsworth Center, New York State Department of Health | Kirsten St. George, Daryl M. Lamson, Alexis Russel, Matthew Shudt, Melissa A Leisner, Jonathan Plitnick, Navjot Singh, John Kelly, Erasmus Schneider, Erica Lasek-Nesselquist |
| EPI_ISL_1227181 | MONTEFIORE MEDICAL CENTER LABORATORIES | Wadsworth Center, New York State Department of Health | Kirsten St. George, Daryl M. Lamson, Alexis Russel, Matthew Shudt, Melissa A Leisner, Jonathan Plitnick, Navjot Singh, John Kelly, Erasmus Schneider, Erica Lasek-Nesselquist |
| EPI_ISL_1227183 | WESTCHESTER MEDICAL CENTER | Wadsworth Center, New York State Department of Health | Kirsten St. George, Daryl M. Lamson, Alexis Russel, Matthew Shudt, Melissa A Leisner, Jonathan Plitnick, Navjot Singh, John Kelly, Erasmus Schneider, Erica Lasek-Nesselquist |
| EPI_ISL_1227184 | MONTEFIORE MEDICAL CENTER LABORATORIES | Wadsworth Center, New York State Department of Health | Kirsten St. George, Daryl M. Lamson, Alexis Russel, Matthew Shudt, Melissa A Leisner, Jonathan Plitnick, Navjot Singh, John Kelly, Erasmus Schneider, Erica Lasek-Nesselquist |
| EPI_ISL_1227185 | URMC LABS | Wadsworth Center, New York State Department of Health | Kirsten St. George, Daryl M. Lamson, Alexis Russel, Matthew Shudt, Melissa A Leisner, Jonathan Plitnick, Navjot Singh, John Kelly, Erasmus Schneider, Erica Lasek-Nesselquist |
| EPI_ISL_1227186 | NYC Pandemic Response Lab | Wadsworth Center, New York State Department of Health | Kirsten St. George, Daryl M. Lamson, Alexis Russel, Matthew Shudt, Melissa A Leisner, Jonathan Plitnick, Navjot Singh, John Kelly, Erasmus Schneider, Erica Lasek-Nesselquist |
| EPI_ISL_1227187 | ALBANY MEDICAL CENTER | Wadsworth Center, New York State Department of Health | Kirsten St. George, Daryl M. Lamson, Alexis Russel, Matthew Shudt, Melissa A Leisner, Jonathan Plitnick, Navjot Singh, John Kelly, Erasmus Schneider, Erica Lasek-Nesselquist |
| EPI_ISL_1227188, EPI_ISL_1227189 | Columbia University Irving Medical Center | Wadsworth Center, New York State Department of Health | Kirsten St. George, Daryl M. Lamson, Alexis Russel, Matthew Shudt, Melissa A Leisner, Jonathan Plitnick, Navjot Singh, John Kelly, Erasmus Schneider, Erica Lasek-Nesselquist |
| EPI_ISL_1227190, EPI_ISL_1227191 | MONTEFIORE MEDICAL CENTER LABORATORIES | Wadsworth Center, New York State Department of Health | Kirsten St. George, Daryl M. Lamson, Alexis Russel, Matthew Shudt, Melissa A Leisner, Jonathan Plitnick, Navjot Singh, John Kelly, Erasmus Schneider, Erica Lasek-Nesselquist |
| EPI_ISL_1227192, EPI_ISL_1227193, EPI_ISL_1227194, EPI_ISL_1227195, EPI_ISL_1227196, EPI_ISL_1227197, EPI_ISL_1227198, EPI_ISL_1227199 | NYC Pandemic Response Lab | Wadsworth Center, New York State Department of Health | Kirsten St. George, Daryl M. Lamson, Alexis Russel, Matthew Shudt, Melissa A Leisner, Jonathan Plitnick, Navjot Singh, John Kelly, Erasmus Schneider, Erica Lasek-Nesselquist |
| EPI_ISL_1227200, EPI_ISL_1227201, EPI_ISL_1227202 | WESTCHESTER MEDICAL CENTER | Wadsworth Center, New York State Department of Health | Kirsten St. George, Daryl M. Lamson, Alexis Russel, Matthew Shudt, Melissa A Leisner, Jonathan Plitnick, Navjot Singh, John Kelly, Erasmus Schneider, Erica Lasek-Nesselquist |
| EPI_ISL_1227203 | MONTEFIORE MEDICAL CENTER LABORATORIES | Wadsworth Center, New York State Department of Health | Kirsten St. George, Daryl M. Lamson, Alexis Russel, Matthew Shudt, Melissa A Leisner, Jonathan Plitnick, Navjot Singh, John Kelly, Erasmus Schneider, Erica Lasek-Nesselquist |
| EPI_ISL_1227204, EPI_ISL_1227205 | WESTCHESTER MEDICAL CENTER | Wadsworth Center, New York State Department of Health | Kirsten St. George, Daryl M. Lamson, Alexis Russel, Matthew Shudt, Melissa A Leisner, Jonathan Plitnick, Navjot Singh, John Kelly, Erasmus Schneider, Erica Lasek-Nesselquist |
| EPI_ISL_1227206 | MEMORIAL SLOAN KETTERING CANCER CENTER | Wadsworth Center, New York State Department of Health | Kirsten St. George, Daryl M. Lamson, Alexis Russel, Matthew Shudt, Melissa A Leisner, Jonathan Plitnick, Navjot Singh, John Kelly, Erasmus Schneider, Erica Lasek-Nesselquist |
| EPI_ISL_1227207 | TEMPUS LABS INC | Wadsworth Center, New York State Department of Health | Kirsten St. George, Daryl M. Lamson, Alexis Russel, Matthew Shudt, Melissa A Leisner, Jonathan Plitnick, Navjot Singh, John Kelly, Erasmus Schneider, Erica Lasek-Nesselquist |
| EPI_ISL_1227208 | MONTEFIORE MEDICAL CENTER LABORATORIES | Wadsworth Center, New York State Department of Health | Kirsten St. George, Daryl M. Lamson, Alexis Russel, Matthew Shudt, Melissa A Leisner, Jonathan Plitnick, Navjot Singh, John Kelly, Erasmus Schneider, Erica Lasek-Nesselquist |
| EPI_ISL_1227209 | BIO-REFERENCE LABORATORIES | Wadsworth Center, New York State Department of Health | Kirsten St. George, Daryl M. Lamson, Alexis Russel, Matthew Shudt, Melissa A Leisner, Jonathan Plitnick, Navjot Singh, John Kelly, Erasmus Schneider, Erica Lasek-Nesselquist |
| EPI_ISL_1227210, EPI_ISL_1227211 | MONTEFIORE MEDICAL CENTER LABORATORIES | Wadsworth Center, New York State Department of Health | Kirsten St. George, Daryl M. Lamson, Alexis Russel, Matthew Shudt, Melissa A Leisner, Jonathan Plitnick, Navjot Singh, John Kelly, Erasmus Schneider, Erica Lasek-Nesselquist |
| EPI_ISL_1227212 | NORTH SHORE UNIVERSITY HOSPITAL | Wadsworth Center, New York State Department of Health | Kirsten St. George, Daryl M. Lamson, Alexis Russel, Matthew Shudt, Melissa A Leisner, Jonathan Plitnick, Navjot Singh, John Kelly, Erasmus Schneider, Erica Lasek-Nesselquist |
| EPI_ISL_1227213, EPI_ISL_1227214 | MONTEFIORE MEDICAL CENTER LABORATORIES | Wadsworth Center, New York State Department of Health | Kirsten St. George, Daryl M. Lamson, Alexis Russel, Matthew Shudt, Melissa A Leisner, Jonathan Plitnick, Navjot Singh, John Kelly, Erasmus Schneider, Erica Lasek-Nesselquist |
| EPI_ISL_1227215 | MEMORIAL SLOAN KETTERING CANCER CENTER | Wadsworth Center, New York State Department of Health | Kirsten St. George, Daryl M. Lamson, Alexis Russel, Matthew Shudt, Melissa A Leisner, Jonathan Plitnick, Navjot Singh, John Kelly, Erasmus Schneider, Erica Lasek-Nesselquist |
| EPI_ISL_1227216 | GLENS FALLS HOSPITAL LABORATORY | Wadsworth Center, New York State Department of Health | Kirsten St. George, Daryl M. Lamson, Alexis Russel, Matthew Shudt, Melissa A Leisner, Jonathan Plitnick, Navjot Singh, John Kelly, Erasmus Schneider, Erica Lasek-Nesselquist |
| EPI_ISL_1227217 | MONTEFIORE MEDICAL CENTER LABORATORIES | Wadsworth Center, New York State Department of Health | Kirsten St. George, Daryl M. Lamson, Alexis Russel, Matthew Shudt, Melissa A Leisner, Jonathan Plitnick, Navjot Singh, John Kelly, Erasmus Schneider, Erica Lasek-Nesselquist |
| EPI_ISL_1227218, EPI_ISL_1227219, EPI_ISL_1227220 | WESTCHESTER MEDICAL CENTER | Wadsworth Center, New York State Department of Health | Kirsten St. George, Daryl M. Lamson, Alexis Russel, Matthew Shudt, Melissa A Leisner, Jonathan Plitnick, Navjot Singh, John Kelly, Erasmus Schneider, Erica Lasek-Nesselquist |
| EPI_ISL_1227221 | ALBANY MEDICAL CENTER | Wadsworth Center, New York State Department of Health | Kirsten St. George, Daryl M. Lamson, Alexis Russel, Matthew Shudt, Melissa A Leisner, Jonathan Plitnick, Navjot Singh, John Kelly, Erasmus Schneider, Erica Lasek-Nesselquist |
| EPI_ISL_1227222 | NYC Pandemic Response Lab | Wadsworth Center, New York State Department of Health | Kirsten St. George, Daryl M. Lamson, Alexis Russel, Matthew Shudt, Melissa A Leisner, Jonathan Plitnick, Navjot Singh, John Kelly, Erasmus Schneider, Erica Lasek-Nesselquist |
| EPI_ISL_1227223 | Columbia University Irving Medical Center | Wadsworth Center, New York State Department of Health | Kirsten St. George, Daryl M. Lamson, Alexis Russel, Matthew Shudt, Melissa A Leisner, Jonathan Plitnick, Navjot Singh, John Kelly, Erasmus Schneider, Erica Lasek-Nesselquist |
| EPI_ISL_1227224 | NYC Pandemic Response Lab | Wadsworth Center, New York State Department of Health | Kirsten St. George, Daryl M. Lamson, Alexis Russel, Matthew Shudt, Melissa A Leisner, Jonathan Plitnick, Navjot Singh, John Kelly, Erasmus Schneider, |

| | | | |
|---|---|---|---|
| | | | Erica Lasek-Nesselquist |
| EPI_ISL_1227225 | URMC LABS | Wadsworth Center, New York State Department of Health | Kirsten St. George, Daryl M. Lamson, Alexis Russel, Matthew Shudt, Melissa A Leisner, Jonathan Plitnick, Navjot Singh, John Kelly, Erasmus Schneider, Erica Lasek-Nesselquist |
| EPI_ISL_1227227, EPI_ISL_1227228 | MONTEFIORE MEDICAL CENTER LABORATORIES | Wadsworth Center, New York State Department of Health | Kirsten St. George, Daryl M. Lamson, Alexis Russel, Matthew Shudt, Melissa A Leisner, Jonathan Plitnick, Navjot Singh, John Kelly, Erasmus Schneider, Erica Lasek-Nesselquist |
| EPI_ISL_1227229 | NYC Pandemic Response Lab | Wadsworth Center, New York State Department of Health | Kirsten St. George, Daryl M. Lamson, Alexis Russel, Matthew Shudt, Melissa A Leisner, Jonathan Plitnick, Navjot Singh, John Kelly, Erasmus Schneider, Erica Lasek-Nesselquist |
| EPI_ISL_1230479 | Public Health Ontario Laboratory | Public Health Ontario Laboratory | Vanessa G Allen, Philip Banh, Yao Chen, Richard de Borja, Alireza Eshaghi, Nahuel Fittipaldi, Christine Frantz, Jonathan B Gubbay, Jennifer L Guthrie, Lawrence Heisler, Esha Joshi, Michael Laszloffy, Aimin Li, Michael CY Li, Dean Maxwell, Sandeep Nagra, Samir N Patel, Jared Simpson, Karthikeyan Sivaraman, Ashleigh Sullivan, Yogi Sundaravadanam, Sarah Teatero, Andre Villegas, Matthew Watson, Sandra Zittermann |
| EPI_ISL_1231730 | Fulgent Genetics | Fulgent Genetics | Harry Gao, Mickey Li, John Gao, Joseph Fierro, Benafsh Sapra, Becky Tsai, Yan Meng, Doreen Ng, James Xie |
| EPI_ISL_1233804 | Michigan Department of Health and Human Services, Bureau of Laboratories | Michigan Department of Health and Human Services, Bureau of Laboratories | Blankenship HM, Riner D, Soehnlen MK |
| EPI_ISL_1234121, EPI_ISL_1234210, EPI_ISL_1234249, EPI_ISL_1234310, EPI_ISL_1234313, EPI_ISL_1234364 | Colorado Department of Public Health and Environment | Colorado Department of Public Health and Environment | Laura Bankers, Molly C. Hetherington-Rauth, Diana Ir, Shannon Ely, Shannon R. Matzinger, Sarah Elizabeth Totten, Emily A. Travanty |
| EPI_ISL_1235195 | CA DPH Viral and Rickettsial Disease Laboratory | Chan-Zuckerberg Biohub | CZB Cliahub Consortium |
| EPI_ISL_1236537, EPI_ISL_1236576 | Houston Methodist Hospital | Houston Methodist Hospital | S. Wesley Long, Randall J. Olsen, Paul A. Christensen, Sishir Subedi, Robert Olson, James J. Davis, Matthew Ojeda Saavedra, Prasanti Yerramilli, Layne Pruitt, Kristina Reppond, Madison N. Shyer, Jessica Cambric, Ilya J. Finkelstein, Jimmy Gollihar, and James M. Musser |
| EPI_ISL_1238836 | MD PHL | MD PHL | Maryland Department of Health Laboratories Administration |
| EPI_ISL_1238905, EPI_ISL_1238944, EPI_ISL_1238957 | Johns Hopkins Hospital Department of Pathology | Johns Hopkins Hospital Department of Pathology | C. Paul Morris, Chun Huai Luo, Adannaya Amadi, Matthew Schwartz, Nicholas Gallagher, Heba H. Mostafa |
| EPI_ISL_683762 | DOHMH Morrisania | New York City Public Health Laboratory | Jade Wang, et al. |
| EPI_ISL_765493, EPI_ISL_765494, EPI_ISL_765495 | MONTEFIORE MEDICAL CENTER LABORATORIES | Wadsworth Center, New York State Department.of Health | Kirsten St. George, Daryl M. Lamson, Alexis Russel, Matthew Shudt, Melissa A Leisner, Jonathan Plitnick, Navjot Singh, John Kelly, Sara Griesemer, Erasmus Schneider, Erica Lasek-Nesselquist |
| EPI_ISL_767581, EPI_ISL_767595, EPI_ISL_767598 | WHITE PLAINS HOSPITAL CENTER LABORATORY | Wadsworth Center, New York State Department.of Health | Kirsten St. George, Daryl M. Lamson, Alexis Russel, Matthew Shudt, Melissa A Leisner, Jonathan Plitnick, Navjot Singh, John Kelly, Sara Griesemer, Erasmus Schneider, Erica Lasek-Nesselquist |
| EPI_ISL_794226, EPI_ISL_802448 | WESTCHESTER MEDICAL CENTER | Wadsworth Center, New York State Department.of Health | Kirsten St. George, Daryl M. Lamson, Alexis Russel, Matthew Shudt, Melissa A Leisner, Jonathan Plitnick, Navjot Singh, John Kelly, Sara Griesemer, Erasmus Schneider, Erica Lasek-Nesselquist |
| EPI_ISL_802788 | Columbia University Irving Medical Center | Wadsworth Center, New York State Department.of Health | Kirsten St. George, Daryl M. Lamson, Alexis Russel, Matthew Shudt, Melissa A Leisner, Jonathan Plitnick, Navjot Singh, John Kelly, Sara Griesemer, Erasmus Schneider, Erica Lasek-Nesselquist |
| EPI_ISL_812733 | DOHMH Corona | New York City Public Health Laboratory | Jade Wang, et al. |
| EPI_ISL_823886 | DOHMH Central Harlem | New York City Public Health Laboratory | Jade Wang, et al. |
| EPI_ISL_830577, EPI_ISL_830578 | NORTHWELL HEALTH LABORATORIES | Wadsworth Center, New York State Department of Health | Kirsten St. George, Daryl M. Lamson, Alexis Russel, Matthew Shudt, Melissa A Leisner, Jonathan Plitnick, Navjot Singh, John Kelly, Erasmus Schneider, Erica Lasek-Nesselquist |
| EPI_ISL_830720 | BIO-REFERENCE LABORATORIES | Wadsworth Center, New York State Department of Health | Kirsten St. George, Daryl M. Lamson, Alexis Russel, Matthew Shudt, Melissa A Leisner, Jonathan Plitnick, Navjot Singh, John Kelly, Erasmus Schneider, Erica Lasek-Nesselquist |
| EPI_ISL_832270, EPI_ISL_832271 | DOHMH Riverside | New York City Public Health Laboratory | Jade Wang, et al. |
| EPI_ISL_849348 | Delaware Public Health Lab | Delaware Public Health Lab | Gregory Hovan |
| EPI_ISL_850743 | Helix / Illumina | Genomics and Discovery, Respiratory Viruses Branch, Division of Viral Diseases, Centers for Disease Control and Prevention | Peter W. Cook, Dhwani Batra, Ben L. Rambo-Martin Eileen de Feo, Jan Antico, Christine Tran, Matthew Tolentino, Shannon Wickline, Kim Gietzen, Brad Sickler, Jingtao Liu, Eric Allen, Phil Febbo, Summer Galloway, Nicole L. Washington, Simon White, Geraint Levan, Kelly Schiabor Barrett, Elizabeth Cirulli, Alexandre Bolze, Ary Ascencio, Charlotte Rivera-Garcia, Ryan Cho, Jason Nguyen, Sherry Wang, Jimmy Ramirez, Tyler Cassens, Efren Sandoval, Magnus Isaksson, William Lee, David Becker, Marc Laurent, James Lu, Clinton R. Paden, Suxiang Tong, Duncan MacCannell |
| EPI_ISL_854449 | WESTCHESTER MEDICAL CENTER | Wadsworth Center, New York State Department of Health | Kirsten St. George, Daryl M. Lamson, Alexis Russel, Matthew Shudt, Melissa A Leisner, Jonathan Plitnick, Navjot Singh, John Kelly, Erasmus Schneider, Erica Lasek-Nesselquist |
| EPI_ISL_854450, EPI_ISL_854457, EPI_ISL_854458, EPI_ISL_854459 | MONTEFIORE MEDICAL CENTER LABORATORIES | Wadsworth Center, New York State Department of Health | Kirsten St. George, Daryl M. Lamson, Alexis Russel, Matthew Shudt, Melissa A Leisner, Jonathan Plitnick, Navjot Singh, John Kelly, Erasmus Schneider, Erica Lasek-Nesselquist |
| EPI_ISL_855171 | Quest Diagnostics | Quest Diagnostics | Rosenthal,S.H., Gerasimova,A., Kagan,R.M., Anderson, B., Hua, M., Liu Y., Bernstein, L.E., Livingston, K.E., Perez, A., Shalhout, D.F., Shlyakhter, I.A., Owen, R., Tanpaiboon, P., Lacbawan, F. |
| EPI_ISL_857056, EPI_ISL_857060 | OCME Office Of Chief Medical Examiner | New York City Public Health Laboratory | Jade Wang, et al. |
| EPI_ISL_857120, EPI_ISL_857163 | DOHMH Central Harlem | New York City Public Health Laboratory | Jade Wang, et al. |
| EPI_ISL_857164 | DOHMH Morrisania | New York City Public Health Laboratory | Jade Wang, et al. |
| EPI_ISL_857204 | DOHMH Central Harlem | New York City Public Health Laboratory | Jade Wang, et al. |
| EPI_ISL_861111, EPI_ISL_861112, EPI_ISL_861113 | New York Presbyterian Hospital | Wadsworth Center, New York State Department of Health | Kirsten St. George, Daryl M. Lamson, Alexis Russel, Matthew Shudt, Melissa A Leisner, Jonathan Plitnick, Navjot Singh, John Kelly, Erasmus Schneider, Erica Lasek-Nesselquist |
| EPI_ISL_861175 | BIO-REFERENCE LABORATORIES | Wadsworth Center, New York State Department of Health | Kirsten St. George, Daryl M. Lamson, Alexis Russel, Matthew Shudt, Melissa A Leisner, Jonathan Plitnick, Navjot Singh, John Kelly, Erasmus Schneider, Erica Lasek-Nesselquist |
| EPI_ISL_861188, EPI_ISL_861189 | NORTH SHORE UNIVERSITY HOSPITAL | Wadsworth Center, New York State Department of Health | Kirsten St. George, Daryl M. Lamson, Alexis Russel, Matthew Shudt, Melissa A Leisner, Jonathan Plitnick, Navjot Singh, John Kelly, Erasmus Schneider, Erica Lasek-Nesselquist |
| EPI_ISL_861244, EPI_ISL_861258, EPI_ISL_861278, EPI_ISL_861280, EPI_ISL_861283, EPI_ISL_861285, EPI_ISL_861300, EPI_ISL_861306, EPI_ISL_861308, EPI_ISL_861315 | MONTEFIORE MEDICAL CENTER LABORATORIES | Wadsworth Center, New York State Department of Health | Kirsten St. George, Daryl M. Lamson, Alexis Russel, Matthew Shudt, Melissa A Leisner, Jonathan Plitnick, Navjot Singh, John Kelly, Erasmus Schneider, Erica Lasek-Nesselquist |
| EPI_ISL_861361, EPI_ISL_861411 | WESTCHESTER MEDICAL CENTER | Wadsworth Center, New York State Department of Health | Kirsten St. George, Daryl M. Lamson, Alexis Russel, Matthew Shudt, Melissa A Leisner, Jonathan Plitnick, Navjot Singh, John Kelly, Erasmus Schneider, Erica Lasek-Nesselquist |
| EPI_ISL_883330 | OCME Office Of Chief Medical Examiner | New York City Public Health Laboratory | Jade Wang, et al. |
| EPI_ISL_883331, EPI_ISL_883338 | DOHMH Morrisania | New York City Public Health Laboratory | Jade Wang, et al. |
| EPI_ISL_883339 | DOHMH Chelsea | New York City Public Health Laboratory | Jade Wang, et al. |

| | | | |
|---|---|---|---|
| EPI_ISL_883340, EPI_ISL_883341, EPI_ISL_883342, EPI_ISL_883343 | DOHMH PHL | New York City Public Health Laboratory | Jade Wang, et al. |
| EPI_ISL_883344, EPI_ISL_883345 | OCME Office Of Chief Medical Examiner | New York City Public Health Laboratory | Jade Wang, et al. |
| EPI_ISL_883346 | DOHMH PHL | New York City Public Health Laboratory | Jade Wang, et al. |
| EPI_ISL_883347 | DOHMH Morrisania | New York City Public Health Laboratory | Jade Wang, et al. |
| EPI_ISL_883447 | NORTHWELL HEALTH LABORATORIES | Wadsworth Center, New York State Department of Health | Kirsten St. George, Daryl M. Lamson, Alexis Russel, Matthew Shudt, Melissa A Leisner, Jonathan Plitnick, Navjot Singh, John Kelly, Erasmus Schneider, Erica Lasek-Nesselquist |
| EPI_ISL_884055, EPI_ISL_884080 | Wadsworth Center, New York State Department of Health | Wadsworth Center, New York State Department of Health | Kirsten St. George, Daryl M. Lamson, Alexis Russel, Matthew Shudt, Melissa A Leisner, Jonathan Plitnick, Navjot Singh, John Kelly, Erasmus Schneider, Erica Lasek-Nesselquist |
| EPI_ISL_886240, EPI_ISL_886270, EPI_ISL_886271, EPI_ISL_886384, EPI_ISL_886531, EPI_ISL_886626, EPI_ISL_887843, EPI_ISL_888376 | Labcorp | Genomics and Discovery, Respiratory Viruses Branch, Division of Viral Diseases, Centers for Disease Control and Prevention | Peter W. Cook,Dhwani Batra,Ben L. Rambo-Martin,Summer Galloway,Brian Krueger,Minoo Agarwal,Eyad Almasri,Debbie Boles,Ayla Burns,Nuthawin Charoensri,Oren Cohen,Susan Countryman,Mary Ann Cristobal,Bobbi Croy,Suzanne Dale,Hrushikesh Deshmukh,Amanda Douglas,Vincent Drouillon,Marcia Eisenberg,Howard Engler,Rama Ghatti,Prashant Gupta,Susan Hicks,Jake Humphrey,Lax Iyer,Manoj Jain,Mohan Kolli,Tim Kuphal,Stanley Letovsky,Michael Levandoski,Craig Lukasik,Jonathan Meltzer,Brian Norvell,Mindy Nye,Scott Parker,Christos Petropoulos,John Pruitt,Steven Ragan,Scott Ryan,Mike Sapeta,Jana Schroth,Suresh Babu Selvaraju,Goran Stevovic,Amanda Suchanek,Andrea Throop,Lyndon Tilson,Thomas Urban,Joe Voshell,Kimberly Wagner,Jonathan Williams,Mary Williamson,Qian Zeng,Tricia Zwiefelhofer,Clinton R. Paden,Suxiang Tong,Duncan MacCannell, |
| EPI_ISL_896227, EPI_ISL_896233, EPI_ISL_896234, EPI_ISL_896391, EPI_ISL_896392, EPI_ISL_896394, EPI_ISL_896395 | Columbia University Irving Medical Center | Wadsworth Center, New York State Department of Health | Kirsten St. George, Daryl M. Lamson, Alexis Russel, Matthew Shudt, Melissa A Leisner, Jonathan Plitnick, Navjot Singh, John Kelly, Erasmus Schneider, Erica Lasek-Nesselquist |
| EPI_ISL_906837, EPI_ISL_906838, EPI_ISL_906839 | National Public Health Laboratory, National Centre for Infectious Diseases | National Public Health Laboratory, National Centre for Infectious Diseases | Tze Minn Mak, Zhenyang Zhou, Lin Cui, Raymond Tzer Pin Lin |
| EPI_ISL_911789 | Johns Hopkins Hospital Department of Pathology | Johns Hopkins Hospital Department of Pathology | C. Paul Morris, Chun Huai Luo, Adannaya Amadi, Matthew Schwartz, Nicholas Gallagher, Heba H. Mostafa |
| EPI_ISL_920158 | University College London, Great Ormond Street Hospital for Children NHS Foundation Trust, Imperial College Healthcare NHS Trust | COVID-19 Genomics UK (COG-UK) Consortium | Sergi Castellano, Rachel Williams, Mark Kristiansen, Paola Resende Silva, Sunando Roy, Tony Brooks, Helena Tutill, Paola Niola, Patricia Dyal, Charlotte Williams, Leysa Forrest, Yasmin Panchbhaya, Jacqueline Findlay, Samuel Weeks, Julianne Brown, Kathryn Harris, Paul Randell, James Price, Alison Holmes, Judith Breuer |
| EPI_ISL_936036, EPI_ISL_936038 | Columbia University Irving Medical Center | Wadsworth Center, New York State Department of Health | Kirsten St. George, Daryl M. Lamson, Alexis Russel, Matthew Shudt, Melissa A Leisner, Jonathan Plitnick, Navjot Singh, John Kelly, Erasmus Schneider, Erica Lasek-Nesselquist |
| EPI_ISL_936248, EPI_ISL_936254, EPI_ISL_936256, EPI_ISL_936261, EPI_ISL_936267, EPI_ISL_936271, EPI_ISL_936276, EPI_ISL_936281, EPI_ISL_936288, EPI_ISL_936291, EPI_ISL_936292, EPI_ISL_936299 | | | |
| see above | MONTEFIORE MEDICAL CENTER LABORATORIES | Wadsworth Center, New York State Department of Health | Kirsten St. George, Daryl M. Lamson, Alexis Russel, Matthew Shudt, Melissa A Leisner, Jonathan Plitnick, Navjot Singh, John Kelly, Erasmus Schneider, Erica Lasek-Nesselquist |
| EPI_ISL_937126 | DOHMH Riverside | New York City Public Health Laboratory | Jade Wang, et al. |
| EPI_ISL_937176 | DOHMH Jamaica | New York City Public Health Laboratory | Jade Wang, et al. |
| EPI_ISL_937190 | OCME Office Of Chief Medical Examiner | New York City Public Health Laboratory | Jade Wang, et al. |
| EPI_ISL_937191, EPI_ISL_937192 | Department of Homeless Services | New York City Public Health Laboratory | Jade Wang, et al. |
| EPI_ISL_937201 | DOHMH Fort Greene | New York City Public Health Laboratory | Jade Wang, et al. |
| EPI_ISL_937205 | DOHMH Central Harlem | New York City Public Health Laboratory | Jade Wang, et al. |
| EPI_ISL_937211 | DOHMH PHL | New York City Public Health Laboratory | Jade Wang, et al. |
| EPI_ISL_937213 | DOHMH Jamaica | New York City Public Health Laboratory | Jade Wang, et al. |
| EPI_ISL_937215 | DOHMH PHL | New York City Public Health Laboratory | Jade Wang, et al. |
| EPI_ISL_937216 | DOHMH Central Harlem | New York City Public Health Laboratory | Jade Wang, et al. |
| EPI_ISL_937218 | DOHMH Chelsea | New York City Public Health Laboratory | Jade Wang, et al. |
| EPI_ISL_937236 | DOHMH Morrisania | New York City Public Health Laboratory | Jade Wang, et al. |
| EPI_ISL_937246, EPI_ISL_937247, EPI_ISL_937255 | OCME Office Of Chief Medical Examiner | New York City Public Health Laboratory | Jade Wang, et al. |
| EPI_ISL_937258, EPI_ISL_937265, EPI_ISL_937272 | NYC HH Elmhurst Hospital Medical Center | New York City Public Health Laboratory | Jade Wang, et al. |
| EPI_ISL_937477 | Maine Health and Environmental Testing Laboratory (Maine HETL) | Tewhey Lab, The Jackson Laboratory | Matluk,N., Dewey,H., Iosue,F., Barter,M., Lynch,R., Munger,H. and Tewhey,R. |
| EPI_ISL_944592, EPI_ISL_944594 | Yale Clinical Virology Lab | Grubaugh Lab - Yale School of Public Health | Joseph Fauver, Tara Alpert, Anderson Brito, Mallery Breban, Anne Wyllie, Chantal Vogels, Mary Petrone, Annie Watkins, Chaney Kalinich, Isabel Ott, Nathan Grubaugh |
| EPI_ISL_962431 | Washington State Department of Health | Seattle Flu Study | Deborah A. Nickerson, Chris D. Frazar, Jover Lee, Benjamin Pelle, Erica Ryke, Matthew Richardson, Amanda Adler, Elisabeth Brandstetter, Peter D. Han, Kairsten Fay, Misja Ilcisin, Kirsten Lacombe, Thomas R. Sibley, Melissa Truong, Caitlin R. Wolf, Romesh Gautom, Geoff Melly, Brian Hiatt, Philip Dykema, Scott Lindquist, Michael Boeckh, Janet A. Englund, Michael Famulare, Barry R. Lutz, Mark J. Rieder, Lea M. Starita, Matthew Thompson, Helen Y. Chu, Jay Shendure, Trevor Bedford |
| EPI_ISL_962492, EPI_ISL_962493 | Omics Sciences Laboratory | Omics Sciences Laboratory | Derly Andrade Molina, Rubén Armas González, Gabriel Morey León, Darlyn Amaya, Katheryn Sacheri Viteri, Emily Sulay Saltos Montalvo, Paula Juliana Gavilanes Jarrín, Juan Carlos Fernández Cadena |
| EPI_ISL_966264 | NYU Langone Health | Departments of Pathology and Medicine, New York University School of Medicine | Adriana Heguy, Dacia Dimartino, Emily Guzman, Christian Marier, Peter Meyn, Sitharam Ramaswami, Gael Westby, Paul Zappile, Yutong Zhang, Paolo Cotzia, Guiqing Wang |
| EPI_ISL_966385 | Department of Homeless Services | New York City Public Health Laboratory | Jade Wang, et al. |
| EPI_ISL_966387 | DOHMH Jamaica | New York City Public Health Laboratory | Jade Wang, et al. |
| EPI_ISL_966400 | OCME Office Of Chief Medical Examiner | New York City Public Health Laboratory | Jade Wang, et al. |
| EPI_ISL_966401 | Department of Homeless Services | New York City Public Health Laboratory | Jade Wang, et al. |
| EPI_ISL_966411 | OCME Office Of Chief Medical Examiner | New York City Public Health Laboratory | Jade Wang, et al. |
| EPI_ISL_966412 | DOHMH Jamaica | New York City Public Health Laboratory | Jade Wang, et al. |
| EPI_ISL_966413 | DOHMH Central Harlem | New York City Public Health Laboratory | Jade Wang, et al. |
| EPI_ISL_966414 | DOHMH Morrisania | New York City Public Health Laboratory | Jade Wang, et al. |
| EPI_ISL_966415 | DOHMH Corona | New York City Public Health Laboratory | Jade Wang, et al. |

| | | | |
|---|---|---|---|
| EPI_ISL_966416 | DOHMH Central Harlem | New York City Public Health Laboratory | Jade Wang, et al. |
| EPI_ISL_966417, EPI_ISL_966418, EPI_ISL_966419 | OCME Office Of Chief Medical Examiner | New York City Public Health Laboratory | Jade Wang, et al. |
| EPI_ISL_966420 | DOHMH Morrisania | New York City Public Health Laboratory | Jade Wang, et al. |
| EPI_ISL_966421, EPI_ISL_966422 | DOHMH Jamaica | New York City Public Health Laboratory | Jade Wang, et al. |
| EPI_ISL_966423 | DOHMH Morrisania | New York City Public Health Laboratory | Jade Wang, et al. |
| EPI_ISL_966424 | DOHMH Jamaica | New York City Public Health Laboratory | Jade Wang, et al. |
| EPI_ISL_966425 | DOHMH Corona | New York City Public Health Laboratory | Jade Wang, et al. |
| EPI_ISL_966426 | DOHMH PHL | New York City Public Health Laboratory | Jade Wang, et al. |
| EPI_ISL_966427 | OCME Office Of Chief Medical Examiner | New York City Public Health Laboratory | Jade Wang, et al. |
| EPI_ISL_981098, EPI_ISL_981099 | Johns Hopkins Hospital Department of Pathology | Johns Hopkins Hospital Department of Pathology | C. Paul Morris, Chun Huai Luo, Adannaya Amadi, Matthew Schwartz, Nicholas Gallagher, Heba H. Mostafa |
| EPI_ISL_982440, EPI_ISL_982442, EPI_ISL_982458, EPI_ISL_982461, EPI_ISL_982465, EPI_ISL_982489, EPI_ISL_983103, EPI_ISL_983114, EPI_ISL_983115 | MONTEFIORE MEDICAL CENTER LABORATORIES | Wadsworth Center, New York State Department of Health | Kirsten St. George, Daryl M. Lamson, Alexis Russel, Matthew Shudt, Melissa A Leisner, Jonathan Plitnick, Navjot Singh, John Kelly, Erasmus Schneider, Erica Lasek-Nesselquist |
| EPI_ISL_984771, EPI_ISL_984772, EPI_ISL_984773, EPI_ISL_984816, EPI_ISL_984817, EPI_ISL_984870, EPI_ISL_984871, EPI_ISL_984883, EPI_ISL_984893, EPI_ISL_984897, EPI_ISL_984898, EPI_ISL_984909, EPI_ISL_984913, EPI_ISL_984927, EPI_ISL_984928, EPI_ISL_984929, EPI_ISL_984930, EPI_ISL_984947, EPI_ISL_994803, EPI_ISL_994804, EPI_ISL_994819, EPI_ISL_994824, EPI_ISL_994826, EPI_ISL_994830, EPI_ISL_994835, EPI_ISL_994838, EPI_ISL_994839, EPI_ISL_994840, EPI_ISL_994841, EPI_ISL_994844, EPI_ISL_994848, EPI_ISL_994853, EPI_ISL_994870, EPI_ISL_994872, EPI_ISL_994876, EPI_ISL_994879, EPI_ISL_994880, EPI_ISL_994883, EPI_ISL_994884, EPI_ISL_994886, EPI_ISL_994890, EPI_ISL_994895, EPI_ISL_994896, EPI_ISL_994898, EPI_ISL_994900, EPI_ISL_994903, EPI_ISL_994906, EPI_ISL_994911, EPI_ISL_994928, EPI_ISL_994929, EPI_ISL_994930, EPI_ISL_994931, EPI_ISL_994943, EPI_ISL_994945, EPI_ISL_994946, EPI_ISL_994952, EPI_ISL_994958, EPI_ISL_994960, EPI_ISL_994961, EPI_ISL_994966, EPI_ISL_994967, EPI_ISL_994968, EPI_ISL_994970, EPI_ISL_994971, EPI_ISL_994974, EPI_ISL_994975, EPI_ISL_994976, EPI_ISL_994981, EPI_ISL_994985, EPI_ISL_994986, EPI_ISL_994988, EPI_ISL_994989, EPI_ISL_994993, EPI_ISL_994994, EPI_ISL_995001, EPI_ISL_995004, EPI_ISL_995006, EPI_ISL_995010, EPI_ISL_995015, EPI_ISL_995017, EPI_ISL_995020, EPI_ISL_995023, EPI_ISL_995024, EPI_ISL_995025, EPI_ISL_995026, EPI_ISL_995032, EPI_ISL_995037, EPI_ISL_995040, EPI_ISL_995041, EPI_ISL_995042, EPI_ISL_995043, EPI_ISL_995046, EPI_ISL_995048, EPI_ISL_995056, EPI_ISL_995057, EPI_ISL_995060, EPI_ISL_995067, EPI_ISL_995071, EPI_ISL_995072, EPI_ISL_995076, EPI_ISL_995087, EPI_ISL_995089, EPI_ISL_995102, EPI_ISL_995110, EPI_ISL_995113, EPI_ISL_995114, EPI_ISL_995118, EPI_ISL_995119, EPI_ISL_995123, EPI_ISL_995131, EPI_ISL_995133, EPI_ISL_995138, EPI_ISL_995139, EPI_ISL_995140, EPI_ISL_995143, EPI_ISL_995144, EPI_ISL_995145, EPI_ISL_995146, EPI_ISL_995148, EPI_ISL_995155, EPI_ISL_995157 | | | |
| see above | Pandemic Response Lab - NYC | Pandemic Response Lab, R&D | Henry Lee, Michael Hammerling, Melissa Hopkins, Cybill del Castillo, William Ward, Pradeep Bugga, Haiping Hao, Jon Laurent |
